# Development and validation of a federated learning framework for detection of subphenotypes of multisystem inflammatory syndrome in children

**DOI:** 10.1101/2024.01.26.24301827

**Authors:** Naimin Jing, Xiaokang Liu, Qiong Wu, Suchitra Rao, Asuncion Mejias, Mitchell Maltenfort, Julia Schuchard, Vitaly Lorman, Hanieh Razzaghi, Ryan Webb, Chuan Zhou, Ravi Jhaveri, Grace M. Lee, Nathan M. Pajor, Deepika Thacker, L. Charles Bailey, Christopher B. Forrest, Yong Chen

## Abstract

**Background:** Multisystem inflammatory syndrome in children (MIS-C) is a severe post-acute sequela of SARS-CoV-2 infection. The highly diverse clinical features of MIS-C necessities characterizing its features by subphenotypes for improved recognition and treatment. However, jointly identifying subphenotypes in multi-site settings can be challenging. We propose a distributed multi-site latent class analysis (dMLCA) approach to jointly learn MIS-C subphenotypes using data across multiple institutions.

**Methods:** We used data from the electronic health records (EHR) systems across nine U.S. children’s hospitals. Among the 3,549,894 patients, we extracted 864 patients < 21 years of age who had received a diagnosis of MIS-C during an inpatient stay or up to one day before admission. Using MIS-C conditions, laboratory results, and procedure information as input features for the patients, we applied our dMLCA algorithm and identified three MIS-C subphenotypes. As validation, we characterized and compared more granular features across subphenotypes. To evaluate the specificity of the identified subphenotypes, we further compared them with the general subphenotypes identified in the COVID-19 infected patients.

**Findings:** Subphenotype 1 (46.1%) represents patients with a mild manifestation of MIS-C not requiring intensive care, with minimal cardiac involvement. Subphenotype 2 (25.3%) is associated with a high risk of shock, cardiac and renal involvement, and an intermediate risk of respiratory symptoms. Subphenotype 3 (28.6%) represents patients requiring intensive care, with a high risk of shock and cardiac involvement, accompanied by a high risk of >4 organ system being impacted. Importantly, for hospital-specific clinical decision-making, our algorithm also revealed a substantial heterogeneity in relative proportions of these three subtypes across hospitals. Properly accounting for such heterogeneity can lead to accurate characterization of the subphenotypes at the patient-level.

**Interpretation:** Our identified three MIS-C subphenotypes have profound implications for personalized treatment strategies, potentially influencing clinical outcomes. Further, the proposed algorithm facilitates federated subphenotyping while accounting for the heterogeneity across hospitals.

**Research in context panel:** *Evidence before this study:* Before undertaking this study, we searched PubMed and preprint articles from in early 2022 for studies published in English that investigated the clinical subphenotypes of MIS-C using the terms “multi-system inflammatory syndrome in children” or “pediatric inflammatory multisystem syndrome”, and “phenotypes”. One study in 2020 divided 63 patients into Kawasaki and non-Kawasaki disease subphenotypes. Another CDC study in 2020 evaluated 3 subclasses of MIS-C in 570 children, with one class representing the highest number of organ systems, a second class with predominant respiratory system involvement, and a third class with features overlapping with Kawasaki Disease. However, both studies were conducted during the early phase of the pandemic when misclassification of cases as Kawasaki disease or acute COVID-19 may have occurred. Therefore, the subphenotypes of MIS-C needs further investigation. In addition, we searched research articles for studies published in English on algorithms for distributed multi-site latent class analysis with the terms “distributed latent class analysis” or “multi-site latent class analysis”. Most of the existing literatures for distributed learning have focused on supervised learning. Literatures discuss latent class analysis for disease sub phenotyping in a multi-site setting where data are distributed across different sites are lacking.

*Added value of this study:* We developed a new algorithm to jointly identify subphenotypes of MIS-C using data across multiple institutions. Our algorithm does not require individual-level data sharing across the institutions while achieves the same result as when the data are pooled. Besides, our algorithm properly accounts for the heterogeneity across sites, and it can lead to accurate characterization of the subphenotypes at the patient-level. We then applied our algorithm to PEDSnet data for identifying the subphenotypes of MIS-C. PEDSnet provides one of the largest MIS-C cohorts described so far, providing sufficient power for detailed analyses on MIS-C subphenotypes. We identified three subphenotypes that can be characterized as mild with minimal cardiac involvement (46.1%), severe requiring intensive care with >4 organ being impacted, and the one with intermediate risk of respiratory symptoms, and high risk of shock, cardiac and renal involvement (25.3%). For hospital-specific clinical decision-making, our algorithm revealed a substantial heterogeneity in relative proportions of these three subtypes across hospitals.

*Implications of all the available evidence:* Our algorithm provides an effective distributed learning framework for disease subphenotyping using multi-site data based on aggregated data only. It facilitates high accuracy while properly accounts for the between-site heterogeneity. The results provide an update to the subphenotypes of MIS-C with larger and more recent data, aid in the understanding of the various disease patterns of MIS-C, and may improve the evaluation and intervention of MIS-C.

## Introduction

Multisystem inflammatory syndrome in children (MIS-C) is a form of post-acute sequelae of SARS-CoV-2 infection (PASC). It is a rare but severe post-infectious hyperinflammatory disorder happening in children and adolescents < 21 years of age that occurs 2-6 weeks after the infection of SARS-CoV-2. The clinical features of MIS-C are diverse. Commonly observed clinical features include fever, gastrointestinal symptoms, cardiac complications, mucocutaneous, and respiratory symptoms, with some features overlapping with Kawasaki disease or macrophage activation syndrome.^1^ The highly heterogeneous and complex spectrum of clinical features makes the diagnoses of MIS-C difficult. Therefore, characterizing its disease patterns by subphenotypes is essential to learn the co-occurrence pattern of the conditions, understand the pathophysiological mechanisms underlying the clinical presentations, and eventually improve its recognition and intervention.

The latent class analysis (LCA) model is a widely used and effective method for disease subphenotyping.^2–9^ However, MIS-C is rare while LCA models often involve many parameters that require sufficient samples to guarantee an accurate estimate. In the MIS-C study supported by the PEDSnet,^12, 13^ the smallest hospital only identified 32 MIS-C patients, which is far from being sufficient for obtaining reliable LCA results. One promising strategy is to integrate data from multiple hospitals to increase the sample size toward reliable disease subphenotyping. However, individual-level data sharing is usually prohibited due to regulatory restrictions and privacy policies. An effective approach is federated learning,^14^ a method designed to analyze distributed data with only summary-level statistics (i.e., aggregated data) being shared across the institutions. However, to the best of our knowledge, there is currently a lack of federated learning algorithms for subphenotyping. Further, it is important to properly account for between-site heterogeneity due to the intrinsic differences in characteristics of patients across hospitals, which can lead to accurate characterization of the subphenotypes at the patient-level. From a clinical perspective, the current understanding of MIS-C subphenotypes is limited, with few earlier studies.^1,2^ Therefore, there is still a critical need to develop an effective federated learning algorithm for LCA, and to characterize MIS-C subphenotypes using this novel method with decentralized data sets.

Data integration can significantly enhance the precision and power of clinical studies by providing large and representative study samples. In recent years, numerous efforts have been made in developing large clinical research networks (e.g., OHDSI,^15^ PEDSnet) and data harmonization tools^16^ to make multi-site data integration possible. In addition, in the last decades, federated learning algorithms have been developed for analyzing distributed data in a privacy-aware and heterogeneity-aware manner.^17–32^ Nevertheless, most of the existing methods have focused on supervised learning, while less attention has been paid to unsupervised clustering tasks, such as subphenotyping via the LCA. Different from supervised learning, the traditional divide-and-conquer strategy (e.g., meta-analysis) cannot be directly applied to subphenotyping with several unique challenges. A prominent challenge is that applying LCA separately at different sites does not guarantee the same number of latent classes, which raises significant difficulty in synthesizing the results. Further, even with the same number of latent classes, the identified latent classes at different sites may not be comparable. As an alternative to divide-and-conquer strategy, novel federated learning algorithms that warrant coherent definitions of the latent classes while account for between-site heterogeneity are critically needed.

In this paper, we developed a federated learning framework for disease subphenotyping and applied it to detect subphenotypes of MIS-C. Different from the divide-and-conquer method, our federated learning algorithm circumvents the class-match issue by collaboratively learning a common set of latent classes shared across all sites. What’s more, between-site heterogeneity can also be properly modeled and handled. We showed the validity and the strength of the proposed method through extensive real-data guided simulation studies. We then applied the proposed algorithm to a large electronic health records (EHR) data cohort and identified three clinically significant MIS-C subphenotypes. We conducted two independent validation studies. First, for identified clinical subtypes, we characterized and compared the demographics and around sixty more granular clinical features across three subphenotypes. Second, we also compared the identified MIS-C subphenotypes with the general subphenotypes in the COVID-19 infected patients. And our study found that our identified MIS-C subphenotypes are truly *de novo* and specific to the MIS-C population. All these findings demonstrated the essential role of our method in identifying disease subphenotypes in multi-site settings. We concluded the paper by discussing strengths and limitations of our method.

## Methods

### Study design and participants

In this retrospective and multi-site cohort study, we developed, trained, and validated a federated learning algorithm that was designed to detect subphenotypes of MIS-C using the EHR data from nine pediatric hospitals in the United States. The data were retrieved from the following nine PEDSnet institutions, which is a national collaboration of pediatric health systems that share EHR data, conduct research, and improve outcomes together: Children’s Hospital of Philadelphia, Cincinnati Children’s Hospital Medical Center, Children’s Hospital Colorado, Ann & Robert H. Lurie Children’s Hospital of Chicago, Nationwide Children’s Hospital, Nemours Children’s Health System (a Delaware and Florida health system), Seattle Children’s Hospital, and Stanford Children’s Health).

We included encounters from children and adolescents < 21 years of age who had visited the nine hospitals between March 2020 and December 2021, with a total of 3,549, 894 patients. To identify the hospitalized MIS-C cases, we used the International Classification of Diseases, Tenth Revision, Clinical Modification (ICD-10-CM) code for “MIS-C” or “Other specified systemic involvement of connective tissue”. We identified a final study cohort of 864 hospitalized MIS-C patients. For details of the cohort selection process refer to **Figure A2**.

We developed ten clinical dichotomous variables to characterize MIS-C patients, including SARS-CoV-2 infection, SARS-CoV-2 exposure 42 days before or during the inpatient stay, shock, and involvement of the following organ systems: cardiac, gastrointestinal, hematological, neurologic, renal, respiratory, and dermatologic. Our evaluation time window of the organ system involvements and shock was during hospitalization or 1 day prior to the hospitalization. A particular organ system involvement was positive if at least one condition, abnormal laboratory result, or procedure related to that system was positive within the specified time window. The detailed information on the condition, laboratory results, and procedures used to define each variable is in **Table A2**. In addition, we have included the code sets used to define all the related conditions, laboratory results, and procedures in the Supplementary Material. The code sets were developed with the effort of a group of clinicians in pediatric diseases.

The use of data was approved by the Children’s Hospital of Philadelphia’s Institutional Review Board (IRB). The informed consent was waived.

### Procedures

Latent class analysis (LCA) is a well-suited approach for our purpose of MIS-C subphenotyping. LCA takes in discrete manifest variables and identifies latent classes (i.e., subphenotypes) based on class-specific co-occurrence pattern among manifest variables. Each latent class can be characterized by a vector of prevalence of the manifest variables within this latent class, which represents a specific pattern of distribution.

To jointly conduct LCA using data from multiple sites, we developed a Distributed Multi-site LCA approach (dMLCA). The proposed dMLCA successfully addresses two key challenges when applying LCA in multi-site settings: first, a common set of latent classes shared by all sites are defined to solve the class-match issue. Second, site-specific class mixing proportions are allowed to account for the between-site heterogeneity in the study population. In **Figure 1**, we illustrated the between-site heterogeneity and how this can be unfolded by the shared latent classes and site-specific class proportions. To be concrete, in MIS-C data analysis where we used ten clinical dichotomous variables to characterize MIS-C patients, the prevalence of these variables within each institution indicates substantial heterogeneity of population across sites (see **Table A1**). Since the subphenotypes of a disease are expected to reflect the intrinsic clinical mechanisms of the disease which do not rely on the hospitals, the subphenotypes should be consistent across sites. By using the varying mixing proportions of subphenotypes across sites, our dMLCA indeed provides a reasonable framework to explain the between-site heterogeneity of populations while retains the consistency of subphenotypes across sites.

**Figure 1.**
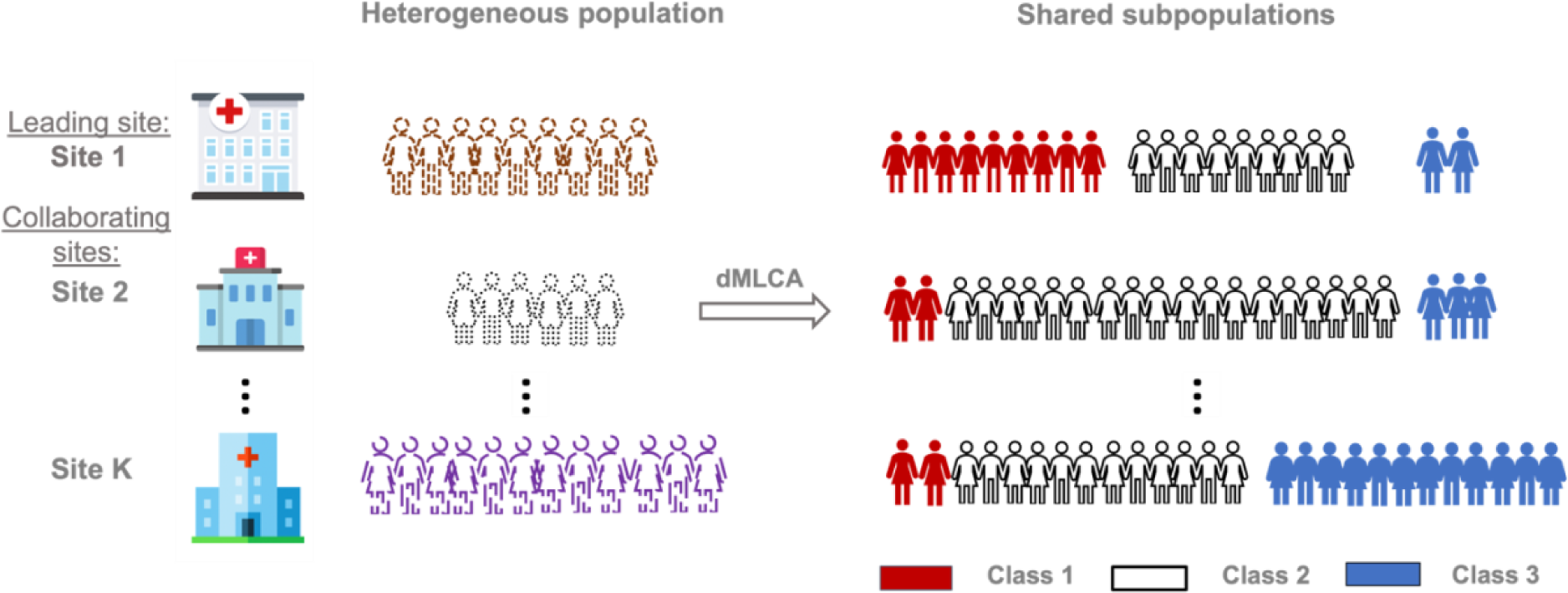
Illustration of the shared subphenotypes, the heterogeneous population across sites and how the varying proportions/prevalence of the subphenotypes account for the heterogeneity. The populations across sites may appear very different (represented by different outlines; see the second column – “Heterogeneous population”), and this overall between-site heterogeneity is unfolded by latent classes (see the third column – “Shared subpopulations”). Suppose the population contains three distinct subphenotypes. Population in Site 1 is an even mixture of two of the subphenotypes (i.e., classes 1 and 2) and with a small portion of the remaining subphenotype, while Site 2 contains a large portion of a subphenotype (i.e., class 2) and small portions of the remaining subphenotypes (i.e., classes 1 and 3). Heterogeneity in the prevalence of the subphenotypes across sites explains the heterogeneity in the population.

Properly handling the between-site heterogeneity is helpful to improve the estimation and prediction accuracy in multi-site analysis. Most current work that uses centralized data, i.e., pooled data analysis, ignores the between-site heterogeneity and simply applies the standard^6, 1^ on the pooled data, which could lead to biased results.^33,34^ In contrast, with dMLCA, the between-site heterogeneity is carefully handled, which can provide us with improved estimation accuracy compared to centralized LCA and further facilitates higher prediction accuracy for patient class membership.

In multi-site studies, individual-level data-sharing is often prohibited due to privacy concerns, necessitating the use of federated learning algorithms. In the MIS-C data analysis, we can access the individual-level data at the Children’s Hospital of Philadelphia (CHOP), while other participant sites might not be able to timely provide individual-level data. To meet the analysis need without sharing individual-level data, we developed a distributed Expectation-Maximization (EM) algorithm requiring only individual-level data from a lead site (i.e., CHOP) and aggregated data from all participant sites to fit dMLCA. EM algorithm^35^ is designed to deal with missing data and therefore is appropriate here by treating the unobserved subphenotypes as missing. At each iteration of the EM algorithm to update the parameters, a key observation to guarantee *lossless estimation* while handling the data-sharing prohibition is that the updating formulas are *exactly decomposable* by sites. Therefore, at each iteration, each site only needs to calculate the decomposable part using its local data and transfer the results (which are aggregated data) to the lead site to update the estimates. No patient-level data sharing is needed in this procedure.

Applying dMLCA can improve patient-level class membership prediction. With the dMLCA fitting results, the class membership for each patient can be inferred from the posterior probability which is the probability of each patient belonging to a certain latent class. The benefits of applying dMLCA to improve patient-level class membership prediction can be displayed much clearer with the formula of calculating the posterior probability:

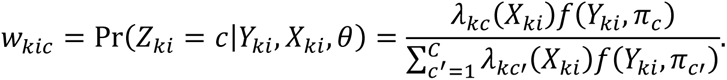

First, the prevalence of manifest variables *Y*_*ki*_ within each latent class *π*_*c*_ is learned jointly in dMLCA, which provides higher estimation accuracy than the single-site analysis and has smaller bias than the divide-and-conquer method; second, the site-specific class mixing proportion *λ*_*kc*_ solely employed in dMLCA takes into account unobserved site-specific factors into consideration, while pooled analysis simply ignores this information; third, when the effects from patient-level covariates *X*_*ki*_, e.g., age, on class membership prediction are of interest, dMLCA allows for a regression model *λ*_*kc*_(*X*_*ki*_) to jointly learn these effects. Then, the class with the maximum posterior can be used as the inferred class membership of a patient.

We summarize the differences between dMLCA and several existing methods in **Table 1**. Our simulation studies have demonstrated that dMLCA outperforms other methods with greater estimation and prediction accuracy. Details and results of the simulation studies are provided in the **Supplemental Material**.

**Table 1.** Comparison of the proposed dMLCA and several existing methods.

|  | Methods |  |  |  |
| --- | --- | --- | --- | --- |
|  | Centralized LCA | Single-site LCA | Divide-and-conquer | dMLCA |
| **Aggregated-data based** | No | Yes | Yes | **Yes** |
| **No class-match issue** | Yes | N/A | No | **Yes** |
| **Heterogeneity-aware** | No | N/A | No | **Yes** |
| **Accurate class membership prediction** | No | Yes | No | **Yes** |
| **Accurate estimation** | Yes | No | No | **Yes** |

### Statistical analysis

LCA requires a pre-specified number of latent classes. To select the number of latent classes, we inspected several model fitting criteria including Akaike Information Criterion (AIC)^36^ and Bayesian Information Criterion (BIC).^37^ We fitted the model with two to six classes and evaluated the corresponding AIC and BIC values (**Table A3**). While AIC continued to decrease as the number of classes increased, BIC favored three latent classes most. To guarantee the interpretability of the latent classes, we also took into account clinicians’ judgement and expertise in selecting the optimal number of classes and decided the number of latent classes to be three. We randomly generated 10 initial values to run dMLCA with three latent classes and reported the one with the maximum likelihood as the final estimator. The algorithm was run in R version 4.1.2 (2021-11-01). As sensitivity analysis, we also provided the analysis results with four latent classes in **Figure A3** in the **Supplementary Material**.

To validate the analysis results, we further characterized and compared the three identified latent classes by using demographic variables (age, race/ethnicity, gender, ICU admission, death, PMCA index, Pre-admission obesity) and several more granular clinical features, including a list of relevant medications, and the conditions, diagnoses, and lab results that were used to define organ systems. Specifically, medications, conditions, abnormal lab results and procedures were coded as dichotomous and were positive if occurred during the evaluation time window. For each variable, we calculated its prevalence in each latent class through an estimated average weighted by the patients’ posterior class membership probabilities (see **Figure 4**).

In addition, we compared the identified MIS-C latent classes with the latent classes identified from the acute COVID-19 but non-MIS-C cohort. This comparison was made to validate that our identified MIS-C subphenotypes are truly *de novo* and specific to the MIS-C population and to demonstrate that the selection criteria for the MIS-C cohort were indeed valid. Specifically, we identified three latent classes for children testing positive for COVID-19 by PCR without MIS-C diagnosis by applying LCA to 59,740 children (see **Panel A) of Figure 5**). As for the manifest variables used for COVID-19 subphenotyping, except that we removed SARS-CoV-2 infection and exposure since all the children in this cohort were infected, we used the same manifest variables as in the MIS-C analysis but with an evaluation time window of −7 to 28 days from the first positive test date. We compared the two sets of subphenotypes identified from the MIS-C and COVID-19 cohorts, respectively, to see their difference. What’s more, we visualized the distances between COVID-19 and MIS-C subpopulations (see **Panel B) of Figure 5**). Specifically, patients were grouped into latent classes according to their maximum posterior class membership probabilities, then the distance between each pair of latent classes was measured by fixation index^38^ and mapped onto a 2-dimensional plot through multidimensional scaling.^39^

## Results

Among 3,549,894 patients with age < 21 who visited any of the nine PEDSnet hospitals during March 2020 and December 2021, 1,180 were diagnosed as MIS-C, and 864 were hospitalized when the MIS-C code was assigned or 1 day after the MIS-C code was assigned (**Figure A2**). **Table 2** describes the characteristics of the study cohort. The hospital names and the numbers that are less than 11 were blinded to protect patient data. The majority of children with MIS-C were between the ages of 16 and 20 and identified as non-Hispanic White. Among the hospitals included in the study, Hospital B had the highest number of MIS-C cases, while Hospital F had the least with only 32 cases. It’s also worth noting that 44% of the MIS-C patients were admitted to an ICU during their hospitalization.

**Table 2.** Characteristics of the study cohort.

| Characteristics |  | Number of patients (proportion)<br>(N = 864) |
| --- | --- | --- |
| Age at MIS-C diagnosis, years | <1 | 30(3.5%) |
|  | 1-4 | 207 (24.0%) |
|  | 5-11 | 171 (19.8%) |
|  | 12-15 | 79 (9.1%) |
|  | 16-20 | 377 (43.6%) |
| Sex | Female | 343 (39.7%) |
|  | Male | 521 (60.3%) |
| Race/Ethnicity | Hispanic | 176 (20.4%) |
|  | Non-Hispanic White | 345 (39.9%) |
|  | Non-Hispanic Black/African-American | 225 (26.0%) |
|  | Non-Hispanic Asian/Pacific Islander | 33 (3.8%) |
|  | Other/Unknown | 46 (5.3%) |
|  | Multiple | 39 (4.5%) |
| Institution | A | 113 (13.1%) |
|  | B | 189 (21.9%) |
|  | C | 49 (5.7%) |
|  | D | 113 (13.1%) |
|  | E | 73 (8.4%) |
|  | F | 32 (3.7%) |
|  | G | 143 (16.6%) |
|  | H | 65 (7.5%) |
|  | I | 87 (10.1%) |
| Obesity | Yes | 100 (11.6%) |
| Admitted to intensive care unit | Yes | 380 (44.0%) |
| Vital status | Yes | <11 (<1.3%) |
| Mechanical ventilation | Yes | 144 (16.7%) |

The dMLCA separated the complex MIS-C cohort into three clinically interpretable subphenotypes with different disease presentations. The resulting subphenotypes, as depicted in **Panel A) of Figure 2**, are characterized as follows: Class 1 represents patients with a milder form of MIS-C not necessitating intensive care and with no or minimal cardiac involvement. Class 2 comprises children exhibiting a severe presentation of MIS-C alongside cardiac system involvement and less than four involved organ systems; lastly, Class 3 represents children with the most severe form of MIS-C showcasing cardiac involvement as well as over four impacted systems, including respiratory, gastrointestinal (GI), renal, hematologic, and dermatological manifestations. These findings have important clinical implications, suggesting that children with GI presentations or skin rashes may have an increased risk of more severe disease, thereby warranting closer monitoring and earlier treatment. The mean posterior probabilities of membership of the three latent class were 0·841, 0·802 and 0·874, respectively, which were large and therefore showed that the classes were well-separated.

**Figure 2.**
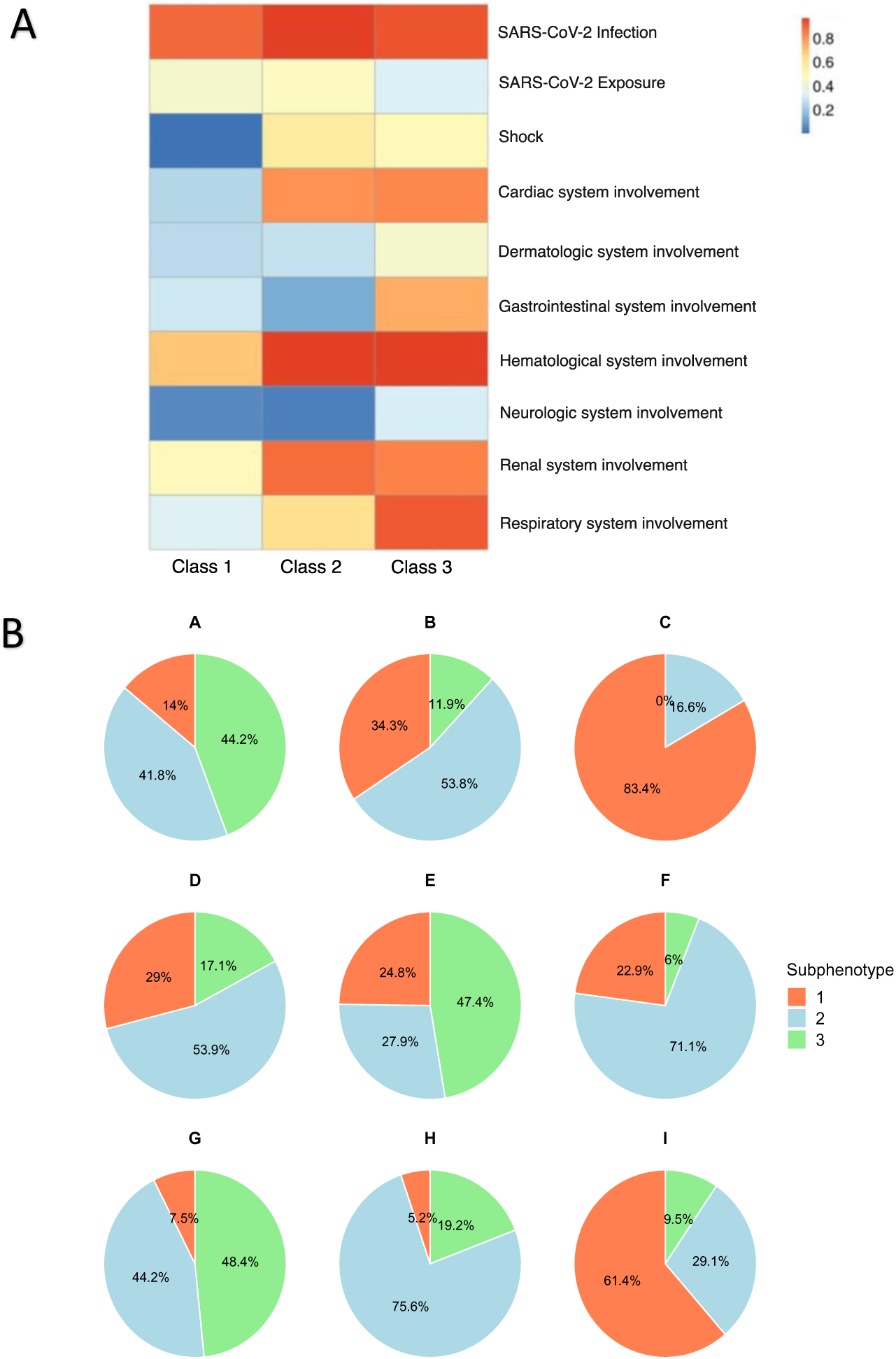
Results of MIS-C data analysis using dMLCA with three latent classes. **A)** A heatmap showing the prevalence of ten manifest variables in three latent classes; Each column represents a latent class, and each row represents a manifest variable. The color of the boxes represents the prevalence. The legend on the top right shows the scale of the colors. Red represents prevalence close to 100% and blue represents prevalence close to 0%. Class 1 corresponds to patients with a milder presentation of MIS-C not requiring intensive care, with no or minimal cardiac involvement. Class 2 represents children with a severe presentation of MIS-C, with cardiac system involvement and < 4 organ systems involved, and Class 3 represents children with the more severe presentation of MIS-C including cardiac involvement along with > 4 systems involved, including respiratory, gastrointestinal (GI), renal, hematologic, and dermatological manifestations. **B)** Pie charts showing the prevalence of the three latent classes by site.

As for the whole population across all the nine sites, the estimated class mixing proportions (i.e., class prevalence) were 46·1%, 25·3%, and 28·6% for Class 1, 2 and 3, respectively.Besides, dMLCA estimated the site-specific class mixing proportions to help understand the population composition at each site. **Panel B) of Figure 2** presents the prevalence of the classes at each of the nine sites (A-I), highlighting that the population composition varied significantly across sites. For example, 42·7% of patients in Site A would fall into Class 2, while only 11·0% of patients in Site B were in Class 2, and in Site C almost no patients fell into Class 2. These differences are likely due to variations in patient populations including race/ethnicity distributions, in patient acuity as some centers may have captured a more selective population, and in evaluation and treatment protocols in place across the different healthcare systems for children with MIS-C.

In addition, we demonstrated that the data integration in dMLCA was lossless by comparing the results obtained from dMLCA to those obtained from fitting LCA with site-specific mixing proportions on pooled data, i.e., the centralized patient-level data supposing data-sharing is allowed. **Figure 3** showed that the estimated parameters in Class 1 from dMLCA using aggregated data only are identical to those obtained from pooled data analysis. Results regarding Class 2 and 3 are the same and are given in the Supplemental Material.

**Figure 3.**
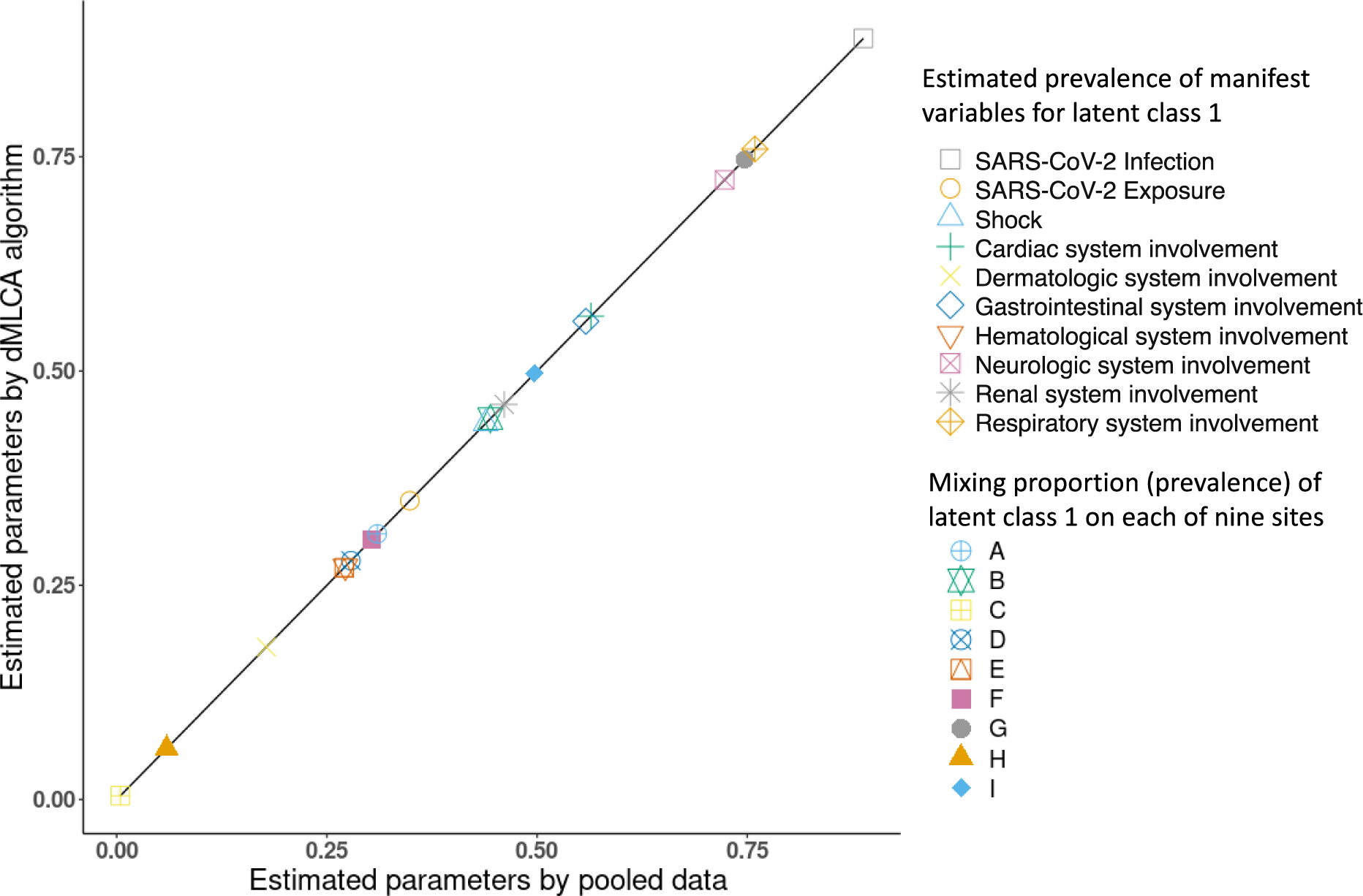
Prevalence of manifest variables for latent class 1 and mixing proportion (prevalence) of latent class 1 estimated by dMLCA algorithm and the pooled data in the MIS-C data analysis (the estimates for latent classes 2-3 are deferred to **Figure A1** in the Supplementary Materials).

As a validation, the subphenotypes were further characterized in **Figure 4**. The classes share close distributions of race and ethnicity, except for the black/African American race which is more prevalent in Classes 2 and 3 and has the highest proportion in Class 3. Class 3 also exhibited a larger proportion of children aged 12 years and older and children with complex chronic conditions. ICU admissions were higher in Classes 2 and 3. The patterns of the specific diagnoses, lab results, and medications were consistent with the co-occurrence patterns of the ten organ system involvement indicator variables found in the primary analysis as shown in **Figure 2**.

**Figure 4.**
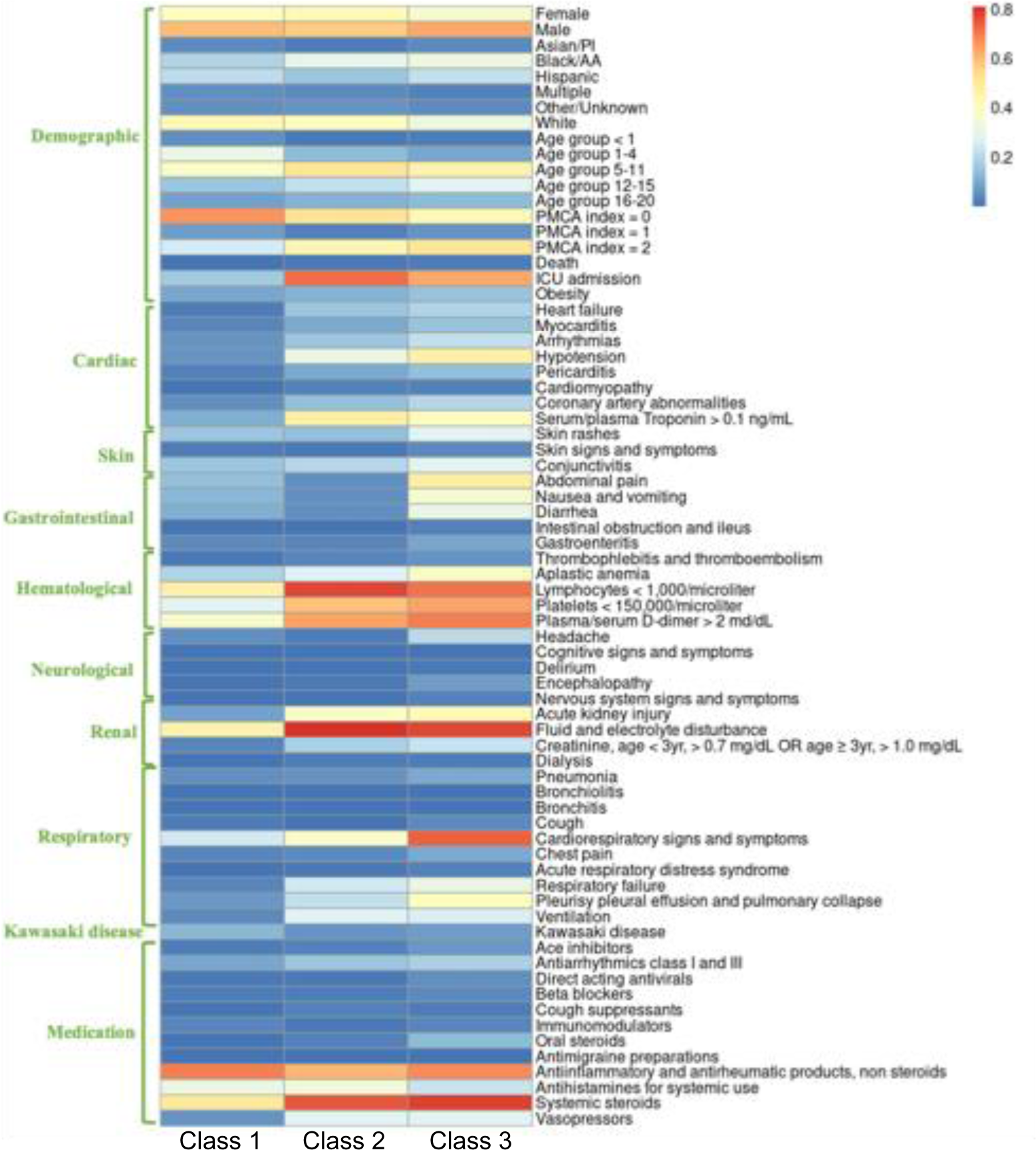
A heatmap showing the prevalence of over 60 demographic, condition, lab and medication variables in the three latent classes found in MIS-C data analysis; Each column represents a latent class, and each row represents a variable. The calculation of prevalence is weighted by posterior class membership probabilities. The color of the boxes represents the prevalence. The legend on the top right shows the scale of the colors. Red represents prevalence close to 100% and blue represents prevalence close to 0%.

We further confirmed the uniqueness of the identified MIS-C latent classes by comparing them with the subphenotypes identified in the general COVID-19 cohort. In **Figure 5, Panel A)**, we presented the characteristics of latent classes of COVID-19 PCR-positive children without MIS-C diagnoses, which were markedly different from the characteristics of the MIS-C latent classes. To further emphasize the separation between the COVID-19 and MIS-C subpopulations, we generated a visual representation of their distances in **Panel B) of Figure 5**. In general, the latent classes of the two cohorts were distinctly separated from each other, with COVID-19 subphenotypes located at the left side of the figure and MIS-C subphenotypes located at the right part of the figure.

**Figure 5.**
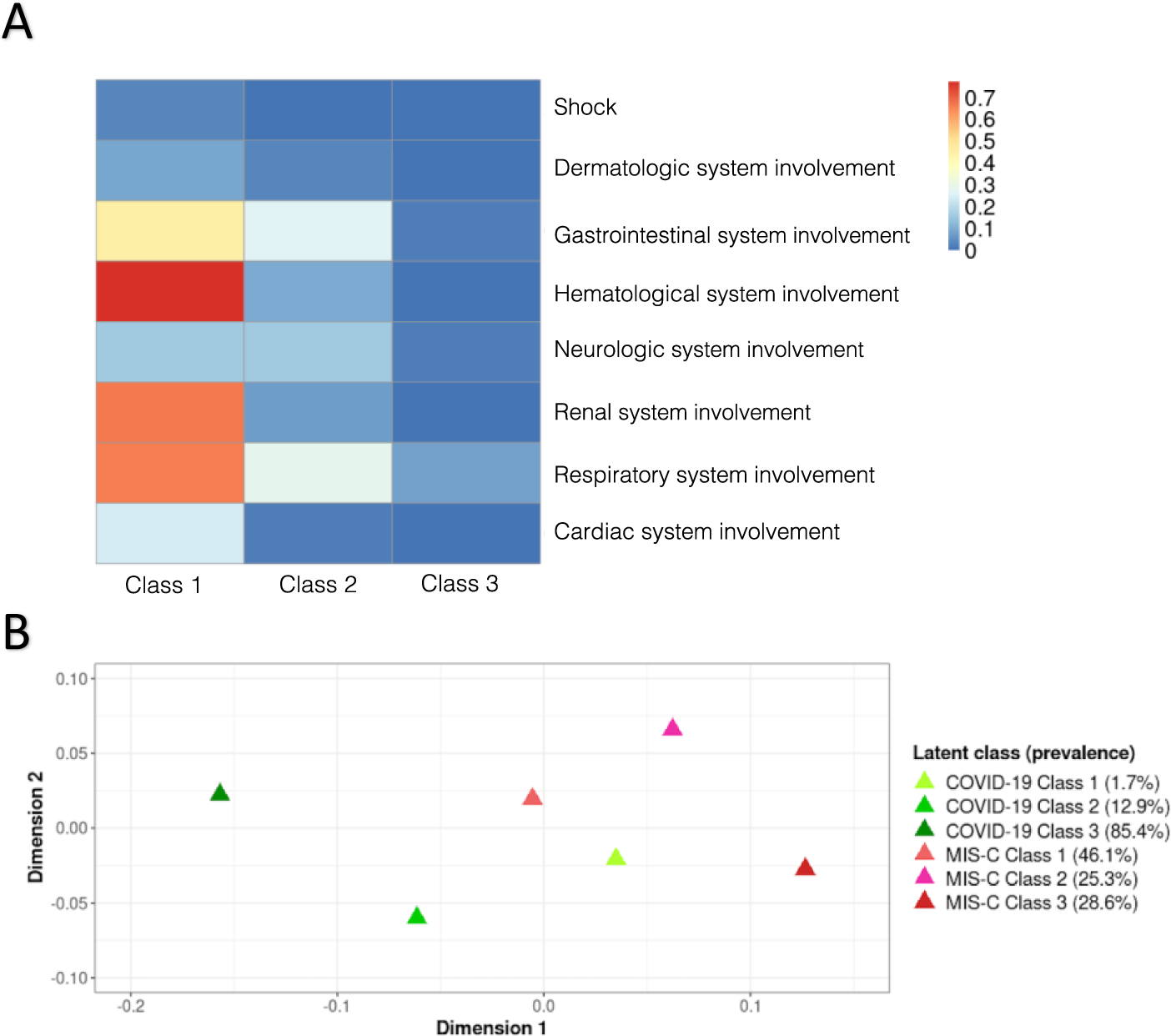
**A)** A heatmap showing the latent classes and their characteristics of children testing positive for SARS-CoV-2 by PCR test. **B)** A 2-Dimensional plot comparing the distances among MIS-C and COVID-19 PCR positive subpopulations. Closer subpopulations have larger similarities. The distance between each pair of latent classes was measured by fixation index (Fst) and mapped onto a 2-dimensional plot through multidimensional scaling.

## Discussion

We developed and validated a federated learning approach, dMLCA, which enables detecting subphenotypes of a disease using data from multiple sites when patient-level data sharing is not allowed. We applied dMLCA to analyze data from nine hospitals and identified three subphenotypes of MIS-C. Compared to divide-and-conquer methods, our dMLCA eliminates potential biases due to inconsistent number of classes or class mismatching, while allowing for site-specific parameters to account for between-site heterogeneity that is often overlooked in existing methods. More importantly, our estimation is statistically lossless compared to the results based on the pooled patient-level data. Overall, dMLCA represents a promising approach for disease subphenotyping that can overcome barriers in data sharing across multiple sites.

One of the major strengths of dMLCA is its novel formulation, which warrants coherent definitions of the latent classes while account for between-site heterogeneity. Specifically, it reduces estimation bias that can occur due to class mismatching for divide-and-conquer type of methods by defining a set of subphenotypes that are common across multiple datasets. Further, dMLCA also accounts for between-site heterogeneity, which is commonly observed in multi-site studies. By allowing for site-specific prevalence of subphenotypes, dMLCA can capture the unique characteristics and differences between different hospitals. This is a critical feature that can explain the heterogeneity and lead to more accurate class membership estimation.

The second advantage of the dMLCA algorithm is its capacity to achieve lossless information integrate across multiple hospitals based on aggregated data only. Since all the calculations involved in EM algorithms are exactly decomposable in terms of hospitals, the dMLCA estimation procedure only requires each participating hospital carry out its calculations locally and send the aggregated information to the leading site. This approach guarantees that the estimation results achieved are equivalent to those obtained when data sharing is allowed. Moreover, the privacy-aware feature of dMLCA is also achieved since no patient-level data is exchanged across different institutions.

Despite these strengths, our dMLCA algorithm has some limitations that warrants further investigations. First, our dMLCA is based on aggregated data only. Such algorithm is applicable to settings similar to the well-known GLORE^19^ and WebDISCO^22^ algorithms for federated logistic regression of binary outcome and Cox proportional hazard model of time-to-event outcomes, respectively. In the future, formal privacy techniques, such as k-anonymity,40 differential privacy,^40, 41^ homomorphic encryption^42^ and multi-party encryptions, ^43^ can be integrated into the step of shared aggregated data. Secondly, similar to the popular GLORE and WebDISCO, dMLCA requires multiple rounds of iterations, which requires automated data-sharing infrastructure, such as in the pSCANNER consortium^44^. It would be interesting to further develop few-shot algorithms for better communication-efficiency.

In our application of dMLCA algorithm to the EHR data cohort, we detected three significant subphenotypes of MIS-C. Specifically, subphenotype 1 (46.1%) represents patients with a mild presentation of MIS-C; subphenotype 2 (25.3%) represents children featuring a high risk of shock, cardiac and renal involvement, and a medium risk of respiratory symptoms; and subphenotype 3 (28.6%) represents patients requiring intensive care, with a high risk of shock and cardiac involvement, accompanied by a high risk of > 4 organ system involvement. Notably, we observed significant differences in the prevalence of these subphenotypes across the hospitals, which likely reflects the varying patient populations. To ensure the validity of our study cohort selection criteria, we further confirmed that the detected MIS-C subphenotypes were distinct from the general COVID-19 subphenotypes. However, we acknowledge that our study has limitations due to a moderate sample size. Therefore, further investigations on the MIS-C subphenotypes with a larger sample size at external pediatric cohorts are needed for confirmatory results.

In summary, our dMLCA algorithm is among the first efforts as unsupervised federated learning algorithms in the real-world data settings. This algorithm enables multiple sites to collaboratively learn disease subphenotypes based on aggregated data only. Moreover, it accommodates site-specific parameters to deal with heterogeneity across different sites. We believe this is a critical advance of federated learning in the disease subphenotyping using multi-site data in real-world settings.

## Data Availability

The data is not publicly available due to privacy concerns. The individual de-identified participant data will not be shared. The data that support the findings of this study may be available through request and DUA process to the corresponding authors.

## Acknowledgments

This study is part of the NIH RECOVER Initiative, which seeks to understand, treat, and prevent the post-acute sequelae of SARS-CoV-2 infection (PASC). For more information on RECOVER, visit https://recovercovid.org/. This research was funded by the National Institutes of Health (NIH) Agreement OT2HL161847-01 as part of the Researching COVID to Enhance Recovery (RECOVER) program of research. This work was supported partially through a Patient-Centered Outcomes Research Institute (PCORI) Project Program Award (ME-2019C3-18315, NJ, XL, QW and YC). We want to acknowledge Drs. Jasmin Divers and Lorna Thorpe’s thoughtful comments, which have greatly improved the presentation of this paper.

## Author Contributions

NJ, XL, and YC designed the model and the distributed algorithm. NJ, XL, QW, SR, CF, and YC devised the project. MM, JS, VL, HR, RW, and CB coordinated the data harmonization. NJ conducted the simulation experiments. NJ, XL, CF, SR, A.M., and YC designed the real-data analysis, and NJ performed the real-data analysis. NJ, XL, and YC drafted the manuscript. A.M., SR and CF provided clinical interpretations of the clinical findings. All coauthors provided critical edits of the early draft and approved the final version of the manuscript.

