## Supplemental Material for "Development and validation of a federated learning framework for detection of subphenotypes of multisystem inflammatory syndrome in children"

**Supplementary material of the manuscript "Development and validation of a federated learning framework for detection of subphenotypes of multisystem inflammatory syndrome in children"**

### Table of Contents

***Figure A1 Prevalence of manifest variables for latent class 2 (Panel A) and latent class 3 (Panel B) and the mixing proportion (prevalence) of latent class 2 (Panel A) and latent class 3 (Panel B) estimated by dMLCA algorithm and the pooled data in the MIS-C data analysis.......2***

***Figure A2 Attrition table for cohort selection........................................................................3***

***Figure A3. Results of MIS-C data analysis using dMLCA with four latent classes. A) Heatmap showing the prevalence of ten manifest variables in the four latent classes; Each column represents a latent class, and each row represents a variable. The color of the boxes represents the prevalence. The legend on the top right shows the scale of the colors. Red represents prevalence close to 100% and blue represents prevalence close to 0%. B) Prevalence of the four latent classes, overall and by site. ......................................................4***

**Figure A1 Prevalence of manifest variables for latent class 2 (Panel A) and latent class 3 (Panel B) and the mixing proportion (prevalence) of latent class 2 (Panel A) and latent class 3 (Panel B) estimated by dMLCA algorithm and the pooled data in the MIS-C data analysis**

A

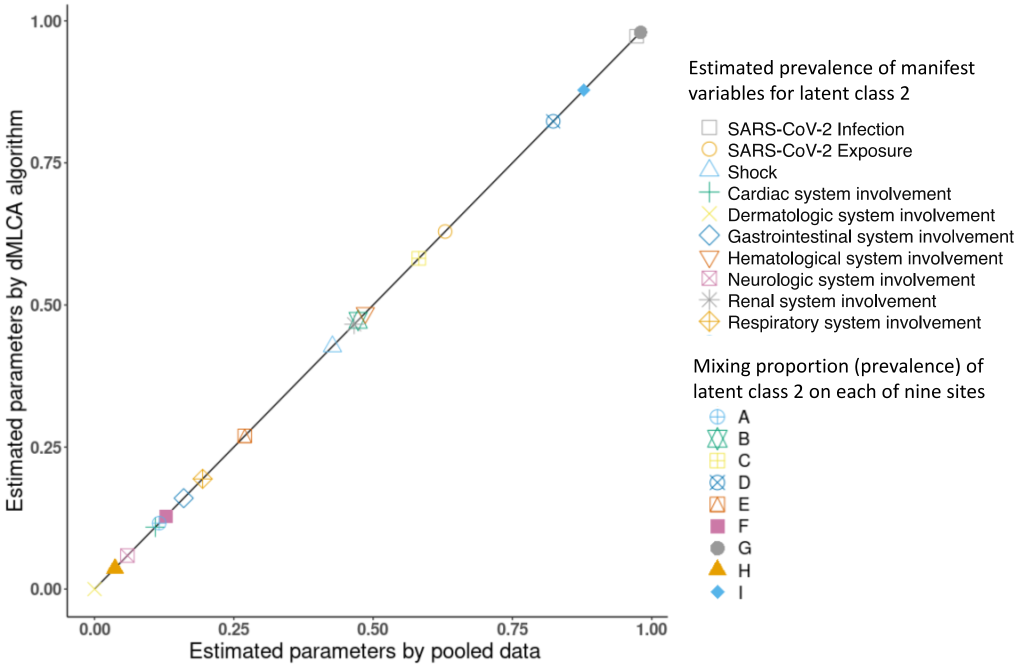

B

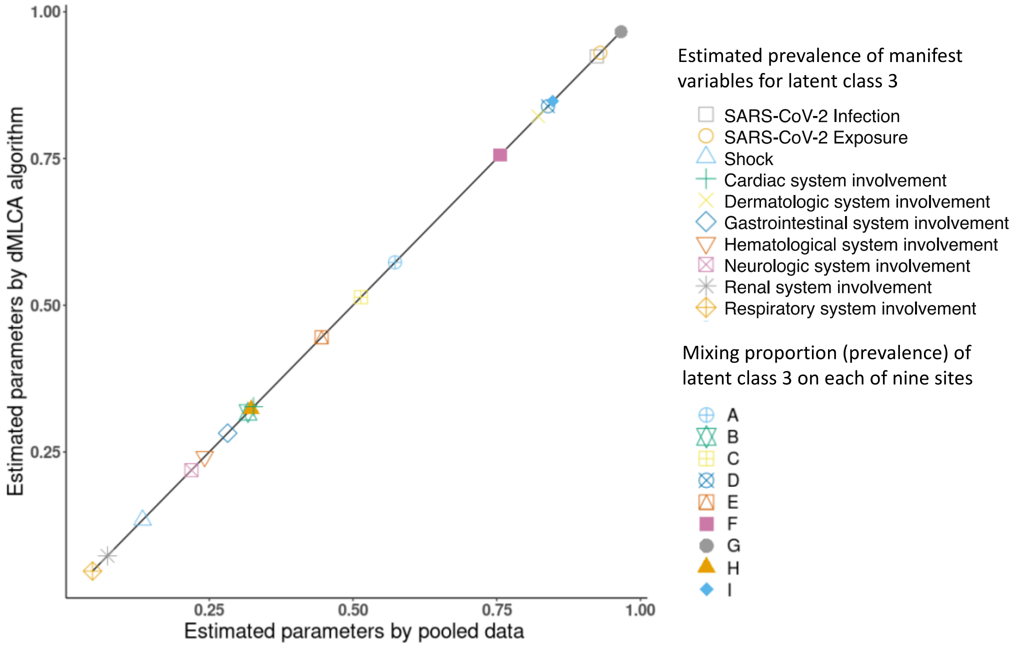

**Figure A2 Attrition table for cohort selection**

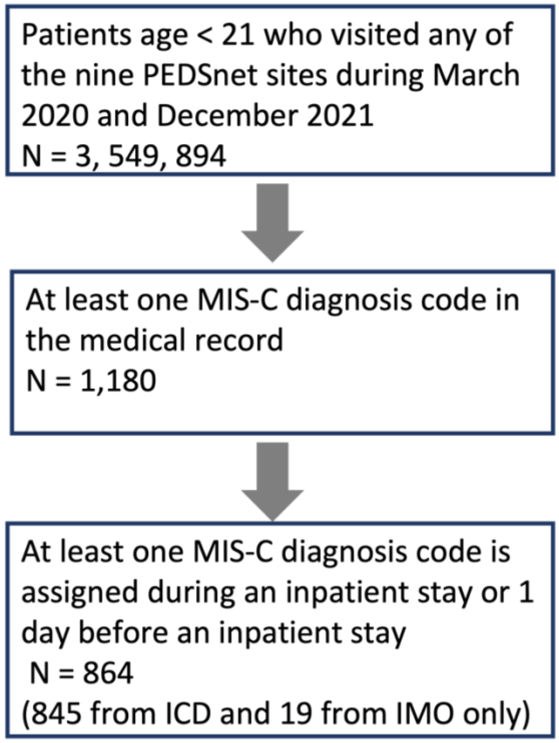

**Figure A3. Results of MIS-C data analysis using dMLCA with four latent classes. A) Heatmap showing the prevalence of ten manifest variables in the four latent classes; Each column represents a latent class, and each row represents a variable. The color of the boxes represents the prevalence. The legend on the top right shows the scale of the colors. Red represents prevalence close to 100% and blue represents prevalence close to 0%. B) Prevalence of the four latent classes, overall and by site.**

A

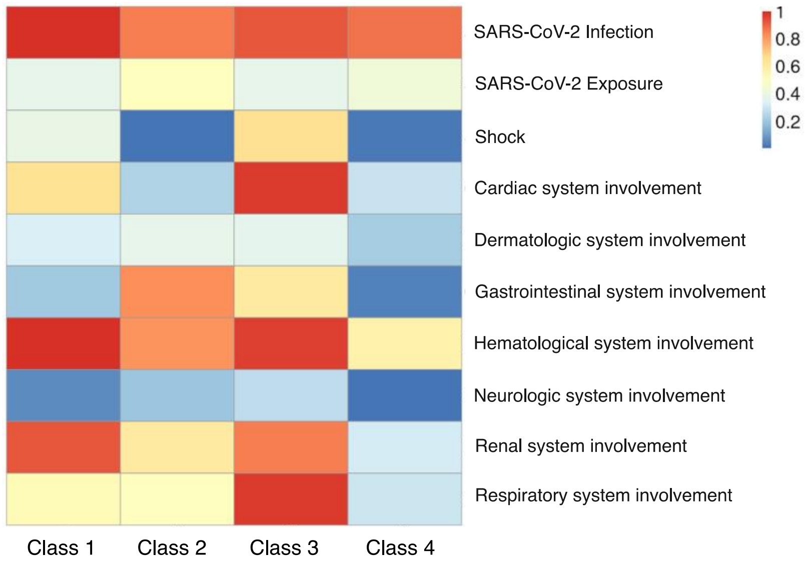

B

| | Class 1 | Class 2 | Class 3 | Class 4 |
|---|---|---|---|---|
| **Overall** | 28.6% | 18.9% | 25.9% | 26.5% |
| **A** | 56.3% | 4.0% | 8.6% | 31.1% |
| **B** | 28.6% | 24.1% | 23.0% | 24.3% |
| **C** | <1% | 39.9% | 58.0% | 2.1% |
| **D** | 29.9% | 17.8% | 21.4% | 30.9% |
| **E** | 23.2% | 4.1% | 44.1% | 28.6% |
| **F** | <1% | 15.3% | 26.8% | 57.9% |
| **G** | 50.9% | 12.3% | 12.3% | 24.5% |
| **H** | 9.9% | 30.8% | 12.7% | 46.6% |
| **I** | <1% | 32.4% | 59.3% | 8.3% |

**Table A1 Prevalence of ten variables across sites**

| Variable | Mean (range) |
|---|---|
| **SARS-CoV-2 Infection** | 89.5% (71.4%-99.1%) |
| **SARS-CoV-2 Exposure** | 38.9% (0.0%-67.1%) |
| **Shock** | 29.0% (12.5%-44.9%) |
| **Cardiac System Involvement** | 58.6% (40.7%-76.7%) |
| **Dermatologic System Involvement** | 31.6% (16.9%-42.9%) |
| **Gastrointestinal System Involvement** | 40.8% (23.9%-87.8%) |
| **Hematological System Involvement** | 82.8% (75.0%-90.5%) |
| **Neurologic System Involvement** | 13.5% (3.1%-36.7%) |
| **Renal System Involvement** | 64.2% (40.6%-78.8%) |
| **Respiratory System Involvement** | 59.2% (39.8%-85.7%) |

#### Table A2 Definition of the variables used in subphenotyping

| Variable | Categories | Definition |
|---|---|---|
| **SARS-CoV-2 Infection** | Yes/No | • Yes, if the following are positive<br> ○ SARS-CoV-2 infection 42 days before MIS-C or during hospitalization<br>• Else, no |
| **SARS-CoV-2 Exposure** | Yes/No | • Yes, if the following are positive<br> ○ SARS-CoV-2 exposure 42 days before MIS-C or during hospitalization<br>• Else, no |
| **Shock** | Yes/No | • Yes, if the following are positive<br> ○ Shock<br>• Else, no. |
| **Cardiac system involvement** | Yes/No | • Yes, if any of the following are positive<br> ○ coronary artery dilatation or aneurysm<br> ○ Heart failure (same as congestive heart failure)<br> ○ Myocarditis<br> ○ Arrhythmias<br> ○ Hypotension<br> ○ Pericarditis<br> ○ Cardiomyopathy<br> ○ Serum/plasma Troponin > 0.1 ng/mL<br>• Else, no. |
| **Dermatologic system involvement** | Yes/No | • Yes, if any of the following are positive<br> ○ Skin rashes<br> ○ Skin signs/symptoms<br> ○ Conjunctivitis<br>• Else, no |
| **Gastrointestinal system involvement** | Yes/No | • Yes, if any of the following are positive<br> ○ Abdominal pain<br> ○ Nausea and vomiting<br> ○ Diarrhea<br> ○ Intestinal obstruction/ileus<br> ○ Gastroenteritis<br>• Else, no |
| **Hematological system involvement** | Yes/No | • Yes, if any of the following are positive at any time during hospitalization<br> ○ Platelets < 150,000/microliter<br> ○ Plasma/serum D-dimer > 2 md/dL<br> ○ Lymphocytes < 1,000/microliter<br> ○ Thrombophlebitis and thromboembolism<br> ○ Aplastic anemia<br>• Else, no |
| **Neurologic system involvement** | Yes/No | • Yes, if any of the following are positive<br> ○ Headache<br> ○ Cognitive signs and symptoms<br> ○ Delirium<br> ○ Encephalopathy<br> ○ Nervous system signs and symptoms<br>• Else, no |
| **Renal system involvement** | Yes/No | • Yes, if any of the following are positive at any time during hospitalization<br> ○ Acute kidney injury |

| | | |
|---|---|---|
| | | ○ Fluid and electrolyte disturbance<br>○ Creatinine, age < 3yr, > 0.7 mg/dL OR age ≥ 3, > 1.0 mg/dL<br>○ Dialysis<br>• Else, no |
| **Respiratory system involvement** | Yes/No | • Yes, if any of the following are positive<br>○ Pneumonia<br>○ Bronchiolitis<br>○ Bronchitis<br>○ Cough<br>○ Cardiorespiratory signs and symptoms<br>○ Chest pain<br>○ Acute respiratory distress syndrome<br>○ Respiratory failure<br>○ Pleurisy pleural effusion and pulmonary collapse<br>○ Mechanical ventilation, either non-invasive or invasive<br>• Else, no |

**Table A3 AIC, BIC and adjusted BIC of dMLCA with two to six classes. * labels the best value.**

| No. of latent classes | 2 | 3 | 4 | 5 | 6 |
|---|---|---|---|---|---|
| AIC | 9305.14 | 9201.82 | 9125.45 | 9072.98 | 9030.04* |
| BIC | 9443.23 | 9430.38* | 9444.48 | 9482.47 | 9530.00 |
| Adjusted BIC | 9351.13 | 9277.94 | 9231.70 | 9209.36 | 9196.55* |

### Method

We present the proposed method in this section. We consider a multi-site study involving $K$ sites with a sample of $n_k$ subjects on site $k$. The total sample size is $N = \sum_{k=1}^{K} n_k$. Suppose the study population of a disease is a mixture of $C$ unobserved subpopulations – or, equivalently, subphenotypes of the disease - also referred to as latent classes. For the $i$-th subject on the $k$-th site, where $i = 1, 2, \dots, n_k$ and $k = 1,2, \dots, K$, we observe $q$ binary manifest variables (also known as outcomes) of interest $Y_{ki} = \left(Y_{ki1}, Y_{ki2}, \dots, Y_{kiq}\right)^\top \in \{0, 1\}^q$, and we denote $Z_{ki} \in \{1,2, \dots, C\}$ as the unobserved latent class membership indicator. Note that the models for categorical (nominal or ordinal) and count data can be formulated similarly with modifications in the distribution of $Y_{ki}$. Further, we collect $p$ observed covariates $X_{ki} = \left(X_{ki1}, X_{ki2}, \dots, X_{kip}\right)^\top \in R^p$. by assuming conditional independence among different outcomes of a subject $Y_{kij}, j = 1,2, \dots, q$ and that $X_{ki}$ affects $Y_{ki}$ only through the class membership given the latent class membership $Z_{ki}$, the distribution of $Y_{ki}$ is

$$f(Y_{ki}|X_{ki}) = \sum_{c=1}^{C} Pr(Z_{ki} = c|X_{ki}) f(Y_{ki}, \pi_c) = \sum_{c=1}^{C} \lambda_{kc}(X_{ki}) \prod_{j=1}^{q} f\left(Y_{kij}, \pi_{cj}\right),$$

where $\pi_{cj} = P(Y_{kij}|Z_{ki} = c)$ and $\lambda_{kc}(X_{ki}) = Pr(Z_{ki} = c|X_{ki}),\ c = 1, \dots, C,$ is the probability of the subjects on the $k$-th site belonging to class $c$, i.e., class membership probability of the subject [**Error! Reference source not found.**]. To formulate $\lambda_{kc}(X_{ki})$, we use the polytomous logistic regression model with site-specific intercepts to account for the heterogeneity in the populations across sites as shown in **Figure 1**. Specifically, by using class $C$ as the reference, we have

$$\lambda_{kc}(X_{ki}) = \frac{\exp\{\alpha_{kc} + \beta_c^\top X_{ki}\}}{1 + \sum_{j=1}^{C-1} \exp\{\alpha_{kj} + \beta_j^\top X_{ki}\}}, \qquad \forall c \in \{1, \dots, C-1\}$$

$$\lambda_{kC}(X_{ki}) = \frac{1}{1 + \sum_{j=1}^{C-1} \exp\{\alpha_{kj} + \beta_j^\top X_{ki}\}}.$$

Here $\alpha_{kc} \in R, k = 1, \dots, K, c = 1, \dots, C-1$ are intercepts that are different across sites and classes, and $\beta_c \in R^p, c = 1,2, \dots, C-1$, are class-specific covariate coefficients that are shared across sites to imply a common effect of covariates on the class membership probabilities. We write $\theta = (\pi^\top, \alpha^\top, \beta^\top)^\top$ to denote all parameters.

#### Algorithm

In this section, we first describe the algorithm when patient-level data sharing is allowed. Then, we show how to perform EM algorithm with no patient-level data. We treat the unobserved class membership $Z_{ki}$'s as missing data and use the expectation-maximization (EM) algorithm to do estimation. Let $\theta^t$ be the estimator of all the parameters at the $t$-th iteration, to update, $\theta^{t+1}$ is the solution to the equation

$$\frac{\partial}{\partial\theta} Q(\theta|\theta^t) = 0, \tag{1}$$

where $Q(\theta|\theta^t)$ is the conditional expectation of the complete loglikelihood given $\theta^t$, $Y$, and $X$. Let $w_{kic}^t = \Pr(Z_{ki} = c|Y_{ki}, X_{ki}, \theta^t)$, then solving (1) gives

$$\pi_{cj}^{t+1} = \frac{\sum_{k=1}^K \sum_{i=1}^{n_k} w_{kic}^t Y_{kij}}{\sum_{k=1}^K \sum_{i=1}^{n_k} w_{kic}^t}. \tag{2}$$

Since there is no closed-form solution for $\alpha^{t+1}$ and $\beta^{t+1}$, we use Newton-Raphson method to solve for $\alpha^{t+1}$ and $\beta^{t+1}$. Let $\Gamma = (\alpha^\top, \beta^\top)^\top$, at $t'$-th iteration of the Newton-Raphson method, we update $\Gamma^{t'}$ by

$$\Gamma^{t'+1} = \Gamma^{t'} - \left\{\frac{\partial^2}{\partial\Gamma\,\partial\Gamma^\top} Q\left(\theta\middle|\pi^t, \Gamma^{t'}\right)\right\}^{-1} \frac{\partial}{\partial\Gamma} Q(\theta|\pi^t, \Gamma^{t'}) \tag{3}$$

until convergence or the maximum number of iterations reached to get $\Gamma^{t+1}$. Note that this is a separate iterative procedure inside each iteration of the EM algorithm. We refer to the iterations in Newton-Raphson method as the 'inner loop' and that in EM algorithm the 'outer loop'. Alternatively, one can update $\alpha^{t+1}$ and $\beta^{t+1}$ using one Newton-Raphson's step to save the inner loop of iterations

$$\Gamma^{t+1} = \Gamma^t - \left\{\frac{\partial^2}{\partial\Gamma\,\partial\Gamma^\top} Q(\theta|\theta^t)\right\}^{-1} \frac{\partial}{\partial\Gamma} Q(\theta|\theta^t). \tag{4}$$

Thanks to the decomposability of the updating formulas (2), (3) and (4) with respect to sites, we can calculate the separated parts in each site and then combine the results in the leading site to perform the updating steps. Note that the information shared among sites consists of only aggregated results and no raw data. This method does not lose any information compared with a pooled estimator because the updating formulas (2) to (4) are all accurately computed.

When update $\alpha^{t+1}$ and $\beta^{t+1}$ in a distributed setting, we use the approach with one Newton-Raphson's step and call it as dMLCA algorithm to reduce communication cost. The one with multiple Newton-Raphson's steps involves more communication rounds among sites caused by inner loop iteration and is called dMLCA-EM. More comparisons between these two approaches and discussions are given in the Result section. Without loss of generality, we treat site 1 as the leading site. Our proposed distributed algorithm dMLCA is summarized below.

**Algorithm 1 (dMLCA)**

**Input**: $Y, X, \theta^0, t = 0$;

1. In site 1 (leading site),
   Randomly generated initial values $\pi^0$ and $\beta^0$ and broadcast.
2. In site 1 to $K$,
   Generate $\alpha_{kc}^0$'s randomly.

**While** the stopping criterion is not met **do**

3. For site $k = 1$ to $K$,
   Compute $\sum_{i=1}^{n_k} w_{kic}^t$ and $\sum_{i=1}^{n_k} w_{kic}^t Y_{kij}$ for each $c = 1, \dots, C$ and $j = 1, \dots, q$, and transfer the results to site 1.
   Compute $\frac{\partial}{\partial\Gamma} Q^k(\theta|\theta^t)|_{\theta=\theta^t}$ and $\frac{\partial^2}{\partial\Gamma\,\partial\Gamma^\top} Q^k(\theta|\theta^t)|_{\theta=\theta^t}$ for the local objective function $Q^k(\theta|\theta^t)$, and transfer the results to site 1.

4. In site 1, update each parameter by

   $\pi_{cj}^{t+1} \leftarrow \frac{\sum_{k=1}^K \sum_{i=1}^{n_k} w_{kic}^t Y_{kij}}{\sum_{k=1}^K \sum_{i=1}^{n_k} w_{kic}^t}$, for each $c = 1, \dots, C$ and $j = 1, \dots, q$

   $S(\Gamma^t) \leftarrow \sum_{k=1}^K \frac{\partial}{\partial\Gamma} Q^k(\theta|\theta^t)|_{\theta=\theta^t}$ ; $H(\Gamma^t) \leftarrow \sum_{k=1}^K \frac{\partial^2}{\partial\Gamma\,\partial\Gamma^\top} Q^k(\theta|\theta^t)|_{\theta=\theta^t}$

   $\Gamma^{t+1} \leftarrow \Gamma^t - H(\Gamma^t)^{-1} S(\Gamma^t)$

5. In site 1, broadcast the updated parameters $\theta^{t+1}$.
6. $t = t + 1$

**End while**

7. Obtain $\hat{\theta} \leftarrow \theta^t$.

**Output**: $\hat{\theta}$.

Note that the initial values can simply be chosen arbitrarily. To guarantee finding the global maximum, multiple initial values are needed to obtain multiple estimators and the one with the largest likelihood is selected as the final estimator. A good candidate of initial values can be the local estimator, however even when using the local estimator as the initial value, multiple attempts of other initial values are still needed. This multiple initialization procedure does not necessarily increase the communication rounds and only requires transferring more digits of data at each round.

**Simulation**

**Design**

We evaluate the performance of the proposed dMLCA algorithm on simulated data. We apply dMLCA under 5 settings and compare it with three methods: 1. dMLCA-EM algorithm; 2. the local estimator using only the data on the leading site; 3. LCA model on the centralized data treating the data as in a single-site analysis. The comparison with the local estimator is commonly seen in distributed methodology papers to study the information gained from multi-site study [1, 2]. The comparison with LCA is to show that this commonly applied strategy can be problematic with the presence of between-site heterogeneity. The performance of the estimators is evaluated by the estimation error in $\pi$, $\frac{1}{Cq}\sum_{c=1}^{C}\sum_{j=1}^{q}\left(\hat{\pi}_{cj} - \pi_{cj}^{*}\right)^2$ , and in the class membership probabilities $\lambda$, $\frac{1}{KCn}\sum_{k=1}^{K}\sum_{c=1}^{C}\sum_{i=1}^{n_k}\left\{\hat{\lambda}_{kc}(X_{ki}) - \lambda_{kc}^{*}(X_{ki})\right\}^2$. Besides, we compare the total number of iterations in dMLCA and dMLCA-EM algorithm to investigate their communication costs.

In our simulation settings, we consider $C = 3$ latent classes, $q = 5$ binary outcomes, and $p = 3$ subject-specific covariates. We set one covariate $X_{ki1}$ to be binary following a Bernoulli distribution with mean 0.5, and the other two covariates are continuous following normal distributions $X_{ki2} \sim N(0, 1)$ and $X_{ki3} \sim N(0, 3)$. We fix the regression parameters $\beta_1 = (0.5, 1, 0.5)^{\top}$ and $\beta_2 = (1, 0, -1)^{\top}$. We set the baseline class membership probabilities $\lambda_{kc}(X_{ki} = 0)$ as in **Table A4** when the number of sites $K = 5$, and when $K = 10$, we set the same values for the extra five sites.

| $\lambda_{kc}(0)$ | Class 1<br>$c = 1$ | Class 2<br>$c = 2$ | Class 3<br>$c = 3$ |
|---|---|---|---|
| Site 1 ($k = 1$) | 0.15 | 0.20 | 0.65 |
| Site 2 ($k = 2$) | 0.34 | 0.33 | 0.33 |
| Site 3 ($k = 3$) | 0.50 | 0.25 | 0.25 |
| Site 4 ($k = 4$) | 0.65 | 0.20 | 0.15 |
| Site 5 ($k = 5$) | 0.10 | 0.15 | 0.75 |

**Table A4 Baseline class membership probabilities $\lambda_{kc}(0)$ in the simulation study.**

In Setting 1 – 3, we let the variation of the outcomes across classes be large by shifting the mean of outcomes in the three classes among (0.1, 0.5, 0.9) and set $\pi = \pi_1$ as in **Table A5**. Fixing $\pi$ at $\pi_1$, we vary the number of sites ($K =$

$5, 10$) and sample size in each site ($n = 500, 1000$) to study how the sample size influences the parameter estimates. We also tried settings where the sample sizes are unequal across sites, which leads to the same conclusion with the equal sample size setting and therefore the results are not given in this paper. In Setting 4 – 5, we fix $K = 10$ and $n = 1000$, and reduce the variation among classes to see the performance of competing estimators when the similarity among classes is increased. In Setting 4, we set $\pi = \pi_2$ where $\pi_2$ is constructed by substituting 0.1, 0.5, 0.9 in $\pi_1$ by 0.2, 0.5, 0.8, respectively. Similarly, in Setting 5, $\pi = \pi_3$ where $\pi_3$ is constructed by substituting 0.1, 0.5, 0.9 in $\pi_1$ by 0.3, 0.5, 0.7, respectively.

| $\pi_{1,cj}$ | Class 1<br>$c = 1$ | Class 2<br>$c = 2$ | Class 3<br>$c = 3$ |
|---|---|---|---|
| Outcome (j= 1) | 0.1 | 0.9 | 0.5 |
| Outcome (j= 2) | 0.5 | 0.1 | 0.9 |
| Outcome (j= 3) | 0.9 | 0.5 | 0.1 |
| Outcome (j= 4) | 0.1 | 0.9 | 0.5 |
| Outcome (j= 5) | 0.5 | 0.1 | 0.9 |

**Table A5 Means of the outcomes $\pi_1$ in Setting 1-3.**

After generating the data $(Y, X)$ using the above settings, we estimate the parameters using the four candidate methods (dMLCA, dMLCA-EM, local, LCA) and calculate the estimation errors. We repeat the procedure for 200 times under each setting and inspect the overall performance of the four candidate methods.

**Results**

**Figure A4** shows the box plot of the 200 estimation errors under the Setting 1, 2, and 3, with varying numbers of sites and patients, and **Figure A5** presents the results from Setting 3, 4, and 5, with varying similarity level among latent classes. In all the settings, both versions of dMLCA have smaller estimation errors and smaller variances than the other two candidates, indicating the benefits of dMLCA gained from borrowing information from all the sites to improve the modeling efficiency and the importance of accounting for heterogeneity.

dMLCA outperforms the local estimator. As the sample size in each site (n) or the number of sites (K) increases, the estimation errors and their variance decrease (**Panel A of Figure A4**), indicating that involving more sites and patients in the study can improve the accuracy. The local estimator is not shown in **Figure A5** because it becomes unstable when the similarity among the classes becomes large. In contrast, both versions of dMLCA estimators perform satisfactorily. With the same sample sizes, both the estimation errors and their variances increase as the similarity among the subpopulations increases. This indicates that for subphenotypes with larger similarities, more information is required to reach the same accuracy as the estimation with very differential subpopulations, and therefore multi-site studies are desirable in this situation.

As shown in **Panel A** of **Figure A4**, dMLCA and LCA on centralized data have similar performance in estimating $\pi$. However, **Panel B** of **Figure A4** shows that LCA performs the worst among all the methods with the largest bias in terms of class membership probability estimation, demonstrating that LCA is not robust to the heterogeneity in the population across sites in terms of class membership probability estimation. The same pattern is shown in **Panel B of Figure A5**. By accounting for site-specific class membership probabilities, dMLCA properly handles the heterogeneous population across sites and provides reliable results.

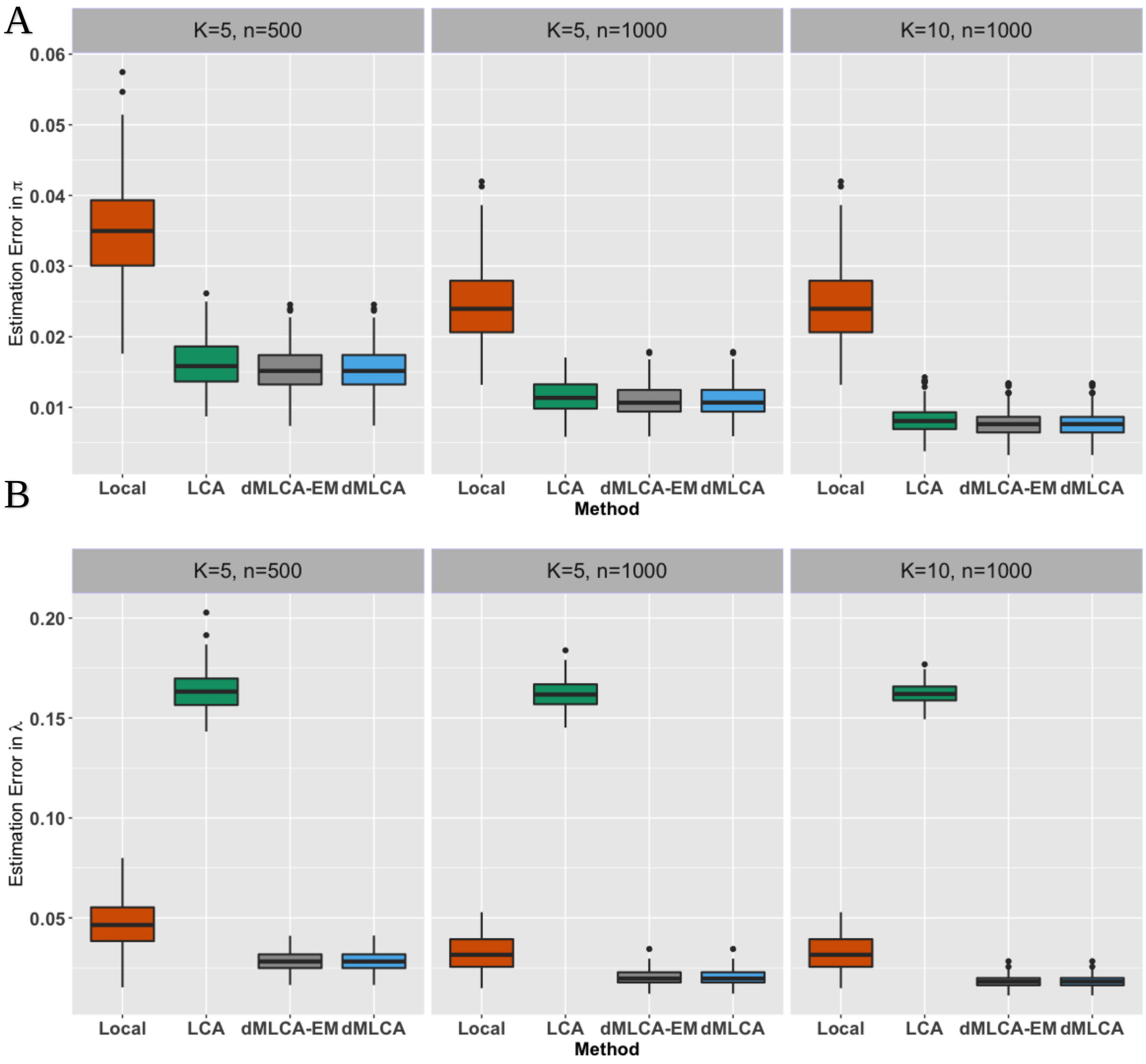

**Figure A4 Estimation errors in manifest variable prevalence $\hat{\pi}$ (Panel A) and estimation errors in class membership probabilities $\hat{\lambda}$ (Panel B) under Setting 1, 2, and 3 over 200 repetitions for the local estimator(red), LCA on pooled data (green), dMLCA-EM (grey), and dMLCA (blue) with various levels of sites $K$ and the sample size in each site $n$.**

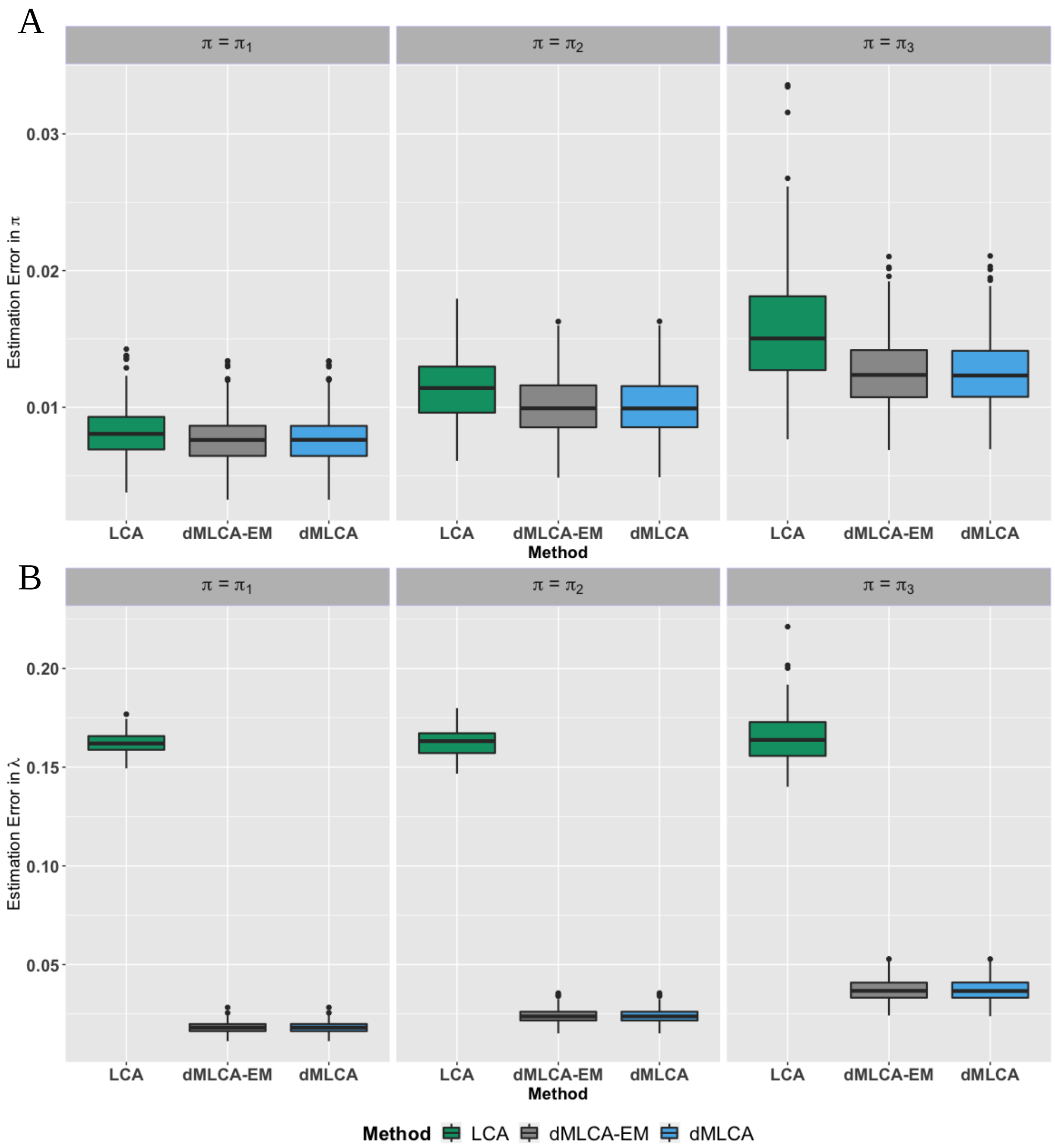

**Figure A5 Estimation errors in manifest variable prevalence $\hat{\pi}$ (Panel A) and estimation errors in class membership probabilities $\hat{\lambda}$ (Penal B) under Setting 3, 4, and 5 over 200 repetitions for LCA on pooled data (green), dMLCA-EM (grey), and dMLCA (blue) with different levels of subphenotype similarity. Specifically, $\pi_1, \pi_2, \pi_3$ have increasing similarities among subtypes and thus have increasing difficulty for accurate estimates. The values of $\pi_1, \pi_2, \pi_3$ are given in Table A4.**

The total number of communication rounds needed to converge for dMLCA and dMLCA-EM under our Setting 3 is displayed in **Figure A6**. Note that dMLCA-EM involves two layers of iterations, and the number of its inner loop iterations can be high. The inner loop within dMLCA-EM also requires communication between sites, therefore, dMLCA requires much fewer communication rounds (median 25) compared with dMLCA-EM (median 220) and is more favorable based on our simulation studies.

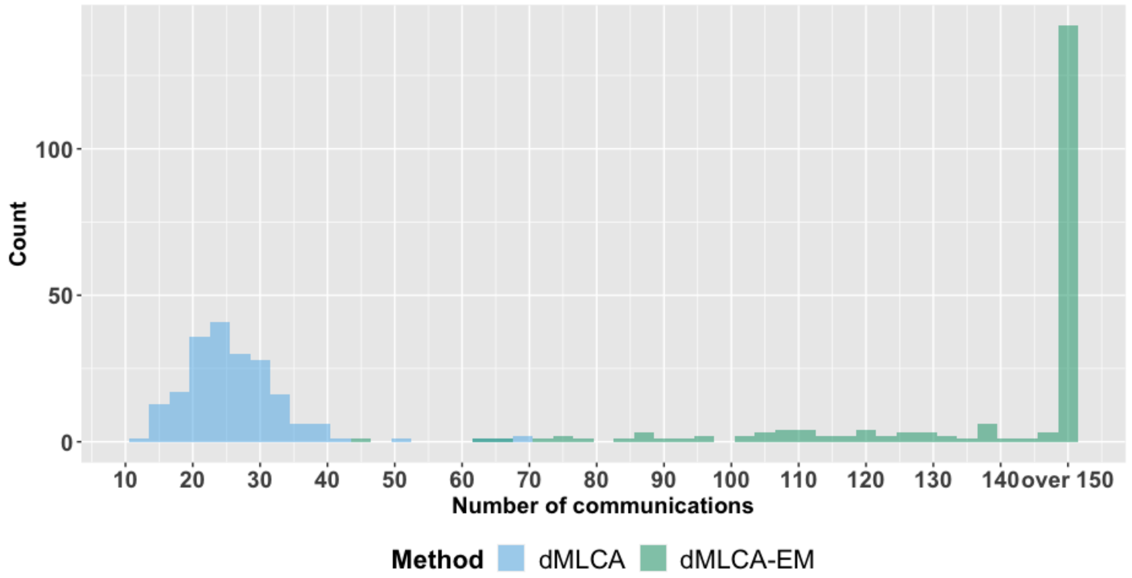

**Figure A6 Number of communications between each site and the leading site over 200 repetitions for the dMLCA (blue) and dMLCA-EM (green), under Setting 3 ($K = 10, n = 1000, \pi = \pi_1$).**

### Table A4 Code set for variables used in the analysis

| | Variable | Concept name | Concept code | Vocabulary | System |
|---|---|---|---|---|---|
| **1** | aortic; peripheral; and visceral artery aneurysms | Aneurysm of carotid artery | I72.0 | ICD10CM | cardiac |
| **2** | aortic; peripheral; and visceral artery aneurysms | Aneurysm of vertebral artery | I72.6 | ICD10CM | cardiac |
| **3** | aortic; peripheral; and visceral artery aneurysms | Aneurysm of other specified arteries | I72.8 | ICD10CM | cardiac |
| **4** | aortic; peripheral; and visceral artery aneurysms | Abdominal aortic aneurysm, ruptured | I71.3 | ICD10CM | cardiac |
| **5** | aortic; peripheral; and visceral artery aneurysms | Dressler's syndrome | I24.1 | ICD10CM | cardiac |
| **6** | aortic; peripheral; and visceral artery aneurysms | Thoracic aortic aneurysm, without rupture | I71.2 | ICD10CM | cardiac |
| **7** | aortic; peripheral; and visceral artery aneurysms | Syphilitic aneurysm of aorta | A52.01 | ICD10CM | cardiac |
| **8** | aortic; peripheral; and visceral artery aneurysms | Aortic aneurysm of unspecified site, without rupture | I71.9 | ICD10CM | cardiac |
| **9** | aortic; peripheral; and visceral artery aneurysms | Aneurysm of unspecified site | I72.9 | ICD10CM | cardiac |
| **10** | aortic; peripheral; and visceral artery aneurysms | Aneurysm of iliac artery | I72.3 | ICD10CM | cardiac |
| **11** | aortic; peripheral; and visceral artery aneurysms | Aneurysm of heart | I25.3 | ICD10CM | cardiac |
| **12** | aortic; peripheral; and visceral artery aneurysms | Aneurysm of aorta in diseases classified elsewhere | I79.0 | ICD10CM | cardiac |
| **13** | aortic; peripheral; and visceral artery aneurysms | Aneurysm of artery of upper extremity | I72.1 | ICD10CM | cardiac |
| **14** | aortic; peripheral; and visceral artery aneurysms | Aneurysm of artery of lower extremity | I72.4 | ICD10CM | cardiac |
| **15** | aortic; peripheral; and visceral artery aneurysms | Aortic aneurysm of unspecified site, ruptured | I71.8 | ICD10CM | cardiac |
| **16** | aortic; peripheral; and visceral artery aneurysms | Abdominal aortic aneurysm, without rupture | I71.4 | ICD10CM | cardiac |
| **17** | aortic; peripheral; and visceral artery aneurysms | Aneurysm of other precerebral arteries | I72.5 | ICD10CM | cardiac |
| **18** | aortic; peripheral; and visceral artery aneurysms | Thoracic aortic aneurysm, ruptured | I71.1 | ICD10CM | cardiac |
| **19** | aortic; peripheral; and visceral artery aneurysms | Aneurysm of renal artery | I72.2 | ICD10CM | cardiac |
| **20** | aortic; peripheral; and visceral artery aneurysms | Thoracoabdominal aortic aneurysm, without rupture | I71.6 | ICD10CM | cardiac |
| **21** | aortic; peripheral; and visceral artery aneurysms | Thoracoabdominal aortic aneurysm, ruptured | I71.5 | ICD10CM | cardiac |
| **22** | arrhythmias | Bifascicular block | I45.2 | ICD10CM | cardiac |
| **23** | arrhythmias | Other cardiac arrhythmias | I49 | ICD10CM | cardiac |
| **24** | arrhythmias | Paroxysmal tachycardia | I47 | ICD10CM | cardiac |
| **25** | arrhythmias | Right fascicular block | I45.0 | ICD10CM | cardiac |

| | | | | | |
|---|---|---|---|---|---|
| **26** | arrhythmias | Chronic atrial fibrillation, unspecified | I48.20 | ICD10CM | cardiac |
| **27** | arrhythmias | Ventricular flutter | I49.02 | ICD10CM | cardiac |
| **28** | arrhythmias | Unspecified atrial fibrillation and atrial flutter | I48.9 | ICD10CM | cardiac |
| **29** | arrhythmias | Paroxysmal atrial fibrillation | I48.0 | ICD10CM | cardiac |
| **30** | arrhythmias | Atrioventricular and left bundle-branch block | I44 | ICD10CM | cardiac |
| **31** | arrhythmias | Unspecified premature depolarization | I49.40 | ICD10CM | cardiac |
| **32** | arrhythmias | Other and unspecified fascicular block | I44.6 | ICD10CM | cardiac |
| **33** | arrhythmias | Unspecified right bundle-branch block | I45.10 | ICD10CM | cardiac |
| **34** | arrhythmias | Persistent atrial fibrillation | I48.1 | ICD10CM | cardiac |
| **35** | arrhythmias | Other specified heart block | I45.5 | ICD10CM | cardiac |
| **36** | arrhythmias | Other and unspecified premature depolarization | I49.4 | ICD10CM | cardiac |
| **37** | arrhythmias | Other specified conduction disorders | I45.89 | ICD10CM | cardiac |
| **38** | arrhythmias | Junctional premature depolarization | I49.2 | ICD10CM | cardiac |
| **39** | arrhythmias | Atrioventricular block, complete | I44.2 | ICD10CM | cardiac |
| **40** | arrhythmias | Other right bundle-branch block | I45.19 | ICD10CM | cardiac |
| **41** | arrhythmias | Unspecified fascicular block | I44.60 | ICD10CM | cardiac |
| **42** | arrhythmias | Other conduction disorders | I45 | ICD10CM | cardiac |
| **43** | arrhythmias | Cardiac arrhythmia, unspecified | I49.9 | ICD10CM | cardiac |
| **44** | arrhythmias | Sick sinus syndrome | I49.5 | ICD10CM | cardiac |
| **45** | arrhythmias | Ventricular tachycardia | I47.2 | ICD10CM | cardiac |
| **46** | arrhythmias | Ventricular fibrillation and flutter | I49.0 | ICD10CM | cardiac |
| **47** | arrhythmias | Atrial premature depolarization | I49.1 | ICD10CM | cardiac |
| **48** | arrhythmias | Atypical atrial flutter | I48.4 | ICD10CM | cardiac |
| **49** | arrhythmias | Pre-excitation syndrome | I45.6 | ICD10CM | cardiac |
| **50** | arrhythmias | Unspecified atrioventricular block | I44.30 | ICD10CM | cardiac |
| **51** | arrhythmias | Atrioventricular block, first degree | I44.0 | ICD10CM | cardiac |
| **52** | arrhythmias | Cardiac arrest | I46 | ICD10CM | cardiac |
| **53** | arrhythmias | Supraventricular tachycardia | I47.1 | ICD10CM | cardiac |
| **54** | arrhythmias | Ventricular fibrillation | I49.01 | ICD10CM | cardiac |
| **55** | arrhythmias | Other specified cardiac arrhythmias | I49.8 | ICD10CM | cardiac |
| **56** | arrhythmias | Encounter for adjustment and management of other part of cardiac pacemaker | Z45.018 | ICD10CM | cardiac |
| **57** | arrhythmias | Presence of cardiac pacemaker | Z95.0 | ICD10CM | cardiac |
| **58** | arrhythmias | Other atrioventricular block | I44.39 | ICD10CM | cardiac |
| **59** | arrhythmias | Permanent atrial fibrillation | I48.21 | ICD10CM | cardiac |
| **60** | arrhythmias | Trifascicular block | I45.3 | ICD10CM | cardiac |
| **61** | arrhythmias | Ventricular premature depolarization | I49.3 | ICD10CM | cardiac |
| **62** | arrhythmias | Left bundle-branch block, unspecified | I44.7 | ICD10CM | cardiac |
| **63** | arrhythmias | Other fascicular block | I44.69 | ICD10CM | cardiac |
| **64** | arrhythmias | Long QT syndrome | I45.81 | ICD10CM | cardiac |
| **65** | arrhythmias | Atrioventricular block, second degree | I44.1 | ICD10CM | cardiac |

| | | | | | |
|---|---|---|---|---|---|
| **66** | arrhythmias | Left anterior fascicular block | I44.4 | ICD10CM | cardiac |
| **67** | arrhythmias | Encounter for checking and testing of cardiac pacemaker pulse generator [battery] | Z45.010 | ICD10CM | cardiac |
| **68** | arrhythmias | Other and unspecified right bundle-branch block | I45.1 | ICD10CM | cardiac |
| **69** | arrhythmias | Other and unspecified atrioventricular block | I44.3 | ICD10CM | cardiac |
| **70** | arrhythmias | Typical atrial flutter | I48.3 | ICD10CM | cardiac |
| **71** | arrhythmias | Nonspecific intraventricular block | I45.4 | ICD10CM | cardiac |
| **72** | arrhythmias | Other premature depolarization | I49.49 | ICD10CM | cardiac |
| **73** | arrhythmias | Chronic atrial fibrillation | I48.2 | ICD10CM | cardiac |
| **74** | arrhythmias | Other persistent atrial fibrillation | I48.19 | ICD10CM | cardiac |
| **75** | arrhythmias | Atrial fibrillation and flutter | I48 | ICD10CM | cardiac |
| **76** | arrhythmias | Left posterior fascicular block | I44.5 | ICD10CM | cardiac |
| **77** | arrhythmias | Conduction disorder, unspecified | I45.9 | ICD10CM | cardiac |
| **78** | arrhythmias | Longstanding persistent atrial fibrillation | I48.11 | ICD10CM | cardiac |
| **79** | arrhythmias | Other specified conduction disorders | I45.8 | ICD10CM | cardiac |
| **80** | arrhythmias | Unspecified atrial flutter | I48.92 | ICD10CM | cardiac |
| **81** | arrhythmias | Re-entry ventricular arrhythmia | I47.0 | ICD10CM | cardiac |
| **82** | arrhythmias | Paroxysmal tachycardia, unspecified | I47.9 | ICD10CM | cardiac |
| **83** | arrhythmias | Unspecified atrial fibrillation | I48.91 | ICD10CM | cardiac |
| **84** | cardiac | Cardiac murmurs and other cardiac sounds | R01 | ICD10CM | cardiac |
| **85** | cardiac | Unspecified abnormalities of heart beat | R00.9 | ICD10CM | cardiac |
| **86** | cardiac | Orthopnea | R06.01 | ICD10CM | cardiac |
| **87** | cardiac | Other cardiac sounds | R01.2 | ICD10CM | cardiac |
| **88** | cardiac | Tachycardia, unspecified | R00.0 | ICD10CM | cardiac |
| **89** | cardiac | Abnormal blood-pressure reading, without diagnosis | R03 | ICD10CM | cardiac |
| **90** | cardiac | Cardiac murmur, unspecified | R01.1 | ICD10CM | cardiac |
| **91** | cardiac | Elevated blood-pressure reading, without diagnosis of hypertension | R03.0 | ICD10CM | cardiac |
| **92** | cardiac | Abnormalities of heart beat | R00 | ICD10CM | cardiac |
| **93** | cardiac | Other abnormalities of heart beat | R00.8 | ICD10CM | cardiac |
| **94** | cardiac | Bradycardia, unspecified | R00.1 | ICD10CM | cardiac |
| **95** | cardiac | Nonspecific low blood-pressure reading | R03.1 | ICD10CM | cardiac |
| **96** | cardiac | Benign and innocent cardiac murmurs | R01.0 | ICD10CM | cardiac |
| **97** | cardiac | Other chest pain | R07.8 | ICD10CM | cardiac |
| **98** | cardiac | Palpitations | R00.2 | ICD10CM | cardiac |
| **99** | cardiomyopathy | Takotsubo syndrome | I51.81 | ICD10CM | cardiac |
| **100** | cardiomyopathy | Obstructive hypertrophic cardiomyopathy | I42.1 | ICD10CM | cardiac |
| **101** | cardiomyopathy | Other restrictive cardiomyopathy | I42.5 | ICD10CM | cardiac |
| **102** | cardiomyopathy | Cardiomyopathy, unspecified | I42.9 | ICD10CM | cardiac |
| **103** | cardiomyopathy | Diphtheritic cardiomyopathy | A36.81 | ICD10CM | cardiac |

| | | | | | |
|---|---|---|---|---|---|
| **104** | cardiomyopathy | Cardiomyopathy in diseases classified elsewhere | I43 | ICD10CM | cardiac |
| **105** | cardiomyopathy | Endomyocardial (eosinophilic) disease | I42.3 | ICD10CM | cardiac |
| **106** | cardiomyopathy | Endocardial fibroelastosis | I42.4 | ICD10CM | cardiac |
| **107** | cardiomyopathy | Cardiomyopathy due to drug and external agent | I42.7 | ICD10CM | cardiac |
| **108** | cardiomyopathy | Other cardiomyopathies | I42.8 | ICD10CM | cardiac |
| **109** | cardiomyopathy | Ischemic cardiomyopathy | I25.5 | ICD10CM | cardiac |
| **110** | cardiomyopathy | Cardiomyopathy | I42 | ICD10CM | cardiac |
| **111** | cardiomyopathy | Other hypertrophic cardiomyopathy | I42.2 | ICD10CM | cardiac |
| **112** | cardiomyopathy | Dilated cardiomyopathy | I42.0 | ICD10CM | cardiac |
| **113** | cardiomyopathy | Peripartum cardiomyopathy | O90.3 | ICD10CM | cardiac |
| **114** | cardiomyopathy | Viral cardiomyopathy | B33.24 | ICD10CM | cardiac |
| **115** | cardiomyopathy | Alcoholic cardiomyopathy | I42.6 | ICD10CM | cardiac |
| **116** | chest pain | Chest pain on breathing | R07.1 | ICD10CM | cardiac |
| **117** | chest pain | Intercostal pain | R07.82 | ICD10CM | cardiac |
| **118** | chest pain | Pleurodynia | R07.81 | ICD10CM | cardiac |
| **119** | chest pain | Precordial pain | R07.2 | ICD10CM | cardiac |
| **120** | chest pain | Chest pain, unspecified | R07.9 | ICD10CM | cardiac |
| **121** | chest pain | Other chest pain | R07.89 | ICD10CM | cardiac |
| **122** | heart failure | Cardiac failure due to anesthesia during pregnancy, first trimester | O29.121 | ICD10CM | cardiac |
| **123** | heart failure | Acute systolic (congestive) heart failure | I50.21 | ICD10CM | cardiac |
| **124** | heart failure | Acute diastolic (congestive) heart failure | I50.31 | ICD10CM | cardiac |
| **125** | heart failure | Heart failure, unspecified | I50.9 | ICD10CM | cardiac |
| **126** | heart failure | Acute right heart failure | I50.811 | ICD10CM | cardiac |
| **127** | heart failure | Cardiac failure due to anesthesia during pregnancy, third trimester | O29.123 | ICD10CM | cardiac |
| **128** | heart failure | Postprocedural heart failure following other surgery | I97.131 | ICD10CM | cardiac |
| **129** | heart failure | Chronic diastolic (congestive) heart failure | I50.32 | ICD10CM | cardiac |
| **130** | heart failure | Presence of fully implantable artificial heart | Z95.812 | ICD10CM | cardiac |
| **131** | heart failure | Unspecified combined systolic (congestive) and diastolic (congestive) heart failure | I50.40 | ICD10CM | cardiac |
| **132** | heart failure | Chronic combined systolic (congestive) and diastolic (congestive) heart failure | I50.42 | ICD10CM | cardiac |
| **133** | heart failure | Presence of heart assist device | Z95.811 | ICD10CM | cardiac |
| **134** | heart failure | Cardiac failure due to anesthesia during pregnancy, unspecified trimester | O29.129 | ICD10CM | cardiac |
| **135** | heart failure | Other heart failure | I50.89 | ICD10CM | cardiac |
| **136** | heart failure | Biventricular heart failure | I50.82 | ICD10CM | cardiac |
| **137** | heart failure | Unspecified systolic (congestive) heart failure | I50.20 | ICD10CM | cardiac |
| **138** | heart failure | Acute combined systolic (congestive) and diastolic (congestive) heart failure | I50.41 | ICD10CM | cardiac |
| **139** | heart failure | Unspecified diastolic (congestive) heart failure | I50.30 | ICD10CM | cardiac |
| **140** | heart failure | Chronic systolic (congestive) heart failure | I50.22 | ICD10CM | cardiac |

| | | | | | |
|---|---|---|---|---|---|
| **141** | heart failure | End stage heart failure | I50.84 | ICD10CM | cardiac |
| **142** | heart failure | Postprocedural heart failure following cardiac surgery | I97.130 | ICD10CM | cardiac |
| **143** | heart failure | Right heart failure due to left heart failure | I50.814 | ICD10CM | cardiac |
| **144** | heart failure | Rheumatic heart failure | I09.81 | ICD10CM | cardiac |
| **145** | heart failure | High output heart failure | I50.83 | ICD10CM | cardiac |
| **146** | heart failure | Left ventricular failure, unspecified | I50.1 | ICD10CM | cardiac |
| **147** | heart failure | Cardiac failure due to anesthesia during pregnancy, second trimester | O29.122 | ICD10CM | cardiac |
| **148** | heart failure | Acute on chronic systolic (congestive) heart failure | I50.23 | ICD10CM | cardiac |
| **149** | heart failure | Acute on chronic diastolic (congestive) heart failure | I50.33 | ICD10CM | cardiac |
| **150** | heart failure | Right heart failure, unspecified | I50.810 | ICD10CM | cardiac |
| **151** | heart failure | Acute on chronic right heart failure | I50.813 | ICD10CM | cardiac |
| **152** | heart failure | Chronic right heart failure | I50.812 | ICD10CM | cardiac |
| **153** | heart failure | Acute on chronic combined systolic (congestive) and diastolic (congestive) heart failure | I50.43 | ICD10CM | cardiac |
| **154** | hypotension | Postprocedural hypotension | I95.81 | ICD10CM | cardiac |
| **155** | hypotension | Maternal hypotension syndrome, second trimester | O26.52 | ICD10CM | cardiac |
| **156** | hypotension | Idiopathic hypotension | I95.0 | ICD10CM | cardiac |
| **157** | hypotension | Orthostatic hypotension | I95.1 | ICD10CM | cardiac |
| **158** | hypotension | Hypotension due to drugs | I95.2 | ICD10CM | cardiac |
| **159** | hypotension | Maternal hypotension syndrome, third trimester | O26.53 | ICD10CM | cardiac |
| **160** | hypotension | Maternal hypotension syndrome, unspecified trimester | O26.50 | ICD10CM | cardiac |
| **161** | hypotension | Hypotension of hemodialysis | I95.3 | ICD10CM | cardiac |
| **162** | hypotension | Hypotension, unspecified | I95.9 | ICD10CM | cardiac |
| **163** | hypotension | Other hypotension | I95.89 | ICD10CM | cardiac |
| **164** | hypotension | Maternal hypotension syndrome, first trimester | O26.51 | ICD10CM | cardiac |
| **165** | myocarditis | Sarcoid myocarditis | D86.85 | ICD10CM | cardiac |
| **166** | myocarditis | Mumps myocarditis | B26.82 | ICD10CM | cardiac |
| **167** | myocarditis | Rheumatic myocarditis | I09.0 | ICD10CM | cardiac |
| **168** | myocarditis | Myocarditis, unspecified | I51.4 | ICD10CM | cardiac |
| **169** | myocarditis | Acute rheumatic myocarditis | I01.2 | ICD10CM | cardiac |
| **170** | myocarditis | Acute myocarditis, unspecified | I40.9 | ICD10CM | cardiac |
| **171** | myocarditis | Infective myocarditis | I40.0 | ICD10CM | cardiac |
| **172** | myocarditis | Influenza due to other identified influenza virus with myocarditis | J10.82 | ICD10CM | cardiac |
| **173** | myocarditis | Toxoplasma myocarditis | B58.81 | ICD10CM | cardiac |
| **174** | myocarditis | Meningococcal carditis, unspecified | A39.50 | ICD10CM | cardiac |
| **175** | myocarditis | Viral carditis, unspecified | B33.20 | ICD10CM | cardiac |
| **176** | myocarditis | Scarlet fever with myocarditis | A38.1 | ICD10CM | cardiac |
| **177** | myocarditis | Meningococcal myocarditis | A39.52 | ICD10CM | cardiac |

| | | | | | |
|---|---|---|---|---|---|
| **178** | myocarditis | Isolated myocarditis | I40.1 | ICD10CM | cardiac |
| **179** | myocarditis | Viral myocarditis | B33.22 | ICD10CM | cardiac |
| **180** | myocarditis | Influenza due to unidentified influenza virus with myocarditis | J11.82 | ICD10CM | cardiac |
| **181** | myocarditis | Myocarditis in diseases classified elsewhere | I41 | ICD10CM | cardiac |
| **182** | myocarditis | Acute myocarditis | I40 | ICD10CM | cardiac |
| **183** | myocarditis | Other acute myocarditis | I40.8 | ICD10CM | cardiac |
| **184** | pericarditis | Pericardial effusion (noninflammatory) | I31.3 | ICD10CM | cardiac |
| **185** | pericarditis | Chronic rheumatic pericarditis | I09.2 | ICD10CM | cardiac |
| **186** | pericarditis | Other specified diseases of pericardium | I31.8 | ICD10CM | cardiac |
| **187** | pericarditis | Chronic constrictive pericarditis | I31.1 | ICD10CM | cardiac |
| **188** | pericarditis | Acute nonspecific idiopathic pericarditis | I30.0 | ICD10CM | cardiac |
| **189** | pericarditis | Viral pericarditis | B33.23 | ICD10CM | cardiac |
| **190** | pericarditis | Pericarditis in diseases classified elsewhere | I32 | ICD10CM | cardiac |
| **191** | pericarditis | Acute pericarditis, unspecified | I30.9 | ICD10CM | cardiac |
| **192** | pericarditis | Disease of pericardium, unspecified | I31.9 | ICD10CM | cardiac |
| **193** | pericarditis | Other forms of acute pericarditis | I30.8 | ICD10CM | cardiac |
| **194** | pericarditis | Chronic adhesive pericarditis | I31.0 | ICD10CM | cardiac |
| **195** | pericarditis | Acute rheumatic pericarditis | I01.0 | ICD10CM | cardiac |
| **196** | pericarditis | Meningococcal pericarditis | A39.53 | ICD10CM | cardiac |
| **197** | pericarditis | Hemopericardium, not elsewhere classified | I31.2 | ICD10CM | cardiac |
| **198** | pericarditis | Cardiac tamponade | I31.4 | ICD10CM | cardiac |
| **199** | pericarditis | Acute pericarditis | I30 | ICD10CM | cardiac |
| **200** | pericarditis | Infective pericarditis | I30.1 | ICD10CM | cardiac |
| **201** | Troponin | Troponin T(Highly Sensitive) \| 89579-7 | 89579-7 | LOINC | cardiac |
| **202** | Troponin | TROPONIN T -LC\|TROPONIN T(HIGHLY SENSITIVE) 18796 -LC\|67151-1 | 67151-1 | LOINC | cardiac |
| **203** | Troponin | TROPONIN I - UCH MN \| TROPONIN I - UCH MN \| 10839- | 10839-9 | LOINC | cardiac |
| **204** | Troponin | TROPONIN T\|6598-7 | 48425-3 | LOINC | cardiac |
| **205** | Troponin | TROPONIN\|10839-9 | 10839-9 | LOINC | cardiac |
| **206** | Troponin | TROPONIN T(HIGHLY SENSITIVE)\|67151-1 | 67151-1 | LOINC | cardiac |
| **207** | Troponin | TROPONIN T\|6598-7 | 6598-7 | LOINC | cardiac |
| **208** | Troponin | TROPONIN I-STAT\|42757-5 | 42757-5 | LOINC | cardiac |
| **209** | Troponin | Troponin-I | 10839-9 | LOINC | cardiac |
| **210** | Troponin | TROPONIN I, PLASMA\|10839-9 | 10839-9 | LOINC | cardiac |
| **211** | Troponin | TROPONIN I\|10839-9 | 10839-9 | LOINC | cardiac |
| **212** | Troponin | Troponin I \| Troponin I | 121870001 | SNOMED | cardiac |
| **213** | Troponin | TROPONIN-I | 10839-9 | LOINC | cardiac |
| **214** | Troponin | TROPONIN-T\|48425-3 | 48425-3 | LOINC | cardiac |
| **215** | Troponin | TROPONIN I - LC\|TROPONIN I\|10839-9 | 10839-9 | LOINC | cardiac |

| | | | | | |
|---|---|---|---|---|---|
| **216** | Troponin | TROPONIN I\|49563-0 | 10839-9 | LOINC | cardiac |
| **217** | Troponin | TROPONIN I -Q\|TROPONIN I\|10839-9 | 10839-9 | LOINC | cardiac |
| **218** | Troponin | TROPONIN-I\|10839-9 | 10839-9 | LOINC | cardiac |
| **219** | Troponin | TROPONIN I\|TROPONIN-I\|10839-9 | 10839-9 | LOINC | cardiac |
| **220** | Troponin | TROPONIN \| TROPONIN \| 10839-9 \| 1230100229 | 10839-9 | LOINC | cardiac |
| **221** | Troponin | TROPONIN T -LC\|TROPONIN T(HIGHLY SENSITIVE) 18796 -LC\|89579-7 | 89579-7 | LOINC | cardiac |
| **222** | conjunctivitis | Other mucopurulent conjunctivitis, unspecified eye | H10.029 | ICD10CM | dermatologic |
| **223** | conjunctivitis | Fitting and adjustment of orthopedic devices | V53.7 | ICD9CM | dermatologic |
| **224** | conjunctivitis | Conjunctivitis, unspecified | 372.3 | ICD9CM | dermatologic |
| **225** | conjunctivitis | Other mucopurulent conjunctivitis, right eye | H10.021 | ICD10CM | dermatologic |
| **226** | conjunctivitis | Unspecified acute conjunctivitis, left eye | H10.32 | ICD10CM | dermatologic |
| **227** | conjunctivitis | Other cyst of bone, left thigh | M85.652 | ICD10CM | dermatologic |
| **228** | conjunctivitis | Acute chemical conjunctivitis | 372.06 | ICD9CM | dermatologic |
| **229** | conjunctivitis | Other conjunctivitis | H10.89 | ICD10CM | dermatologic |
| **230** | conjunctivitis | Other chronic allergic conjunctivitis | H10.45 | ICD10CM | dermatologic |
| **231** | conjunctivitis | Other disorders of bone development and growth, right femur | M89.251 | ICD10CM | dermatologic |
| **232** | conjunctivitis | Emergency use of U07.1 \| COVID-19 | U07.1 | ICD10CM | dermatologic |
| **233** | conjunctivitis | Unspecified acute conjunctivitis, bilateral | H10.33 | ICD10CM | dermatologic |
| **234** | conjunctivitis | Other mucopurulent conjunctivitis, bilateral | H10.023 | ICD10CM | dermatologic |
| **235** | conjunctivitis | Unspecified conjunctivitis | H10.9 | ICD10CM | dermatologic |
| **236** | conjunctivitis | Viral conjunctivitis, unspecified | B30.9 | ICD10CM | dermatologic |
| **237** | conjunctivitis | Vernal conjunctivitis | H10.44 | ICD10CM | dermatologic |
| **238** | conjunctivitis | Other mucopurulent conjunctivitis, left eye | H10.022 | ICD10CM | dermatologic |
| **239** | conjunctivitis | Other mucopurulent conjunctivitis | 372.03 | ICD9CM | dermatologic |
| **240** | conjunctivitis | Acute atopic conjunctivitis | 372.05 | ICD9CM | dermatologic |
| **241** | conjunctivitis | Other chronic allergic conjunctivitis | 372.14 | ICD9CM | dermatologic |
| **242** | conjunctivitis | Acute atopic conjunctivitis, bilateral | H10.13 | ICD10CM | dermatologic |
| **243** | conjunctivitis | Acute atopic conjunctivitis, unspecified eye | H10.10 | ICD10CM | dermatologic |
| **244** | conjunctivitis | Localized edema | R60.0 | ICD10CM | dermatologic |
| **245** | conjunctivitis | Acute conjunctivitis, unspecified | 372 | ICD9CM | dermatologic |
| **246** | conjunctivitis | Unspecified acute conjunctivitis, right eye | H10.31 | ICD10CM | dermatologic |
| **247** | conjunctivitis | Unspecified acute conjunctivitis, unspecified eye | H10.30 | ICD10CM | dermatologic |
| **248** | conjunctivitis | Other viral conjunctivitis | B30.8 | ICD10CM | dermatologic |
| **249** | conjunctivitis | Unspecified diseases of conjunctiva due to viruses | 77.99 | ICD9CM | dermatologic |
| **250** | skin rashes | Rash and other nonspecific skin eruption | R21 | ICD10CM | dermatologic |
| **251** | skins signs and symptoms | Other disturbances of skin sensation | R20.8 | ICD10CM | dermatologic |
| **252** | skins signs and symptoms | Localized swelling, mass and lump, unspecified | R22.9 | ICD10CM | dermatologic |
| **253** | skins signs and symptoms | Other skin changes | R23 | ICD10CM | dermatologic |
| **254** | skins signs and symptoms | Hypoesthesia of skin | R20.1 | ICD10CM | dermatologic |

| | | | | | |
|---|---|---|---|---|---|
| **255** | skins signs and symptoms | Paresthesia of skin | R20.2 | ICD10CM | dermatologic |
| **256** | skins signs and symptoms | Changes in skin texture | R23.4 | ICD10CM | dermatologic |
| **257** | skins signs and symptoms | Localized swelling, mass and lump, upper limb | R22.3 | ICD10CM | dermatologic |
| **258** | skins signs and symptoms | Spontaneous ecchymoses | R23.3 | ICD10CM | dermatologic |
| **259** | skins signs and symptoms | Localized swelling, mass and lump of skin and subcutaneous tissue | R22 | ICD10CM | dermatologic |
| **260** | skins signs and symptoms | Localized swelling, mass and lump, lower limb | R22.4 | ICD10CM | dermatologic |
| **261** | skins signs and symptoms | Localized swelling, mass and lump, neck | R22.1 | ICD10CM | dermatologic |
| **262** | skins signs and symptoms | Localized swelling, mass and lump, head | R22.0 | ICD10CM | dermatologic |
| **263** | skins signs and symptoms | Localized swelling, mass and lump, right upper limb | R22.31 | ICD10CM | dermatologic |
| **264** | skins signs and symptoms | Localized swelling, mass and lump, left upper limb | R22.32 | ICD10CM | dermatologic |
| **265** | skins signs and symptoms | Disturbances of skin sensation | R20 | ICD10CM | dermatologic |
| **266** | skins signs and symptoms | Unspecified disturbances of skin sensation | R20.9 | ICD10CM | dermatologic |
| **267** | skins signs and symptoms | Anesthesia of skin | R20.0 | ICD10CM | dermatologic |
| **268** | skins signs and symptoms | Localized swelling, mass and lump, upper limb, bilateral | R22.33 | ICD10CM | dermatologic |
| **269** | skins signs and symptoms | Flushing | R23.2 | ICD10CM | dermatologic |
| **270** | skins signs and symptoms | Localized swelling, mass and lump, lower limb, bilateral | R22.43 | ICD10CM | dermatologic |
| **271** | skins signs and symptoms | Localized swelling, mass and lump, right lower limb | R22.41 | ICD10CM | dermatologic |
| **272** | skins signs and symptoms | Localized swelling, mass and lump, unspecified lower limb | R22.40 | ICD10CM | dermatologic |
| **273** | skins signs and symptoms | Localized swelling, mass and lump, trunk | R22.2 | ICD10CM | dermatologic |
| **274** | skins signs and symptoms | Hyperesthesia | R20.3 | ICD10CM | dermatologic |
| **275** | skins signs and symptoms | Other skin changes | R23.8 | ICD10CM | dermatologic |
| **276** | skins signs and symptoms | Pallor | R23.1 | ICD10CM | dermatologic |
| **277** | skins signs and symptoms | Localized swelling, mass and lump, unspecified upper limb | R22.30 | ICD10CM | dermatologic |
| **278** | skins signs and symptoms | Unspecified skin changes | R23.9 | ICD10CM | dermatologic |
| **279** | skins signs and symptoms | Localized swelling, mass and lump, left lower limb | R22.42 | ICD10CM | dermatologic |
| **280** | abdominal pain | Generalized abdominal tenderness | R10.817 | ICD10CM | gastrointestinal |
| **281** | abdominal pain | Right upper quadrant abdominal tenderness | R10.811 | ICD10CM | gastrointestinal |
| **282** | abdominal pain | Lower abdominal pain, unspecified | R10.30 | ICD10CM | gastrointestinal |
| **283** | abdominal pain | Epigastric abdominal tenderness | R10.816 | ICD10CM | gastrointestinal |
| **284** | abdominal pain | Epigastric rebound abdominal tenderness | R10.826 | ICD10CM | gastrointestinal |
| **285** | abdominal pain | Left lower quadrant abdominal rigidity | R19.34 | ICD10CM | gastrointestinal |
| **286** | abdominal pain | Periumbilic abdominal tenderness | R10.815 | ICD10CM | gastrointestinal |
| **287** | abdominal pain | Left upper quadrant pain | R10.12 | ICD10CM | gastrointestinal |
| **288** | abdominal pain | Unspecified abdominal pain | R10.9 | ICD10CM | gastrointestinal |
| **289** | abdominal pain | Right upper quadrant rebound abdominal tenderness | R10.821 | ICD10CM | gastrointestinal |

| | | | | | |
|---|---|---|---|---|---|
| **290** | abdominal pain | Left lower quadrant abdominal tenderness | R10.814 | ICD10CM | gastrointestinal |
| **291** | abdominal pain | Abdominal tenderness, unspecified site | R10.819 | ICD10CM | gastrointestinal |
| **292** | abdominal pain | Right upper quadrant pain | R10.11 | ICD10CM | gastrointestinal |
| **293** | abdominal pain | Periumbilic rebound abdominal tenderness | R10.825 | ICD10CM | gastrointestinal |
| **294** | abdominal pain | Periumbilic abdominal rigidity | R19.35 | ICD10CM | gastrointestinal |
| **295** | abdominal pain | Pelvic and perineal pain | R10.2 | ICD10CM | gastrointestinal |
| **296** | abdominal pain | Colic | R10.83 | ICD10CM | gastrointestinal |
| **297** | abdominal pain | Left lower quadrant rebound abdominal tenderness | R10.824 | ICD10CM | gastrointestinal |
| **298** | abdominal pain | Generalized abdominal rigidity | R19.37 | ICD10CM | gastrointestinal |
| **299** | abdominal pain | Left upper quadrant abdominal rigidity | R19.32 | ICD10CM | gastrointestinal |
| **300** | abdominal pain | Acute abdomen | R10.0 | ICD10CM | gastrointestinal |
| **301** | abdominal pain | Generalized rebound abdominal tenderness | R10.827 | ICD10CM | gastrointestinal |
| **302** | abdominal pain | Upper abdominal pain, unspecified | R10.10 | ICD10CM | gastrointestinal |
| **303** | abdominal pain | Right lower quadrant abdominal tenderness | R10.813 | ICD10CM | gastrointestinal |
| **304** | abdominal pain | Left lower quadrant pain | R10.32 | ICD10CM | gastrointestinal |
| **305** | abdominal pain | Epigastric abdominal rigidity | R19.36 | ICD10CM | gastrointestinal |
| **306** | abdominal pain | Left upper quadrant abdominal tenderness | R10.812 | ICD10CM | gastrointestinal |
| **307** | abdominal pain | Rebound abdominal tenderness, unspecified site | R10.829 | ICD10CM | gastrointestinal |
| **308** | abdominal pain | Left upper quadrant rebound abdominal tenderness | R10.822 | ICD10CM | gastrointestinal |
| **309** | abdominal pain | Epigastric pain | R10.13 | ICD10CM | gastrointestinal |
| **310** | abdominal pain | Periumbilical pain | R10.33 | ICD10CM | gastrointestinal |
| **311** | abdominal pain | Abdominal rigidity, unspecified site | R19.30 | ICD10CM | gastrointestinal |
| **312** | abdominal pain | Right lower quadrant rebound abdominal tenderness | R10.823 | ICD10CM | gastrointestinal |
| **313** | abdominal pain | Right lower quadrant pain | R10.31 | ICD10CM | gastrointestinal |
| **314** | abdominal pain | Right lower quadrant abdominal rigidity | R19.33 | ICD10CM | gastrointestinal |
| **315** | abdominal pain | Right upper quadrant abdominal rigidity | R19.31 | ICD10CM | gastrointestinal |
| **316** | abdominal pain | Generalized abdominal pain | R10.84 | ICD10CM | gastrointestinal |
| **317** | abdominal pain | Gas pain | R14.1 | ICD10CM | gastrointestinal |
| **318** | abnormal liver enzymes | Elevation of levels of liver transaminase levels | R74.01 | ICD10CM | gastrointestinal |
| **319** | abnormal liver enzymes | Nonspecific elevation of levels of transaminase and lactic acid dehydrogenase [LDH] | R74.0 | ICD10CM | gastrointestinal |
| **320** | abnormal liver enzymes | Elevation of levels of lactic acid dehydrogenase [LDH] | R74.02 | ICD10CM | gastrointestinal |
| **321** | abnormal liver enzymes | Abnormal results of liver function studies | R94.5 | ICD10CM | gastrointestinal |
| **322** | diarrhea | Diarrhea, unspecified | R19.7 | ICD10CM | gastrointestinal |
| **323** | gastroenteritis | Viral and other specified intestinal infections | A08 | ICD10CM | gastrointestinal |
| **324** | gastroenteritis | Other microscopic colitis | K52.838 | ICD10CM | gastrointestinal |
| **325** | gastroenteritis | Toxic gastroenteritis and colitis | K52.1 | ICD10CM | gastrointestinal |
| **326** | gastroenteritis | Eosinophilic colitis | K52.82 | ICD10CM | gastrointestinal |
| **327** | gastroenteritis | Infectious gastroenteritis and colitis, unspecified | A09 | ICD10CM | gastrointestinal |

| | | | | | |
|---|---|---|---|---|---|
| **328** | gastroenteritis | Acute gastroenteropathy due to Norwalk agent and other small round viruses | A08.1 | ICD10CM | gastrointestinal |
| **329** | gastroenteritis | Other specified noninfective gastroenteritis and colitis | K52.8 | ICD10CM | gastrointestinal |
| **330** | gastroenteritis | Food protein-induced enterocolitis syndrome | K52.21 | ICD10CM | gastrointestinal |
| **331** | gastroenteritis | Calicivirus enteritis | A08.31 | ICD10CM | gastrointestinal |
| **332** | gastroenteritis | Other specified intestinal infections | A08.8 | ICD10CM | gastrointestinal |
| **333** | gastroenteritis | Allergic and dietetic gastroenteritis and colitis | K52.2 | ICD10CM | gastrointestinal |
| **334** | gastroenteritis | Other viral enteritis | A08.39 | ICD10CM | gastrointestinal |
| **335** | gastroenteritis | Microscopic colitis, unspecified | K52.839 | ICD10CM | gastrointestinal |
| **336** | gastroenteritis | Other viral enteritis | A08.3 | ICD10CM | gastrointestinal |
| **337** | gastroenteritis | Lymphocytic colitis | K52.832 | ICD10CM | gastrointestinal |
| **338** | gastroenteritis | Viral intestinal infection, unspecified | A08.4 | ICD10CM | gastrointestinal |
| **339** | gastroenteritis | Collagenous colitis | K52.831 | ICD10CM | gastrointestinal |
| **340** | gastroenteritis | Microscopic colitis | K52.83 | ICD10CM | gastrointestinal |
| **341** | gastroenteritis | Noninfective gastroenteritis and colitis, unspecified | K52.9 | ICD10CM | gastrointestinal |
| **342** | gastroenteritis | Other and unspecified noninfective gastroenteritis and colitis | K52 | ICD10CM | gastrointestinal |
| **343** | gastroenteritis | Eosinophilic gastritis or gastroenteritis | K52.81 | ICD10CM | gastrointestinal |
| **344** | gastroenteritis | Acute gastroenteropathy due to Norwalk agent | A08.11 | ICD10CM | gastrointestinal |
| **345** | gastroenteritis | Astrovirus enteritis | A08.32 | ICD10CM | gastrointestinal |
| **346** | gastroenteritis | Food protein-induced enteropathy | K52.22 | ICD10CM | gastrointestinal |
| **347** | gastroenteritis | Other allergic and dietetic gastroenteritis and colitis | K52.29 | ICD10CM | gastrointestinal |
| **348** | gastroenteritis | Gastroenteritis and colitis due to radiation | K52.0 | ICD10CM | gastrointestinal |
| **349** | gastroenteritis | Acute gastroenteropathy due to other small round viruses | A08.19 | ICD10CM | gastrointestinal |
| **350** | gastroenteritis | Adenoviral enteritis | A08.2 | ICD10CM | gastrointestinal |
| **351** | gastroenteritis | Other specified noninfective gastroenteritis and colitis | K52.89 | ICD10CM | gastrointestinal |
| **352** | gastroenteritis | Indeterminate colitis | K52.3 | ICD10CM | gastrointestinal |
| **353** | gastroenteritis | Rotaviral enteritis | A08.0 | ICD10CM | gastrointestinal |
| **354** | intestinal obstruction and ileus | Intestinal adhesions [bands] with obstruction (postinfection) | K56.5 | ICD10CM | gastrointestinal |
| **355** | intestinal obstruction and ileus | Paralytic ileus | K56.0 | ICD10CM | gastrointestinal |
| **356** | intestinal obstruction and ileus | Intestinal adhesions [bands], unspecified as to partial versus complete obstruction | K56.50 | ICD10CM | gastrointestinal |
| **357** | intestinal obstruction and ileus | Other intestinal obstruction | K56.69 | ICD10CM | gastrointestinal |
| **358** | intestinal obstruction and ileus | Postprocedural intestinal obstruction | K91.3 | ICD10CM | gastrointestinal |
| **359** | intestinal obstruction and ileus | Intussusception | K56.1 | ICD10CM | gastrointestinal |
| **360** | intestinal obstruction and ileus | Unspecified intestinal obstruction | K56.60 | ICD10CM | gastrointestinal |

| | | | | | |
|---|---|---|---|---|---|
| **361** | intestinal obstruction and ileus | Other intestinal obstruction unspecified as to partial versus complete obstruction | K56.699 | ICD10CM | gastrointestinal |
| **362** | intestinal obstruction and ileus | Complete intestinal obstruction, unspecified as to cause | K56.601 | ICD10CM | gastrointestinal |
| **363** | intestinal obstruction and ileus | Other impaction of intestine | K56.49 | ICD10CM | gastrointestinal |
| **364** | intestinal obstruction and ileus | Intestinal adhesions [bands], with partial obstruction | K56.51 | ICD10CM | gastrointestinal |
| **365** | intestinal obstruction and ileus | Other complete intestinal obstruction | K56.691 | ICD10CM | gastrointestinal |
| **366** | intestinal obstruction and ileus | Fecal impaction | K56.41 | ICD10CM | gastrointestinal |
| **367** | intestinal obstruction and ileus | Ileus, unspecified | K56.7 | ICD10CM | gastrointestinal |
| **368** | intestinal obstruction and ileus | Postprocedural complete intestinal obstruction | K91.32 | ICD10CM | gastrointestinal |
| **369** | intestinal obstruction and ileus | Volvulus | K56.2 | ICD10CM | gastrointestinal |
| **370** | intestinal obstruction and ileus | Obstruction of duodenum | K31.5 | ICD10CM | gastrointestinal |
| **371** | intestinal obstruction and ileus | Unspecified intestinal obstruction, unspecified as to partial versus complete obstruction | K56.609 | ICD10CM | gastrointestinal |
| **372** | intestinal obstruction and ileus | Other partial intestinal obstruction | K56.690 | ICD10CM | gastrointestinal |
| **373** | intestinal obstruction and ileus | Partial intestinal obstruction, unspecified as to cause | K56.600 | ICD10CM | gastrointestinal |
| **374** | intestinal obstruction and ileus | Intestinal adhesions [bands] with complete obstruction | K56.52 | ICD10CM | gastrointestinal |
| **375** | intestinal obstruction and ileus | Postprocedural partial intestinal obstruction | K91.31 | ICD10CM | gastrointestinal |
| **376** | intestinal obstruction and ileus | Postprocedural intestinal obstruction, unspecified as to partial versus complete | K91.30 | ICD10CM | gastrointestinal |
| **377** | nausea and vomiting | Vomiting following gastrointestinal surgery | K91.0 | ICD10CM | gastrointestinal |
| **378** | nausea and vomiting | Nausea with vomiting, unspecified | R11.2 | ICD10CM | gastrointestinal |
| **379** | nausea and vomiting | Vomiting of fecal matter | R11.13 | ICD10CM | gastrointestinal |
| **380** | nausea and vomiting | Vomiting | R11.1 | ICD10CM | gastrointestinal |
| **381** | nausea and vomiting | Vomiting without nausea | R11.11 | ICD10CM | gastrointestinal |
| **382** | nausea and vomiting | Other vomiting without nausea | R11.3 | ICD10CM | gastrointestinal |
| **383** | nausea and vomiting | Vomiting, unspecified | R11.10 | ICD10CM | gastrointestinal |
| **384** | nausea and vomiting | Bilious vomiting | R11.14 | ICD10CM | gastrointestinal |
| **385** | nausea and vomiting | Projectile vomiting | R11.12 | ICD10CM | gastrointestinal |
| **386** | nausea and vomiting | Cyclical vomiting syndrome unrelated to migraine | R11.15 | ICD10CM | gastrointestinal |
| **387** | nausea and vomiting | Late vomiting of pregnancy | O21.2 | ICD10CM | gastrointestinal |
| **388** | nausea and vomiting | Other vomiting complicating pregnancy | O21.8 | ICD10CM | gastrointestinal |
| **389** | nausea and vomiting | Vomiting of pregnancy, unspecified | O21.9 | ICD10CM | gastrointestinal |
| **390** | nausea and vomiting | Nausea and vomiting | R11 | ICD10CM | gastrointestinal |
| **391** | nausea and vomiting | Nausea | R11.0 | ICD10CM | gastrointestinal |

| | | | | | |
|---|---|---|---|---|---|
| **392** | aplastic anemia | Other drug-induced pancytopenia | D61.811 | ICD10CM | hematological |
| **393** | aplastic anemia | Other constitutional aplastic anemia | D61.09 | ICD10CM | hematological |
| **394** | aplastic anemia | Congenital dyserythropoietic anemia | D64.4 | ICD10CM | hematological |
| **395** | aplastic anemia | Anemia in chronic kidney disease | D63.1 | ICD10CM | hematological |
| **396** | aplastic anemia | Anemia in other chronic diseases classified elsewhere | D63.8 | ICD10CM | hematological |
| **397** | aplastic anemia | Antineoplastic chemotherapy induced pancytopenia | D61.810 | ICD10CM | hematological |
| **398** | aplastic anemia | Aplastic anemia, unspecified | D61.9 | ICD10CM | hematological |
| **399** | aplastic anemia | Hereditary sideroblastic anemia | D64.0 | ICD10CM | hematological |
| **400** | aplastic anemia | Anemia due to antineoplastic chemotherapy | D64.81 | ICD10CM | hematological |
| **401** | aplastic anemia | Aplastic anemia due to other external agents | D61.2 | ICD10CM | hematological |
| **402** | aplastic anemia | Anemia, unspecified | D64.9 | ICD10CM | hematological |
| **403** | aplastic anemia | Idiopathic aplastic anemia | D61.3 | ICD10CM | hematological |
| **404** | aplastic anemia | Other anemias | D64 | ICD10CM | hematological |
| **405** | aplastic anemia | Other specified anemias | D64.89 | ICD10CM | hematological |
| **406** | aplastic anemia | Acquired pure red cell aplasia, unspecified | D60.9 | ICD10CM | hematological |
| **407** | aplastic anemia | Other pancytopenia | D61.818 | ICD10CM | hematological |
| **408** | aplastic anemia | Secondary sideroblastic anemia due to drugs and toxins | D64.2 | ICD10CM | hematological |
| **409** | aplastic anemia | Secondary sideroblastic anemia due to disease | D64.1 | ICD10CM | hematological |
| **410** | aplastic anemia | Anemia in neoplastic disease | D63.0 | ICD10CM | hematological |
| **411** | aplastic anemia | Other acquired pure red cell aplasias | D60.8 | ICD10CM | hematological |
| **412** | aplastic anemia | Drug-induced aplastic anemia | D61.1 | ICD10CM | hematological |
| **413** | aplastic anemia | Constitutional aplastic anemia | D61.0 | ICD10CM | hematological |
| **414** | aplastic anemia | Other specified aplastic anemias and other bone marrow failure syndromes | D61.89 | ICD10CM | hematological |
| **415** | aplastic anemia | Pancytopenia | D61.81 | ICD10CM | hematological |
| **416** | aplastic anemia | Chronic acquired pure red cell aplasia | D60.0 | ICD10CM | hematological |
| **417** | aplastic anemia | Constitutional (pure) red blood cell aplasia | D61.01 | ICD10CM | hematological |
| **418** | aplastic anemia | Other specified aplastic anemias and other bone marrow failure syndromes | D61.8 | ICD10CM | hematological |
| **419** | aplastic anemia | Acquired pure red cell aplasia [erythroblastopenia] | D60 | ICD10CM | hematological |
| **420** | aplastic anemia | Other specified anemias | D64.8 | ICD10CM | hematological |
| **421** | aplastic anemia | Other aplastic anemias and other bone marrow failure syndromes | D61 | ICD10CM | hematological |
| **422** | aplastic anemia | Transient acquired pure red cell aplasia | D60.1 | ICD10CM | hematological |
| **423** | aplastic anemia | Myelophthisis | D61.82 | ICD10CM | hematological |
| **424** | aplastic anemia | Other sideroblastic anemias | D64.3 | ICD10CM | hematological |
| **425** | D-dimer | D-DIMER\|48065-7 | 48065-7 | LOINC | hematological |
| **426** | D-dimer | D-DIMER \| D-DIMER \| 48065-7 \| 1230300005 | 48065-7 | LOINC | hematological |
| **427** | D-dimer | D-Dimer | 48065-7 | LOINC | hematological |
| **428** | D-dimer | D DIMER\|D-DIMER - LC\|48065-7 | 48065-7 | LOINC | hematological |

| | | | | | |
|---|---|---|---|---|---|
| **429** | D-dimer | D-DIMER QUANTITATIVE\|48065-7 | 48065-7 | LOINC | hematological |
| **430** | D-dimer | D-DIMER, PLASMA\|48065-7 | 48065-7 | LOINC | hematological |
| **431** | D-dimer | D-Dimer, Quantitative \| 48065-7 | 48065-7 | LOINC | hematological |
| **432** | D-dimer | D DIMER\|D DIMER\|85379 | 48065-7 | LOINC | hematological |
| **433** | D-dimer | D DIMER\|D DIMER\|15179-5 | 15179-5 | LOINC | hematological |
| **434** | D-dimer | D-Dimer \| 48065-7 | 48065-7 | LOINC | hematological |
| **435** | D-dimer | D-DIMER,QUANTITATIVE (NO EQUATION) - UCH MN \| D-DI | 48065-7 | LOINC | hematological |
| **436** | D-dimer | D DIMER\|D DIMER\|15179-5 | 48065-7 | LOINC | hematological |
| **437** | D-dimer | QUANTITATIVE D-DIMER\|48065-7 | 48065-7 | LOINC | hematological |
| **438** | D-dimer | D-Dimer, ELISA \| 48065-7 | 48065-7 | LOINC | hematological |
| **439** | D-dimer | D-Dimer \| D-Dimer | 70648006 | SNOMED | hematological |
| **440** | D-dimer | D-DIMER | 48065-7 | LOINC | hematological |
| **441** | D-dimer | D-DIMER,QUANTITATIVE (NO EQUATION) - UCH MN \| D DI | 48065-7 | LOINC | hematological |
| **442** | D-dimer | D DIMER\|D-DIMER, QUANTITATIVE-Q\|48065-7 | 48065-7 | LOINC | hematological |
| **443** | D-dimer | ANTITHROMBIN III\|48065-7 | 48065-7 | LOINC | hematological |
| **444** | D-dimer | D-DIMER, QUANTITATIVE\|48065-7 | 48065-7 | LOINC | hematological |
| **445** | Lymphocytes | LYMPHOCYTE ABSOLUTE | 26474-7 | LOINC | hematological |
| **446** | Lymphocytes | LYMPHOCYTES (ABS #) \| CBC WITH DIFF \| 26474-7 \| 71 | 26474-7 | LOINC | hematological |
| **447** | Lymphocytes | LYMPHOCYTES ABSOLUTE (AUTO DIFFERENTIAL) - UCH MN | 731-0 | LOINC | hematological |
| **448** | Lymphocytes | LYMPHOCYTES (ABS #) - NOC \| CBC WITH AUTO DIFF - N | 731-0 | LOINC | hematological |
| **449** | Lymphocytes | LYMPHS EXT\|731-0 | 731-0 | LOINC | hematological |
| **450** | Lymphocytes | DIFFERENTIAL, MANUAL -Q\|ABSOLUTE LYMPHOCYTES\|91341.001 | 26474-7 | LOINC | hematological |
| **451** | Lymphocytes | LYMPH ABSOLUTE\|731-0 | 731-0 | LOINC | hematological |
| **452** | Lymphocytes | Lymphocytes, Absolute | 26474-7 | LOINC | hematological |
| **453** | Lymphocytes | Lymphocytes, Variant \| Lymphocytes, Variant | 74765001 | SNOMED | hematological |
| **454** | Lymphocytes | CBC W/DIFF AND PLATELETS -Q\|ABSOLUTE LYMPHOCYTES\|85025 | 26474-7 | LOINC | hematological |
| **455** | Lymphocytes | DIFFERENTIAL, MANUAL -Q\|ABSOLUTE LYMPHOCYTES\|415.002 | 26474-7 | LOINC | hematological |
| **456** | Lymphocytes | LYMPHOCYTES ABSOLUTE\|731-0 | 731-0 | LOINC | hematological |
| **457** | Lymphocytes | CBC,PLATELET, WITH DIFFERENTIAL\|ABSOLUTE LYMPHOCYTES\|85025 | 26474-7 | LOINC | hematological |
| **458** | Lymphocytes | Lymphocytes, Abs. \| Lymphocytes, Abs. | 74765001 | SNOMED | hematological |
| **459** | Lymphocytes | LYMPHOCYTE ABS (MANUAL DIFFERENTIAL)(INTERFACED) - | 732-8 | LOINC | hematological |
| **460** | Lymphocytes | Lymphocytes, ABS (man diff) \| 732-8 | 732-8 | LOINC | hematological |
| **461** | Lymphocytes | LYMPHOCYTES, ABSOLUTE (EXT RSLT)\|731-0 | 731-0 | LOINC | hematological |

| | | | | | |
|---|---|---|---|---|---|
| **462** | Lymphocytes | CBC,PLATELET, WITH DIFFERENTIAL\|ABSOLUTE LYMPHOCYTES\|15192-8 | 26474-7 | LOINC | hematological |
| **463** | Lymphocytes | LYMPHOCYTES, ABSOLUTE\|731-0 | 731-0 | LOINC | hematological |
| **464** | Lymphocytes | LYMPHOCYTES (ABS #) \| CBC AUTODIFF FOR SEPSIS \| 26 | 26474-7 | LOINC | hematological |
| **465** | Lymphocytes | MANUAL DIFFERENTIAL-WAM\|ABSOLUTE LYMPHOCYTES-WAM\|732-8 | 732-8 | LOINC | hematological |
| **466** | Lymphocytes | ABSOLUTE LYMPHS\|26474-7 | 26474-7 | LOINC | hematological |
| **467** | Lymphocytes | CBC,PLATELET, WITH DIFFERENTIAL\|ABSOLUTE LYMPHOCYTES-Q\|731-0 | 26474-7 | LOINC | hematological |
| **468** | Lymphocytes | CBC W/DIFF AND PLATELETS -Q\|ABSOLUTE LYMPHOCYTES-Q\|731-0 | 26474-7 | LOINC | hematological |
| **469** | Lymphocytes | Lymphocytes \| 732-8 | 732-8 | LOINC | hematological |
| **470** | Lymphocytes | LYMPHOCYTES (ABS #) \| CBC WITH AUTODIFF \| 26474-7 | 26474-7 | LOINC | hematological |
| **471** | Lymphocytes | ABSOLUTE LYMPHOCYTES\|731-0 | 26474-7 | LOINC | hematological |
| **472** | Lymphocytes | Lymphocyte, Absolute \| 731-0 | 731-0 | LOINC | hematological |
| **473** | Lymphocytes | CBC W/DIFF AND PLATELETS -Q\|ABSOLUTE LYMPHOCYTES-Q\|85025 | 26474-7 | LOINC | hematological |
| **474** | Lymphocytes | CBC,PLATELET, WITH DIFFERENTIAL\|ABSOLUTE LYMPHOCYTES\|732-8 | 26474-7 | LOINC | hematological |
| **475** | Lymphocytes | ABSOLUTE LYMPHOCYTE COUNT\|26474-7 | 26474-7 | LOINC | hematological |
| **476** | Lymphocytes | LYMPHOCYTE ABSOLUTE\|731-0 | 731-0 | LOINC | hematological |
| **477** | Lymphocytes | LYMPHOCYTE\|736-9 | 26474-7 | LOINC | hematological |
| **478** | Lymphocytes | Absolute Lymphocytes \| 731-0 | 731-0 | LOINC | hematological |
| **479** | Lymphocytes | LYM\|26474-7 | 26474-7 | LOINC | hematological |
| **480** | Lymphocytes | DIFFERENTIAL, MANUAL -Q\|ABSOLUTE LYMPHOCYTES\|732-8 | 26474-7 | LOINC | hematological |
| **481** | Lymphocytes | CBC,PLATELET, WITH DIFFERENTIAL\|ABSOLUTE LYMPHOCYTES AUTOMATED COUNT\|731-0 | 731-0 | LOINC | hematological |
| **482** | Lymphocytes | ABSOLUTE LYMPHOCYTE, POC\|731-0 | 731-0 | LOINC | hematological |
| **483** | Lymphocytes | ABS LYMPHOCYTE COUNT\|26474-7 | 26474-7 | LOINC | hematological |
| **484** | Lymphocytes | ABSOLUTE LYMPHOCYTES\|731-0 | 731-0 | LOINC | hematological |
| **485** | Lymphocytes | LYM, ABS \| 731-0 | 731-0 | LOINC | hematological |
| **486** | Lymphocytes | LYMPHOCYTES (ABS #) \| CBC SEPSIS W/DIFF \| 26474-7 | 26474-7 | LOINC | hematological |
| **487** | Lymphocytes | ABSOLUTE LYMPHOCYTE COUNT\|731-0 | 731-0 | LOINC | hematological |
| **488** | Platelets | PLATELET COUNT \| CBC WITH DIFF \| 777-3 \| 24 | 777-3 | LOINC | hematological |
| **489** | Platelets | PLATELET ESTIMATION WAM (LAB ORDER ONLY)\|PLATELET ESTIMATE\|85025.004 | 26515-7 | LOINC | hematological |
| **490** | Platelets | CBC WITH DIFFERENTIAL/PLATELET -LC\|PLATELETS-LC\|85025 | 26515-7 | LOINC | hematological |
| **491** | Platelets | PLATELETS, BLOOD\|777-3 | 777-3 | LOINC | hematological |

| | | | | | |
|---|---|---|---|---|---|
| **492** | Platelets | Platelet Count (PLT) \| 777-3 | 777-3 | LOINC | hematological |
| **493** | Platelets | CBC W/DIFF AND PLATELETS -Q\|PLATELET COUNT-Q\|777-3 | 26515-7 | LOINC | hematological |
| **494** | Platelets | COMPLETE BLOOD COUNT ONCOLOGY\|PLATELET COUNT\|85027 | 26515-7 | LOINC | hematological |
| **495** | Platelets | CBC,PLATELET, WITH DIFFERENTIAL\|PLATELET COUNT\|777-3 | 777-3 | LOINC | hematological |
| **496** | Platelets | CBC, PLATELET; NO DIFFL - LC\|PLATELETS\|85027 | 26515-7 | LOINC | hematological |
| **497** | Platelets | CBC, PLATELET, NO DIFFERENTIAL\|PLATELET COUNT\|777-3 | 777-3 | LOINC | hematological |
| **498** | Platelets | PLATELET COUNT\|777-3 | 26515-7 | LOINC | hematological |
| **499** | Platelets | PLATELET PROFILE\|PLATELET COUNT\|26515-7 | 26515-7 | LOINC | hematological |
| **500** | Platelets | PLATELET COUNT \| CBC WITH AUTODIFF \| 777-3 \| 12302 | 777-3 | LOINC | hematological |
| **501** | Platelets | CBC,PLATELET, WITH DIFFERENTIAL\|PLATELETS-LC\|777-3 | 26515-7 | LOINC | hematological |
| **502** | Platelets | PLATELET COUNT | 26515-7 | LOINC | hematological |
| **503** | Platelets | PLATELET COUNT \| CBC WITH NO DIFF \| 777-3 \| 123020 | 777-3 | LOINC | hematological |
| **504** | Platelets | CBC,PLATELET, WITH DIFFERENTIAL\|PLATELET COUNT-Q\|777-3 | 26515-7 | LOINC | hematological |
| **505** | Platelets | Platelet Count \| 777-3 | 777-3 | LOINC | hematological |
| **506** | Platelets | Platelet count \| 777-3 | 777-3 | LOINC | hematological |
| **507** | Platelets | PLATELET COUNT \| PLATELET COUNT - PERFORMABLE \| 77 | 777-3 | LOINC | hematological |
| **508** | Platelets | PLATELET COUNT \| CBC WITH NO DIFF \| 777-3 \| 24 | 777-3 | LOINC | hematological |
| **509** | Platelets | PLATELETS\|777-3 | 26515-7 | LOINC | hematological |
| **510** | Platelets | PLT\|777-3 | 26515-7 | LOINC | hematological |
| **511** | Platelets | DIFFERENTIAL, MANUAL -Q\|PLATELET ESTIMATION\|415.002 | 26515-7 | LOINC | hematological |
| **512** | Platelets | PLTS\|14869-2 | 26515-7 | LOINC | hematological |
| **513** | Platelets | PLATELET COUNT - UCH MN \| CBC WITH MANUAL DIFF - U | 777-3 | LOINC | hematological |
| **514** | Platelets | PLATELET COUNT \| PLATELET COUNT \| 777-3 \| 24 | 777-3 | LOINC | hematological |
| **515** | Platelets | PLATELET COUNT \| CBC AUTODIFF FOR SEPSIS \| 777-3 \| | 777-3 | LOINC | hematological |
| **516** | Platelets | COMPLETE BLOOD COUNT ONCOLOGY\|PLATELET ESTIMATE\|85027 | 26515-7 | LOINC | hematological |
| **517** | Platelets | PLATELET COUNT \| PLATELET COUNT \| 777-3 \| 12302000 | 777-3 | LOINC | hematological |
| **518** | Platelets | PLATELET COUNT (H/O)\|777-3 | 777-3 | LOINC | hematological |
| **519** | Platelets | CBC,PLATELET, WITH DIFFERENTIAL\|PLATELET COUNT\|26515-7 | 26515-7 | LOINC | hematological |

| | | | | | |
|---|---|---|---|---|---|
| **520** | Platelets | PLATELET COUNT \| CBC SEPSIS W/DIFF \| 777-3 \| 24 | 777-3 | LOINC | hematological |
| **521** | Platelets | PLATELETS, BLOOD (EXT RSLT)\|777-3 | 777-3 | LOINC | hematological |
| **522** | Platelets | CBC WITH DIFFERENTIAL/PLATELET - LC\|PLATELETS-LC\|777-3 | 26515-7 | LOINC | hematological |
| **523** | Platelets | CBC, PLATELET, NO DIFFERENTIAL\|PLATELET ESTIMATE\|85027 | 26515-7 | LOINC | hematological |
| **524** | Platelets | PLATELETS EXT\|777-3 | 777-3 | LOINC | hematological |
| **525** | Platelets | PLT, POC \|777-3 | 777-3 | LOINC | hematological |
| **526** | Platelets | CBC W/DIFF AND PLATELETS -Q\|PLATELET COUNT\|85025 | 26515-7 | LOINC | hematological |
| **527** | Platelets | CBC, PLATELET, NO DIFFERENTIAL\|PLATELET COUNT\|26515-7 | 26515-7 | LOINC | hematological |
| **528** | Platelets | CBC, PLATELET, NO DIFFERENTIAL\|PLATELETS-LC\|777-3 | 26515-7 | LOINC | hematological |
| **529** | Platelets | CBC,PLATELET, WITH DIFFERENTIAL\|PLATELET ESTIMATE\|85025 | 26515-7 | LOINC | hematological |
| **530** | Platelets | PLATELET COUNT - UCH MN \| PLATELET COUNT - UCH MN | 777-3 | LOINC | hematological |
| **531** | Platelets | CBC, PLATELET, NO DIFFERENTIAL\|PLATELET COUNT\|85027 | 26515-7 | LOINC | hematological |
| **532** | Platelets | PLATELET COUNT - NOC \| CBC WITH AUTO DIFF - NOC SY | 777-3 | LOINC | hematological |
| **533** | Platelets | PLATELET\|777-3 | 777-3 | LOINC | hematological |
| **534** | Platelets | CBC/DIFF AMBIGUOUS DEFAULT - LC\|PLATELETS-LC\|777-3 | 26515-7 | LOINC | hematological |
| **535** | Platelets | CBC, PLATELET, NO DIFFERENTIAL\|PLATELET COUNT-Q\|777-3 | 26515-7 | LOINC | hematological |
| **536** | Platelets | PLATELET PROFILE\|PLATELET COUNT\|806727 | 26515-7 | LOINC | hematological |
| **537** | Platelets | PLATELET EST/MORPH\|6927 | 26515-7 | LOINC | hematological |
| **538** | Platelets | PLATELET COUNT - UCH MN \| CBC SEPSIS (PERFORMABLE) | 777-3 | LOINC | hematological |
| **539** | Platelets | Platelet Count | 26515-7 | LOINC | hematological |
| **540** | Platelets | CBC,PLATELET, WITH DIFFERENTIAL\|PLATELET COUNT\|85025 | 26515-7 | LOINC | hematological |
| **541** | Platelets | PLATELET COUNT - OUTSIDE LAB \| OUTSIDE LAB-CBC \| 2 | 26515-7 | LOINC | hematological |
| **542** | Platelets | CBC (H&H, RBC, INDICES, WBC, PLT) - Q\|PLATELET COUNT\|85027 | 26515-7 | LOINC | hematological |
| **543** | Platelets | CBC W/DIFF AND PLATELETS -Q\|PLATELET COUNT-Q\|85025 | 26515-7 | LOINC | hematological |
| **544** | Platelets | MANUAL DIFFERENTIAL-WAM\|PLATELET ESTIMATE\|85007.001 | 26515-7 | LOINC | hematological |
| **545** | Platelets | PLATELETS\|777-3 | 777-3 | LOINC | hematological |
| **546** | Platelets | PLATELET COUNT - UCH MN \| CBC NO AUTO DIFF - UCH M | 777-3 | LOINC | hematological |
| **547** | thrombophlebitis and thromboembolism | Embolism and thrombosis of other parts of aorta | I74.19 | ICD10CM | hematological |

| | | | | | |
|---|---|---|---|---|---|
| **548** | thrombophlebitis and thromboembolism | Acute embolism and thrombosis of unspecified deep veins of distal lower extremity, bilateral | I82.4Z3 | ICD10CM | hematological |
| **549** | thrombophlebitis and thromboembolism | Chronic embolism and thrombosis of unspecified femoral vein | I82.519 | ICD10CM | hematological |
| **550** | thrombophlebitis and thromboembolism | Chronic embolism and thrombosis of unspecified deep veins of left lower extremity | I82.502 | ICD10CM | hematological |
| **551** | thrombophlebitis and thromboembolism | Chronic embolism and thrombosis of femoral vein, bilateral | I82.513 | ICD10CM | hematological |
| **552** | thrombophlebitis and thromboembolism | Phlebitis and thrombophlebitis of right tibial vein | I80.231 | ICD10CM | hematological |
| **553** | thrombophlebitis and thromboembolism | Acute embolism and thrombosis of superficial veins of left upper extremity | I82.612 | ICD10CM | hematological |
| **554** | thrombophlebitis and thromboembolism | Acute embolism and thrombosis of subclavian vein, bilateral | I82.B13 | ICD10CM | hematological |
| **555** | thrombophlebitis and thromboembolism | Deep phlebothrombosis in pregnancy, unspecified trimester | O22.30 | ICD10CM | hematological |
| **556** | thrombophlebitis and thromboembolism | Chronic embolism and thrombosis of other specified deep vein of unspecified lower extremity | I82.599 | ICD10CM | hematological |
| **557** | thrombophlebitis and thromboembolism | Chronic embolism and thrombosis of calf muscular vein, bilateral | I82.563 | ICD10CM | hematological |
| **558** | thrombophlebitis and thromboembolism | Acute embolism and thrombosis of unspecified veins of unspecified upper extremity | I82.609 | ICD10CM | hematological |
| **559** | thrombophlebitis and thromboembolism | Acute embolism and thrombosis of unspecified iliac vein | I82.429 | ICD10CM | hematological |
| **560** | thrombophlebitis and thromboembolism | Deep phlebothrombosis in pregnancy, first trimester | O22.31 | ICD10CM | hematological |
| **561** | thrombophlebitis and thromboembolism | Chronic embolism and thrombosis of unspecified veins of left upper extremity | I82.702 | ICD10CM | hematological |
| **562** | thrombophlebitis and thromboembolism | Acute embolism and thrombosis of internal jugular vein, bilateral | I82.C13 | ICD10CM | hematological |
| **563** | thrombophlebitis and thromboembolism | Chronic embolism and thrombosis of deep veins of unspecified upper extremity | I82.729 | ICD10CM | hematological |
| **564** | thrombophlebitis and thromboembolism | Acute embolism and thrombosis of unspecified deep veins of left proximal lower extremity | I82.4Y2 | ICD10CM | hematological |
| **565** | thrombophlebitis and thromboembolism | Chronic embolism and thrombosis of unspecified deep veins of unspecified proximal lower extremity | I82.5Y9 | ICD10CM | hematological |
| **566** | thrombophlebitis and thromboembolism | Atheroembolism of left lower extremity | I75.022 | ICD10CM | hematological |
| **567** | thrombophlebitis and thromboembolism | Acute embolism and thrombosis of deep veins of left upper extremity | I82.622 | ICD10CM | hematological |
| **568** | thrombophlebitis and thromboembolism | Chronic embolism and thrombosis of unspecified calf muscular vein | I82.569 | ICD10CM | hematological |
| **569** | thrombophlebitis and thromboembolism | Phlebitis and thrombophlebitis of left popliteal vein | I80.222 | ICD10CM | hematological |
| **570** | thrombophlebitis and thromboembolism | Chronic embolism and thrombosis of deep veins of upper extremity, bilateral | I82.723 | ICD10CM | hematological |
| **571** | thrombophlebitis and thromboembolism | Chronic embolism and thrombosis of right iliac vein | I82.521 | ICD10CM | hematological |
| **572** | thrombophlebitis and thromboembolism | Chronic embolism and thrombosis of unspecified deep veins of unspecified lower extremity | I82.509 | ICD10CM | hematological |

| | | | | | |
|---|---|---|---|---|---|
| **573** | thrombophlebitis and thromboembolism | Embolism and thrombosis of superficial veins of unspecified lower extremity | I82.819 | ICD10CM | hematological |
| **574** | thrombophlebitis and thromboembolism | Chronic embolism and thrombosis of unspecified tibial vein | I82.549 | ICD10CM | hematological |
| **575** | thrombophlebitis and thromboembolism | Superficial thrombophlebitis in pregnancy, unspecified trimester | O22.20 | ICD10CM | hematological |
| **576** | thrombophlebitis and thromboembolism | Phlebitis and thrombophlebitis of superficial vessels of lower extremities, bilateral | I80.03 | ICD10CM | hematological |
| **577** | thrombophlebitis and thromboembolism | Superficial thrombophlebitis in the puerperium | O87.0 | ICD10CM | hematological |
| **578** | thrombophlebitis and thromboembolism | Phlebitis and thrombophlebitis of right popliteal vein | I80.221 | ICD10CM | hematological |
| **579** | thrombophlebitis and thromboembolism | Chronic embolism and thrombosis of subclavian vein, bilateral | I82.B23 | ICD10CM | hematological |
| **580** | thrombophlebitis and thromboembolism | Phlebitis and thrombophlebitis of superficial vessels of right lower extremity | I80.01 | ICD10CM | hematological |
| **581** | thrombophlebitis and thromboembolism | Chronic embolism and thrombosis of left axillary vein | I82.A22 | ICD10CM | hematological |
| **582** | thrombophlebitis and thromboembolism | Septic arterial embolism | I76 | ICD10CM | hematological |
| **583** | thrombophlebitis and thromboembolism | Chronic embolism and thrombosis of right peroneal vein | I82.551 | ICD10CM | hematological |
| **584** | thrombophlebitis and thromboembolism | Acute embolism and thrombosis of femoral vein, bilateral | I82.413 | ICD10CM | hematological |
| **585** | thrombophlebitis and thromboembolism | Phlebitis and thrombophlebitis of unspecified deep vessels of right lower extremity | I80.201 | ICD10CM | hematological |
| **586** | thrombophlebitis and thromboembolism | Superficial thrombophlebitis in pregnancy, second trimester | O22.22 | ICD10CM | hematological |
| **587** | thrombophlebitis and thromboembolism | Phlebitis and thrombophlebitis of left calf muscular vein | I80.252 | ICD10CM | hematological |
| **588** | thrombophlebitis and thromboembolism | Thrombosis of atrium, auricular appendage, and ventricle as current complications following acute myocardial infarction | I23.6 | ICD10CM | hematological |
| **589** | thrombophlebitis and thromboembolism | Chronic embolism and thrombosis of left internal jugular vein | I82.C22 | ICD10CM | hematological |
| **590** | thrombophlebitis and thromboembolism | Budd-Chiari syndrome | I82.0 | ICD10CM | hematological |
| **591** | thrombophlebitis and thromboembolism | Other pulmonary embolism with acute cor pulmonale | I26.09 | ICD10CM | hematological |
| **592** | thrombophlebitis and thromboembolism | Acute embolism and thrombosis of other thoracic veins | I82.290 | ICD10CM | hematological |
| **593** | thrombophlebitis and thromboembolism | Embolism and thrombosis of renal vein | I82.3 | ICD10CM | hematological |
| **594** | thrombophlebitis and thromboembolism | Chronic embolism and thrombosis of unspecified deep veins of right distal lower extremity | I82.5Z1 | ICD10CM | hematological |
| **595** | thrombophlebitis and thromboembolism | Acute embolism and thrombosis of other specified deep vein of unspecified lower extremity | I82.499 | ICD10CM | hematological |
| **596** | thrombophlebitis and thromboembolism | Phlebitis and thrombophlebitis of unspecified tibial vein | I80.239 | ICD10CM | hematological |
| **597** | thrombophlebitis and thromboembolism | Acute embolism and thrombosis of right femoral vein | I82.411 | ICD10CM | hematological |

| | | | | | |
|---|---|---|---|---|---|
| **598** | thrombophlebitis and thromboembolism | Acute embolism and thrombosis of unspecified deep veins of right proximal lower extremity | I82.4Y1 | ICD10CM | hematological |
| **599** | thrombophlebitis and thromboembolism | Atheroembolism of other site | I75.89 | ICD10CM | hematological |
| **600** | thrombophlebitis and thromboembolism | Chronic embolism and thrombosis of unspecified deep veins of distal lower extremity, bilateral | I82.5Z3 | ICD10CM | hematological |
| **601** | thrombophlebitis and thromboembolism | Phlebitis and thrombophlebitis of left tibial vein | I80.232 | ICD10CM | hematological |
| **602** | thrombophlebitis and thromboembolism | Acute embolism and thrombosis of unspecified peroneal vein | I82.459 | ICD10CM | hematological |
| **603** | thrombophlebitis and thromboembolism | Atheroembolism of bilateral upper extremities | I75.013 | ICD10CM | hematological |
| **604** | thrombophlebitis and thromboembolism | Acute embolism and thrombosis of superior vena cava | I82.210 | ICD10CM | hematological |
| **605** | thrombophlebitis and thromboembolism | Atheroembolism of right lower extremity | I75.021 | ICD10CM | hematological |
| **606** | thrombophlebitis and thromboembolism | Chronic embolism and thrombosis of left calf muscular vein | I82.562 | ICD10CM | hematological |
| **607** | thrombophlebitis and thromboembolism | Acute embolism and thrombosis of unspecified tibial vein | I82.449 | ICD10CM | hematological |
| **608** | thrombophlebitis and thromboembolism | Acute embolism and thrombosis of left axillary vein | I82.A12 | ICD10CM | hematological |
| **609** | thrombophlebitis and thromboembolism | Acute embolism and thrombosis of right axillary vein | I82.A11 | ICD10CM | hematological |
| **610** | thrombophlebitis and thromboembolism | Chronic embolism and thrombosis of other specified deep vein of right lower extremity | I82.591 | ICD10CM | hematological |
| **611** | thrombophlebitis and thromboembolism | Atheroembolism of left upper extremity | I75.012 | ICD10CM | hematological |
| **612** | thrombophlebitis and thromboembolism | Phlebitis and thrombophlebitis of tibial vein, bilateral | I80.233 | ICD10CM | hematological |
| **613** | thrombophlebitis and thromboembolism | Chronic embolism and thrombosis of other thoracic veins | I82.291 | ICD10CM | hematological |
| **614** | thrombophlebitis and thromboembolism | Other pulmonary embolism without acute cor pulmonale | I26.99 | ICD10CM | hematological |
| **615** | thrombophlebitis and thromboembolism | Embolism and thrombosis of superficial veins of right lower extremity | I82.811 | ICD10CM | hematological |
| **616** | thrombophlebitis and thromboembolism | Acute embolism and thrombosis of unspecified deep veins of unspecified lower extremity | I82.409 | ICD10CM | hematological |
| **617** | thrombophlebitis and thromboembolism | Embolism and thrombosis of superficial veins of left lower extremity | I82.812 | ICD10CM | hematological |
| **618** | thrombophlebitis and thromboembolism | Chronic embolism and thrombosis of unspecified deep veins of unspecified distal lower extremity | I82.5Z9 | ICD10CM | hematological |
| **619** | thrombophlebitis and thromboembolism | Acute embolism and thrombosis of axillary vein, bilateral | I82.A13 | ICD10CM | hematological |
| **620** | thrombophlebitis and thromboembolism | Saddle embolus of pulmonary artery with acute cor pulmonale | I26.02 | ICD10CM | hematological |
| **621** | thrombophlebitis and thromboembolism | Chronic embolism and thrombosis of unspecified peroneal vein | I82.559 | ICD10CM | hematological |
| **622** | thrombophlebitis and thromboembolism | Embolism and thrombosis of arteries of extremities, unspecified | I74.4 | ICD10CM | hematological |

| | | | | | |
|---|---|---|---|---|---|
| **623** | thrombophlebitis and thromboembolism | Phlebitis and thrombophlebitis of unspecified deep vessels of lower extremities, bilateral | I80.203 | ICD10CM | hematological |
| **624** | thrombophlebitis and thromboembolism | Pulmonary embolism with acute cor pulmonale | I26.0 | ICD10CM | hematological |
| **625** | thrombophlebitis and thromboembolism | Chronic embolism and thrombosis of left popliteal vein | I82.532 | ICD10CM | hematological |
| **626** | thrombophlebitis and thromboembolism | Deep phlebothrombosis in the puerperium | O87.1 | ICD10CM | hematological |
| **627** | thrombophlebitis and thromboembolism | Atheroembolism of kidney | I75.81 | ICD10CM | hematological |
| **628** | thrombophlebitis and thromboembolism | Embolism and thrombosis of unspecified parts of aorta | I74.10 | ICD10CM | hematological |
| **629** | thrombophlebitis and thromboembolism | Acute embolism and thrombosis of other specified veins | I82.890 | ICD10CM | hematological |
| **630** | thrombophlebitis and thromboembolism | Acute embolism and thrombosis of peroneal vein, bilateral | I82.453 | ICD10CM | hematological |
| **631** | thrombophlebitis and thromboembolism | Chronic embolism and thrombosis of peroneal vein, bilateral | I82.553 | ICD10CM | hematological |
| **632** | thrombophlebitis and thromboembolism | Chronic embolism and thrombosis of other specified veins | I82.891 | ICD10CM | hematological |
| **633** | thrombophlebitis and thromboembolism | Phlebitis and thrombophlebitis of popliteal vein, bilateral | I80.223 | ICD10CM | hematological |
| **634** | thrombophlebitis and thromboembolism | Embolism and thrombosis of abdominal aorta | I74.0 | ICD10CM | hematological |
| **635** | thrombophlebitis and thromboembolism | Chronic embolism and thrombosis of left iliac vein | I82.522 | ICD10CM | hematological |
| **636** | thrombophlebitis and thromboembolism | Chronic embolism and thrombosis of unspecified deep veins of lower extremity, bilateral | I82.503 | ICD10CM | hematological |
| **637** | thrombophlebitis and thromboembolism | Acute embolism and thrombosis of calf muscular vein, bilateral | I82.463 | ICD10CM | hematological |
| **638** | thrombophlebitis and thromboembolism | Chronic embolism and thrombosis of left femoral vein | I82.512 | ICD10CM | hematological |
| **639** | thrombophlebitis and thromboembolism | Acute embolism and thrombosis of iliac vein, bilateral | I82.423 | ICD10CM | hematological |
| **640** | thrombophlebitis and thromboembolism | Single subsegmental pulmonary embolism without acute cor pulmonale | I26.93 | ICD10CM | hematological |
| **641** | thrombophlebitis and thromboembolism | Chronic embolism and thrombosis of inferior vena cava | I82.221 | ICD10CM | hematological |
| **642** | thrombophlebitis and thromboembolism | Acute embolism and thrombosis of unspecified deep veins of lower extremity, bilateral | I82.403 | ICD10CM | hematological |
| **643** | thrombophlebitis and thromboembolism | Embolism and thrombosis of iliac artery | I74.5 | ICD10CM | hematological |
| **644** | thrombophlebitis and thromboembolism | Chronic embolism and thrombosis of superior vena cava | I82.211 | ICD10CM | hematological |
| **645** | thrombophlebitis and thromboembolism | Atheroembolism of unspecified lower extremity | I75.029 | ICD10CM | hematological |
| **646** | thrombophlebitis and thromboembolism | Embolism and thrombosis of other arteries | I74.8 | ICD10CM | hematological |
| **647** | thrombophlebitis and thromboembolism | Phlebitis and thrombophlebitis of unspecified site | I80.9 | ICD10CM | hematological |

| | | | | | |
|---|---|---|---|---|---|
| **648** | thrombophlebitis and thromboembolism | Phlebitis and thrombophlebitis of calf muscular vein, bilateral | I80.253 | ICD10CM | hematological |
| **649** | thrombophlebitis and thromboembolism | Acute embolism and thrombosis of right calf muscular vein | I82.461 | ICD10CM | hematological |
| **650** | thrombophlebitis and thromboembolism | Septic pulmonary embolism without acute cor pulmonale | I26.90 | ICD10CM | hematological |
| **651** | thrombophlebitis and thromboembolism | Phlebitis and thrombophlebitis of left iliac vein | I80.212 | ICD10CM | hematological |
| **652** | thrombophlebitis and thromboembolism | Acute embolism and thrombosis of left subclavian vein | I82.B12 | ICD10CM | hematological |
| **653** | thrombophlebitis and thromboembolism | Acute embolism and thrombosis of other specified deep vein of left lower extremity | I82.492 | ICD10CM | hematological |
| **654** | thrombophlebitis and thromboembolism | Phlebitis and thrombophlebitis of unspecified popliteal vein | I80.229 | ICD10CM | hematological |
| **655** | thrombophlebitis and thromboembolism | Chronic embolism and thrombosis of left peroneal vein | I82.552 | ICD10CM | hematological |
| **656** | thrombophlebitis and thromboembolism | Acute embolism and thrombosis of other specified deep vein of lower extremity, bilateral | I82.493 | ICD10CM | hematological |
| **657** | thrombophlebitis and thromboembolism | Phlebitis and thrombophlebitis of left femoral vein | I80.12 | ICD10CM | hematological |
| **658** | thrombophlebitis and thromboembolism | Acute embolism and thrombosis of left calf muscular vein | I82.462 | ICD10CM | hematological |
| **659** | thrombophlebitis and thromboembolism | Chronic embolism and thrombosis of unspecified veins of unspecified upper extremity | I82.709 | ICD10CM | hematological |
| **660** | thrombophlebitis and thromboembolism | Phlebitis and thrombophlebitis of unspecified iliac vein | I80.219 | ICD10CM | hematological |
| **661** | thrombophlebitis and thromboembolism | Saddle embolus of pulmonary artery without acute cor pulmonale | I26.92 | ICD10CM | hematological |
| **662** | thrombophlebitis and thromboembolism | Superficial thrombophlebitis in pregnancy, first trimester | O22.21 | ICD10CM | hematological |
| **663** | thrombophlebitis and thromboembolism | Phlebitis and thrombophlebitis of superficial vessels of unspecified lower extremity | I80.00 | ICD10CM | hematological |
| **664** | thrombophlebitis and thromboembolism | Acute embolism and thrombosis of left femoral vein | I82.412 | ICD10CM | hematological |
| **665** | thrombophlebitis and thromboembolism | Acute embolism and thrombosis of unspecified deep veins of proximal lower extremity, bilateral | I82.4Y3 | ICD10CM | hematological |
| **666** | thrombophlebitis and thromboembolism | Phlebitis and thrombophlebitis of femoral vein, bilateral | I80.13 | ICD10CM | hematological |
| **667** | thrombophlebitis and thromboembolism | Chronic embolism and thrombosis of unspecified axillary vein | I82.A29 | ICD10CM | hematological |
| **668** | thrombophlebitis and thromboembolism | Phlebitis and thrombophlebitis of other deep vessels of lower extremity, bilateral | I80.293 | ICD10CM | hematological |
| **669** | thrombophlebitis and thromboembolism | Chronic embolism and thrombosis of axillary vein, bilateral | I82.A23 | ICD10CM | hematological |
| **670** | thrombophlebitis and thromboembolism | Chronic embolism and thrombosis of internal jugular vein, bilateral | I82.C23 | ICD10CM | hematological |
| **671** | thrombophlebitis and thromboembolism | Embolism and thrombosis of superficial veins of lower extremities, bilateral | I82.813 | ICD10CM | hematological |
| **672** | thrombophlebitis and thromboembolism | Phlebitis and thrombophlebitis of lower extremities, unspecified | I80.3 | ICD10CM | hematological |

| | | | | | |
|---|---|---|---|---|---|
| **673** | thrombophlebitis and thromboembolism | Chronic embolism and thrombosis of other specified deep vein of left lower extremity | I82.592 | ICD10CM | hematological |
| **674** | thrombophlebitis and thromboembolism | Acute embolism and thrombosis of unspecified deep veins of unspecified proximal lower extremity | I82.4Y9 | ICD10CM | hematological |
| **675** | thrombophlebitis and thromboembolism | Acute embolism and thrombosis of left peroneal vein | I82.452 | ICD10CM | hematological |
| **676** | thrombophlebitis and thromboembolism | Embolism and thrombosis of unspecified artery | I74.9 | ICD10CM | hematological |
| **677** | thrombophlebitis and thromboembolism | Septic pulmonary embolism with acute cor pulmonale | I26.01 | ICD10CM | hematological |
| **678** | thrombophlebitis and thromboembolism | Chronic pulmonary embolism | I27.82 | ICD10CM | hematological |
| **679** | thrombophlebitis and thromboembolism | Phlebitis and thrombophlebitis of right femoral vein | I80.11 | ICD10CM | hematological |
| **680** | thrombophlebitis and thromboembolism | Acute embolism and thrombosis of left iliac vein | I82.422 | ICD10CM | hematological |
| **681** | thrombophlebitis and thromboembolism | Acute embolism and thrombosis of unspecified deep veins of left lower extremity | I82.402 | ICD10CM | hematological |
| **682** | thrombophlebitis and thromboembolism | Acute embolism and thrombosis of right iliac vein | I82.421 | ICD10CM | hematological |
| **683** | thrombophlebitis and thromboembolism | Acute embolism and thrombosis of unspecified deep veins of unspecified distal lower extremity | I82.4Z9 | ICD10CM | hematological |
| **684** | thrombophlebitis and thromboembolism | Other arterial embolism and thrombosis of abdominal aorta | I74.09 | ICD10CM | hematological |
| **685** | thrombophlebitis and thromboembolism | Acute embolism and thrombosis of right popliteal vein | I82.431 | ICD10CM | hematological |
| **686** | thrombophlebitis and thromboembolism | Phlebitis and thrombophlebitis of other deep vessels of unspecified lower extremity | I80.299 | ICD10CM | hematological |
| **687** | thrombophlebitis and thromboembolism | Phlebitis and thrombophlebitis of right peroneal vein | I80.241 | ICD10CM | hematological |
| **688** | thrombophlebitis and thromboembolism | Acute embolism and thrombosis of unspecified subclavian vein | I82.B19 | ICD10CM | hematological |
| **689** | thrombophlebitis and thromboembolism | Atheroembolism of bilateral lower extremities | I75.023 | ICD10CM | hematological |
| **690** | thrombophlebitis and thromboembolism | Acute embolism and thrombosis of unspecified axillary vein | I82.A19 | ICD10CM | hematological |
| **691** | thrombophlebitis and thromboembolism | Superficial thrombophlebitis in pregnancy, third trimester | O22.23 | ICD10CM | hematological |
| **692** | thrombophlebitis and thromboembolism | Acute embolism and thrombosis of unspecified veins of upper extremity, bilateral | I82.603 | ICD10CM | hematological |
| **693** | thrombophlebitis and thromboembolism | Chronic embolism and thrombosis of left tibial vein | I82.542 | ICD10CM | hematological |
| **694** | thrombophlebitis and thromboembolism | Phlebitis and thrombophlebitis of right calf muscular vein | I80.251 | ICD10CM | hematological |
| **695** | thrombophlebitis and thromboembolism | Acute embolism and thrombosis of left internal jugular vein | I82.C12 | ICD10CM | hematological |
| **696** | thrombophlebitis and thromboembolism | Chronic embolism and thrombosis of popliteal vein, bilateral | I82.533 | ICD10CM | hematological |
| **697** | thrombophlebitis and thromboembolism | Acute embolism and thrombosis of other specified deep vein of right lower extremity | I82.491 | ICD10CM | hematological |

| | | | | | |
|---|---|---|---|---|---|
| **698** | thrombophlebitis and thromboembolism | Acute embolism and thrombosis of inferior vena cava | I82.220 | ICD10CM | hematological |
| **699** | thrombophlebitis and thromboembolism | Pulmonary embolism without acute cor pulmonale | I26.9 | ICD10CM | hematological |
| **700** | thrombophlebitis and thromboembolism | Embolism and thrombosis of thoracic aorta | I74.11 | ICD10CM | hematological |
| **701** | thrombophlebitis and thromboembolism | Deep phlebothrombosis in pregnancy, second trimester | O22.32 | ICD10CM | hematological |
| **702** | thrombophlebitis and thromboembolism | Chronic embolism and thrombosis of right internal jugular vein | I82.C21 | ICD10CM | hematological |
| **703** | thrombophlebitis and thromboembolism | Atheroembolism of unspecified upper extremity | I75.019 | ICD10CM | hematological |
| **704** | thrombophlebitis and thromboembolism | Chronic embolism and thrombosis of right tibial vein | I82.541 | ICD10CM | hematological |
| **705** | thrombophlebitis and thromboembolism | Arterial embolism and thrombosis | I74 | ICD10CM | hematological |
| **706** | thrombophlebitis and thromboembolism | Deep phlebothrombosis in pregnancy, third trimester | O22.33 | ICD10CM | hematological |
| **707** | thrombophlebitis and thromboembolism | Acute embolism and thrombosis of unspecified deep veins of right distal lower extremity | I82.4Z1 | ICD10CM | hematological |
| **708** | thrombophlebitis and thromboembolism | Portal vein thrombosis | I81 | ICD10CM | hematological |
| **709** | thrombophlebitis and thromboembolism | Chronic embolism and thrombosis of unspecified veins of upper extremity, bilateral | I82.703 | ICD10CM | hematological |
| **710** | thrombophlebitis and thromboembolism | Embolism and thrombosis of arteries of the lower extremities | I74.3 | ICD10CM | hematological |
| **711** | thrombophlebitis and thromboembolism | Chronic embolism and thrombosis of unspecified vein | I82.91 | ICD10CM | hematological |
| **712** | thrombophlebitis and thromboembolism | Acute embolism and thrombosis of deep veins of unspecified upper extremity | I82.629 | ICD10CM | hematological |
| **713** | thrombophlebitis and thromboembolism | Acute embolism and thrombosis of superficial veins of right upper extremity | I82.611 | ICD10CM | hematological |
| **714** | thrombophlebitis and thromboembolism | Chronic embolism and thrombosis of unspecified deep veins of right lower extremity | I82.501 | ICD10CM | hematological |
| **715** | thrombophlebitis and thromboembolism | Acute embolism and thrombosis of unspecified calf muscular vein | I82.469 | ICD10CM | hematological |
| **716** | thrombophlebitis and thromboembolism | Acute embolism and thrombosis of unspecified deep veins of right lower extremity | I82.401 | ICD10CM | hematological |
| **717** | thrombophlebitis and thromboembolism | Chronic embolism and thrombosis of right femoral vein | I82.511 | ICD10CM | hematological |
| **718** | thrombophlebitis and thromboembolism | Acute embolism and thrombosis of unspecified veins of left upper extremity | I82.602 | ICD10CM | hematological |
| **719** | thrombophlebitis and thromboembolism | Chronic embolism and thrombosis of right axillary vein | I82.A21 | ICD10CM | hematological |
| **720** | thrombophlebitis and thromboembolism | Phlebitis and thrombophlebitis of right iliac vein | I80.211 | ICD10CM | hematological |
| **721** | thrombophlebitis and thromboembolism | Phlebitis and thrombophlebitis of left peroneal vein | I80.242 | ICD10CM | hematological |
| **722** | thrombophlebitis and thromboembolism | Chronic embolism and thrombosis of other specified deep vein of lower extremity, bilateral | I82.593 | ICD10CM | hematological |

| | | | | | |
|---|---|---|---|---|---|
| **723** | thrombophlebitis and thromboembolism | Chronic embolism and thrombosis of superficial veins of upper extremity, bilateral | I82.713 | ICD10CM | hematological |
| **724** | thrombophlebitis and thromboembolism | Phlebitis and thrombophlebitis of unspecified deep vessels of left lower extremity | I80.202 | ICD10CM | hematological |
| **725** | thrombophlebitis and thromboembolism | Chronic embolism and thrombosis of unspecified internal jugular vein | I82.C29 | ICD10CM | hematological |
| **726** | thrombophlebitis and thromboembolism | Phlebitis and thrombophlebitis of unspecified deep vessels of unspecified lower extremity | I80.209 | ICD10CM | hematological |
| **727** | thrombophlebitis and thromboembolism | Chronic embolism and thrombosis of superficial veins of right upper extremity | I82.711 | ICD10CM | hematological |
| **728** | thrombophlebitis and thromboembolism | Intracardiac thrombosis, not elsewhere classified | I51.3 | ICD10CM | hematological |
| **729** | thrombophlebitis and thromboembolism | Acute embolism and thrombosis of deep veins of right upper extremity | I82.621 | ICD10CM | hematological |
| **730** | thrombophlebitis and thromboembolism | Chronic embolism and thrombosis of superficial veins of left upper extremity | I82.712 | ICD10CM | hematological |
| **731** | thrombophlebitis and thromboembolism | Thrombophlebitis migrans | I82.1 | ICD10CM | hematological |
| **732** | thrombophlebitis and thromboembolism | Phlebitis and thrombophlebitis of unspecified peroneal vein | I80.249 | ICD10CM | hematological |
| **733** | thrombophlebitis and thromboembolism | Phlebitis and thrombophlebitis of other deep vessels of left lower extremity | I80.292 | ICD10CM | hematological |
| **734** | thrombophlebitis and thromboembolism | Acute embolism and thrombosis of superficial veins of upper extremity, bilateral | I82.613 | ICD10CM | hematological |
| **735** | thrombophlebitis and thromboembolism | Phlebitis and thrombophlebitis of unspecified femoral vein | I80.10 | ICD10CM | hematological |
| **736** | thrombophlebitis and thromboembolism | Chronic embolism and thrombosis of unspecified deep veins of left proximal lower extremity | I82.5Y2 | ICD10CM | hematological |
| **737** | thrombophlebitis and thromboembolism | Saddle embolus of abdominal aorta | I74.01 | ICD10CM | hematological |
| **738** | thrombophlebitis and thromboembolism | Acute embolism and thrombosis of deep veins of upper extremity, bilateral | I82.623 | ICD10CM | hematological |
| **739** | thrombophlebitis and thromboembolism | Acute embolism and thrombosis of right internal jugular vein | I82.C11 | ICD10CM | hematological |
| **740** | thrombophlebitis and thromboembolism | Chronic embolism and thrombosis of superficial veins of unspecified upper extremity | I82.719 | ICD10CM | hematological |
| **741** | thrombophlebitis and thromboembolism | Chronic embolism and thrombosis of right popliteal vein | I82.531 | ICD10CM | hematological |
| **742** | thrombophlebitis and thromboembolism | Chronic embolism and thrombosis of right calf muscular vein | I82.561 | ICD10CM | hematological |
| **743** | thrombophlebitis and thromboembolism | Acute embolism and thrombosis of right subclavian vein | I82.B11 | ICD10CM | hematological |
| **744** | thrombophlebitis and thromboembolism | Embolism and thrombosis of arteries of the upper extremities | I74.2 | ICD10CM | hematological |
| **745** | thrombophlebitis and thromboembolism | Chronic embolism and thrombosis of unspecified veins of right upper extremity | I82.701 | ICD10CM | hematological |
| **746** | thrombophlebitis and thromboembolism | Phlebitis and thrombophlebitis of iliac vein, bilateral | I80.213 | ICD10CM | hematological |
| **747** | thrombophlebitis and thromboembolism | Acute embolism and thrombosis of superficial veins of unspecified upper extremity | I82.619 | ICD10CM | hematological |

| | | | | | |
|---|---|---|---|---|---|
| **748** | thrombophlebitis and thromboembolism | Phlebitis and thrombophlebitis of peroneal vein, bilateral | I80.243 | ICD10CM | hematological |
| **749** | thrombophlebitis and thromboembolism | Atheroembolism of right upper extremity | I75.011 | ICD10CM | hematological |
| **750** | thrombophlebitis and thromboembolism | Acute embolism and thrombosis of left popliteal vein | I82.432 | ICD10CM | hematological |
| **751** | thrombophlebitis and thromboembolism | Chronic embolism and thrombosis of unspecified iliac vein | I82.529 | ICD10CM | hematological |
| **752** | thrombophlebitis and thromboembolism | Chronic embolism and thrombosis of tibial vein, bilateral | I82.543 | ICD10CM | hematological |
| **753** | thrombophlebitis and thromboembolism | Chronic embolism and thrombosis of deep veins of right upper extremity | I82.721 | ICD10CM | hematological |
| **754** | thrombophlebitis and thromboembolism | Acute embolism and thrombosis of left tibial vein | I82.442 | ICD10CM | hematological |
| **755** | thrombophlebitis and thromboembolism | Phlebitis and thrombophlebitis of superficial vessels of left lower extremity | I80.02 | ICD10CM | hematological |
| **756** | thrombophlebitis and thromboembolism | Chronic embolism and thrombosis of right subclavian vein | I82.B21 | ICD10CM | hematological |
| **757** | thrombophlebitis and thromboembolism | Acute embolism and thrombosis of tibial vein, bilateral | I82.443 | ICD10CM | hematological |
| **758** | thrombophlebitis and thromboembolism | Embolism and thrombosis of other and unspecified parts of aorta | I74.1 | ICD10CM | hematological |
| **759** | thrombophlebitis and thromboembolism | Phlebitis and thrombophlebitis of other sites | I80.8 | ICD10CM | hematological |
| **760** | thrombophlebitis and thromboembolism | Acute embolism and thrombosis of unspecified deep veins of left distal lower extremity | I82.4Z2 | ICD10CM | hematological |
| **761** | thrombophlebitis and thromboembolism | Puerperal septic thrombophlebitis | O86.81 | ICD10CM | hematological |
| **762** | thrombophlebitis and thromboembolism | Acute embolism and thrombosis of right peroneal vein | I82.451 | ICD10CM | hematological |
| **763** | thrombophlebitis and thromboembolism | Chronic embolism and thrombosis of unspecified deep veins of left distal lower extremity | I82.5Z2 | ICD10CM | hematological |
| **764** | thrombophlebitis and thromboembolism | Acute embolism and thrombosis of unspecified femoral vein | I82.419 | ICD10CM | hematological |
| **765** | thrombophlebitis and thromboembolism | Acute embolism and thrombosis of unspecified veins of right upper extremity | I82.601 | ICD10CM | hematological |
| **766** | thrombophlebitis and thromboembolism | Chronic embolism and thrombosis of unspecified deep veins of proximal lower extremity, bilateral | I82.5Y3 | ICD10CM | hematological |
| **767** | thrombophlebitis and thromboembolism | Acute embolism and thrombosis of popliteal vein, bilateral | I82.433 | ICD10CM | hematological |
| **768** | thrombophlebitis and thromboembolism | Chronic embolism and thrombosis of left subclavian vein | I82.B22 | ICD10CM | hematological |
| **769** | thrombophlebitis and thromboembolism | Multiple subsegmental pulmonary emboli without acute cor pulmonale | I26.94 | ICD10CM | hematological |
| **770** | thrombophlebitis and thromboembolism | Chronic embolism and thrombosis of deep veins of left upper extremity | I82.722 | ICD10CM | hematological |
| **771** | thrombophlebitis and thromboembolism | Chronic embolism and thrombosis of unspecified subclavian vein | I82.B29 | ICD10CM | hematological |
| **772** | thrombophlebitis and thromboembolism | Acute embolism and thrombosis of unspecified popliteal vein | I82.439 | ICD10CM | hematological |

| | | | | | |
|---|---|---|---|---|---|
| **773** | thrombophlebitis and thromboembolism | Chronic embolism and thrombosis of unspecified popliteal vein | I82.539 | ICD10CM | hematological |
| **774** | thrombophlebitis and thromboembolism | Chronic embolism and thrombosis of iliac vein, bilateral | I82.523 | ICD10CM | hematological |
| **775** | thrombophlebitis and thromboembolism | Acute embolism and thrombosis of unspecified internal jugular vein | I82.C19 | ICD10CM | hematological |
| **776** | thrombophlebitis and thromboembolism | Phlebitis and thrombophlebitis of unspecified calf muscular vein | I80.259 | ICD10CM | hematological |
| **777** | thrombophlebitis and thromboembolism | Phlebitis and thrombophlebitis of other deep vessels of right lower extremity | I80.291 | ICD10CM | hematological |
| **778** | thrombophlebitis and thromboembolism | Pulmonary embolism | I26 | ICD10CM | hematological |
| **779** | thrombophlebitis and thromboembolism | Acute embolism and thrombosis of unspecified vein | I82.90 | ICD10CM | hematological |
| **780** | thrombophlebitis and thromboembolism | Chronic embolism and thrombosis of unspecified deep veins of right proximal lower extremity | I82.5Y1 | ICD10CM | hematological |
| **781** | thrombophlebitis and thromboembolism | Acute embolism and thrombosis of right tibial vein | I82.441 | ICD10CM | hematological |
| **782** | cognitive signs and symptoms | Visual agnosia | R48.3 | ICD10CM | neurological |
| **783** | cognitive signs and symptoms | Agnosia | R48.1 | ICD10CM | neurological |
| **784** | cognitive signs and symptoms | Apraxia | R48.2 | ICD10CM | neurological |
| **785** | cognitive signs and symptoms | Unspecified symbolic dysfunctions | R48.9 | ICD10CM | neurological |
| **786** | cognitive signs and symptoms | Unspecified symptoms and signs involving general sensations and perceptions | R44.9 | ICD10CM | neurological |
| **787** | cognitive signs and symptoms | Other symbolic dysfunctions | R48.8 | ICD10CM | neurological |
| **788** | cognitive signs and symptoms | Other symptoms and signs involving general sensations and perceptions | R44.8 | ICD10CM | neurological |
| **789** | delirium | Opioid related disorders | F11 | ICD10CM | neurological |
| **790** | delirium | Other psychoactive substance related disorders | F19 | ICD10CM | neurological |
| **791** | delirium | Hallucinogen related disorders | F16 | ICD10CM | neurological |
| **792** | delirium | Inhalant related disorders | F18 | ICD10CM | neurological |
| **793** | delirium | Cannabis related disorders | F12 | ICD10CM | neurological |
| **794** | delirium | Amnestic disorder due to known physiological condition | F04 | ICD10CM | neurological |
| **795** | delirium | Other stimulant related disorders | F15 | ICD10CM | neurological |
| **796** | delirium | Alcohol related disorders | F10 | ICD10CM | neurological |
| **797** | delirium | Stupor | R40.1 | ICD10CM | neurological |
| **798** | delirium | Delirium due to known physiological condition | F05 | ICD10CM | neurological |
| **799** | delirium | Somnolence, stupor and coma | R40 | ICD10CM | neurological |
| **800** | delirium | Sedative, hypnotic, or anxiolytic related disorders | F13 | ICD10CM | neurological |
| **801** | delirium | Cocaine related disorders | F14 | ICD10CM | neurological |
| **802** | encephalopathy | Zoster encephalitis | B02.0 | ICD10CM | neurological |

| | | | | | |
|---|---|---|---|---|---|
| **803** | encephalopathy | Other myelitis | G04.89 | ICD10CM | neurological |
| **804** | encephalopathy | Mumps encephalitis | B26.2 | ICD10CM | neurological |
| **805** | encephalopathy | Acute disseminated encephalitis and encephalomyelitis (ADEM) | G04.0 | ICD10CM | neurological |
| **806** | encephalopathy | Other acute necrotizing hemorrhagic encephalopathy | G04.39 | ICD10CM | neurological |
| **807** | encephalopathy | Central European tick-borne encephalitis | A84.1 | ICD10CM | neurological |
| **808** | encephalopathy | Postinfectious acute disseminated encephalitis and encephalomyelitis (postinfectious ADEM) | G04.01 | ICD10CM | neurological |
| **809** | encephalopathy | Enteroviral encephalitis | A85.0 | ICD10CM | neurological |
| **810** | encephalopathy | Unspecified viral encephalitis | A86 | ICD10CM | neurological |
| **811** | encephalopathy | Australian encephalitis | A83.4 | ICD10CM | neurological |
| **812** | encephalopathy | Japanese encephalitis | A83.0 | ICD10CM | neurological |
| **813** | encephalopathy | Other specified mental disorders due to known physiological condition | F06.8 | ICD10CM | neurological |
| **814** | encephalopathy | Encephalopathy, unspecified | G93.40 | ICD10CM | neurological |
| **815** | encephalopathy | Other mosquito-borne viral encephalitis | A83.8 | ICD10CM | neurological |
| **816** | encephalopathy | Other specified viral encephalitis | A85.8 | ICD10CM | neurological |
| **817** | encephalopathy | Postimmunization acute necrotizing hemorrhagic encephalopathy | G04.32 | ICD10CM | neurological |
| **818** | encephalopathy | Encephalitis and encephalomyelitis, unspecified | G04.90 | ICD10CM | neurological |
| **819** | encephalopathy | Tick-borne viral encephalitis, unspecified | A84.9 | ICD10CM | neurological |
| **820** | encephalopathy | Other tick-borne viral encephalitis | A84.8 | ICD10CM | neurological |
| **821** | encephalopathy | Far Eastern tick-borne encephalitis [Russian spring-summer encephalitis] | A84.0 | ICD10CM | neurological |
| **822** | encephalopathy | Tick-borne viral encephalitis | A84 | ICD10CM | neurological |
| **823** | encephalopathy | Postimmunization acute disseminated encephalitis, myelitis and encephalomyelitis | G04.02 | ICD10CM | neurological |
| **824** | encephalopathy | Other and unspecified encephalopathy | G93.4 | ICD10CM | neurological |
| **825** | encephalopathy | Other encephalitis and encephalomyelitis | G04.81 | ICD10CM | neurological |
| **826** | encephalopathy | Herpesviral encephalitis | B00.4 | ICD10CM | neurological |
| **827** | encephalopathy | Mosquito-borne viral encephalitis | A83 | ICD10CM | neurological |
| **828** | encephalopathy | Varicella encephalitis, myelitis and encephalomyelitis | B01.1 | ICD10CM | neurological |
| **829** | encephalopathy | California encephalitis | A83.5 | ICD10CM | neurological |
| **830** | encephalopathy | Other viral encephalitis, not elsewhere classified | A85 | ICD10CM | neurological |
| **831** | encephalopathy | Postinfectious acute necrotizing hemorrhagic encephalopathy | G04.31 | ICD10CM | neurological |
| **832** | encephalopathy | Unspecified mental disorder due to known physiological condition | F06.9 | ICD10CM | neurological |
| **833** | encephalopathy | Myelitis, unspecified | G04.91 | ICD10CM | neurological |
| **834** | encephalopathy | Subacute sclerosing panencephalitis | A81.1 | ICD10CM | neurological |
| **835** | encephalopathy | Other encephalopathy | G93.49 | ICD10CM | neurological |
| **836** | encephalopathy | Adenoviral encephalitis | A85.1 | ICD10CM | neurological |

| | | | | | |
|---|---|---|---|---|---|
| **837** | encephalopathy | Mosquito-borne viral encephalitis, unspecified | A83.9 | ICD10CM | neurological |
| **838** | encephalopathy | Acute necrotizing hemorrhagic encephalopathy, unspecified | G04.30 | ICD10CM | neurological |
| **839** | encephalopathy | Toxoplasma meningoencephalitis | B58.2 | ICD10CM | neurological |
| **840** | encephalopathy | Measles complicated by encephalitis | B05.0 | ICD10CM | neurological |
| **841** | encephalopathy | Metabolic encephalopathy | G93.41 | ICD10CM | neurological |
| **842** | encephalopathy | Acute disseminated encephalitis and encephalomyelitis, unspecified | G04.00 | ICD10CM | neurological |
| **843** | encephalopathy | Bacterial meningoencephalitis and meningomyelitis, not elsewhere classified | G04.2 | ICD10CM | neurological |
| **844** | encephalopathy | Tropical spastic paraplegia | G04.1 | ICD10CM | neurological |
| **845** | encephalopathy | Postencephalitic parkinsonism | G21.3 | ICD10CM | neurological |
| **846** | encephalopathy | Plasmodium falciparum malaria with cerebral complications | B50.0 | ICD10CM | neurological |
| **847** | headache | Post-traumatic headache | G44.3 | ICD10CM | neurological |
| **848** | headache | Migraine without aura, intractable, with status migrainosus | G43.011 | ICD10CM | neurological |
| **849** | headache | Migraine without aura, not intractable, with status migrainosus | G43.001 | ICD10CM | neurological |
| **850** | headache | Status migrainosus | G43.2 | ICD10CM | neurological |
| **851** | headache | Cluster headache syndrome, unspecified | G44.00 | ICD10CM | neurological |
| **852** | headache | Migraine, unspecified, intractable | G43.91 | ICD10CM | neurological |
| **853** | headache | Hemicrania continua | G44.51 | ICD10CM | neurological |
| **854** | headache | Persistent migraine aura with cerebral infarction, not intractable | G43.60 | ICD10CM | neurological |
| **855** | headache | Ophthalmoplegic migraine | G43.B | ICD10CM | neurological |
| **856** | headache | Persistent migraine aura with cerebral infarction, not intractable, without status migrainosus | G43.609 | ICD10CM | neurological |
| **857** | headache | Migraine with aura, not intractable, with status migrainosus | G43.101 | ICD10CM | neurological |
| **858** | headache | Chronic paroxysmal hemicrania | G44.04 | ICD10CM | neurological |
| **859** | headache | Drug-induced headache, not elsewhere classified, not intractable | G44.40 | ICD10CM | neurological |
| **860** | headache | Menstrual migraine, intractable | G43.83 | ICD10CM | neurological |
| **861** | headache | Ophthalmoplegic migraine, not intractable | G43.B0 | ICD10CM | neurological |
| **862** | headache | Other headache syndromes | G44 | ICD10CM | neurological |
| **863** | headache | Persistent migraine aura without cerebral infarction, not intractable, with status migrainosus | G43.501 | ICD10CM | neurological |
| **864** | headache | Menstrual migraine, not intractable, with status migrainosus | G43.D01 | ICD10CM | neurological |
| **865** | headache | Other specified headache syndromes | G44.8 | ICD10CM | neurological |
| **866** | headache | Post-traumatic headache, unspecified, intractable | G44.301 | ICD10CM | neurological |
| **867** | headache | Episodic paroxysmal hemicrania, not intractable | G44.039 | ICD10CM | neurological |
| **868** | headache | Migraine without aura, not intractable | G43.00 | ICD10CM | neurological |
| **869** | headache | Other trigeminal autonomic cephalgias (TAC), intractable | G44.091 | ICD10CM | neurological |

| | | | | | |
|---|---|---|---|---|---|
| **870** | headache | Menstrual migraine, intractable, without status migrainosus | G43.839 | ICD10CM | neurological |
| **871** | headache | Persistent migraine aura without cerebral infarction, intractable | G43.51 | ICD10CM | neurological |
| **872** | headache | Migraine with aura, not intractable | G43.10 | ICD10CM | neurological |
| **873** | headache | Short lasting unilateral neuralgiform headache with conjunctival injection and tearing (SUNCT), not intractable | G44.059 | ICD10CM | neurological |
| **874** | headache | Chronic migraine without aura | G43.7 | ICD10CM | neurological |
| **875** | headache | Episodic paroxysmal hemicrania | G44.03 | ICD10CM | neurological |
| **876** | headache | Hemiplegic migraine | G43.4 | ICD10CM | neurological |
| **877** | headache | Chronic migraine without aura, intractable | G43.71 | ICD10CM | neurological |
| **878** | headache | Chronic post-traumatic headache, not intractable | G44.329 | ICD10CM | neurological |
| **879** | headache | Periodic headache syndromes in child or adult, not intractable, with status migrainosus | G43.C01 | ICD10CM | neurological |
| **880** | headache | Cluster headaches and other trigeminal autonomic cephalgias (TAC) | G44.0 | ICD10CM | neurological |
| **881** | headache | Migraine, unspecified | G43.9 | ICD10CM | neurological |
| **882** | headache | Persistent migraine aura with cerebral infarction, intractable, with status migrainosus | G43.611 | ICD10CM | neurological |
| **883** | headache | Migraine with aura | G43.1 | ICD10CM | neurological |
| **884** | headache | Persistent migraine aura without cerebral infarction | G43.5 | ICD10CM | neurological |
| **885** | headache | Migraine without aura | G43.0 | ICD10CM | neurological |
| **886** | headache | Headache associated with sexual activity | G44.82 | ICD10CM | neurological |
| **887** | headache | Post-traumatic headache, unspecified, not intractable | G44.309 | ICD10CM | neurological |
| **888** | headache | Menstrual migraine, intractable, without status migrainosus | G43.D19 | ICD10CM | neurological |
| **889** | headache | Abdominal migraine, intractable | G43.D1 | ICD10CM | neurological |
| **890** | headache | Episodic paroxysmal hemicrania, intractable | G44.031 | ICD10CM | neurological |
| **891** | headache | Chronic cluster headache, intractable | G44.021 | ICD10CM | neurological |
| **892** | headache | Cyclical vomiting, intractable, without status migrainosus | G43.A19 | ICD10CM | neurological |
| **893** | headache | Short lasting unilateral neuralgiform headache with conjunctival injection and tearing (SUNCT) | G44.05 | ICD10CM | neurological |
| **894** | headache | Cyclical vomiting | G43.A | ICD10CM | neurological |
| **895** | headache | Ophthalmoplegic migraine, intractable, with status migrainosus | G43.B11 | ICD10CM | neurological |
| **896** | headache | Persistent migraine aura without cerebral infarction, not intractable, without status migrainosus | G43.509 | ICD10CM | neurological |
| **897** | headache | Chronic tension-type headache, intractable | G44.221 | ICD10CM | neurological |
| **898** | headache | Chronic post-traumatic headache | G44.32 | ICD10CM | neurological |
| **899** | headache | Tension-type headache, unspecified, intractable | G44.201 | ICD10CM | neurological |
| **900** | headache | Hemiplegic migraine, intractable | G43.41 | ICD10CM | neurological |
| **901** | headache | Vascular headache, not elsewhere classified, intractable | G44.11 | ICD10CM | neurological |

| | | | | | |
|---|---|---|---|---|---|
| **902** | headache | Cyclical vomiting, not intractable, with status migrainosus | G43.A01 | ICD10CM | neurological |
| **903** | headache | Chronic paroxysmal hemicrania, not intractable | G44.049 | ICD10CM | neurological |
| **904** | headache | Short lasting unilateral neuralgiform headache with conjunctival injection and tearing (SUNCT), intractable | G44.051 | ICD10CM | neurological |
| **905** | headache | Cyclical vomiting, not intractable, without status migrainosus | G43.A09 | ICD10CM | neurological |
| **906** | headache | Headache | R51 | ICD10CM | neurological |
| **907** | headache | Persistent migraine aura with cerebral infarction, intractable, without status migrainosus | G43.619 | ICD10CM | neurological |
| **908** | headache | Persistent migraine aura with cerebral infarction | G43.6 | ICD10CM | neurological |
| **909** | headache | Persistent migraine aura without cerebral infarction, not intractable | G43.50 | ICD10CM | neurological |
| **910** | headache | Menstrual migraine, not intractable | G43.82 | ICD10CM | neurological |
| **911** | headache | Persistent migraine aura with cerebral infarction, not intractable, with status migrainosus | G43.601 | ICD10CM | neurological |
| **912** | headache | Other migraine | G43.8 | ICD10CM | neurological |
| **913** | headache | Cyclical vomiting, intractable, with status migrainosus | G43.A11 | ICD10CM | neurological |
| **914** | headache | Episodic cluster headache, not intractable | G44.019 | ICD10CM | neurological |
| **915** | headache | Migraine, unspecified, intractable, without status migrainosus | G43.919 | ICD10CM | neurological |
| **916** | headache | Drug-induced headache, not elsewhere classified, intractable | G44.41 | ICD10CM | neurological |
| **917** | headache | Hemiplegic migraine, not intractable, without status migrainosus | G43.409 | ICD10CM | neurological |
| **918** | headache | Chronic cluster headache, not intractable | G44.029 | ICD10CM | neurological |
| **919** | headache | Migraine without aura, intractable, without status migrainosus | G43.019 | ICD10CM | neurological |
| **920** | headache | Hemiplegic migraine, not intractable | G43.40 | ICD10CM | neurological |
| **921** | headache | Migraine with aura, intractable, without status migrainosus | G43.119 | ICD10CM | neurological |
| **922** | headache | Migraine, unspecified, not intractable | G43.90 | ICD10CM | neurological |
| **923** | headache | Episodic tension-type headache | G44.21 | ICD10CM | neurological |
| **924** | headache | Other trigeminal autonomic cephalgias (TAC), not intractable | G44.099 | ICD10CM | neurological |
| **925** | headache | Ophthalmoplegic migraine, not intractable, with status migrainosus | G43.B01 | ICD10CM | neurological |
| **926** | headache | Other migraine, not intractable, with status migrainosus | G43.801 | ICD10CM | neurological |
| **927** | headache | Complicated headache syndromes | G44.5 | ICD10CM | neurological |
| **928** | headache | Other migraine, not intractable | G43.80 | ICD10CM | neurological |
| **929** | headache | Ophthalmoplegic migraine, intractable, without status migrainosus | G43.B19 | ICD10CM | neurological |
| **930** | headache | Other complicated headache syndrome | G44.59 | ICD10CM | neurological |
| **931** | headache | Hemiplegic migraine, intractable, without status migrainosus | G43.419 | ICD10CM | neurological |

| | | | | | |
|---|---|---|---|---|---|
| **932** | headache | Tension-type headache | G44.2 | ICD10CM | neurological |
| **933** | headache | New daily persistent headache (NDPH) | G44.52 | ICD10CM | neurological |
| **934** | headache | Migraine, unspecified, not intractable, with status migrainosus | G43.901 | ICD10CM | neurological |
| **935** | headache | Post-traumatic headache, unspecified | G44.30 | ICD10CM | neurological |
| **936** | headache | Episodic cluster headache | G44.01 | ICD10CM | neurological |
| **937** | headache | Other migraine, intractable, with status migrainosus | G43.811 | ICD10CM | neurological |
| **938** | headache | Chronic paroxysmal hemicrania, intractable | G44.041 | ICD10CM | neurological |
| **939** | headache | Menstrual migraine, not intractable, without status migrainosus | G43.829 | ICD10CM | neurological |
| **940** | headache | Periodic headache syndromes in child or adult, intractable | G43.C1 | ICD10CM | neurological |
| **941** | headache | Migraine with aura, not intractable, without status migrainosus | G43.109 | ICD10CM | neurological |
| **942** | headache | Other headache syndrome | G44.89 | ICD10CM | neurological |
| **943** | headache | Migraine with aura, intractable | G43.11 | ICD10CM | neurological |
| **944** | headache | Periodic headache syndromes in child or adult, intractable, without status migrainosus | G43.C19 | ICD10CM | neurological |
| **945** | headache | Chronic migraine without aura, intractable, without status migrainosus | G43.719 | ICD10CM | neurological |
| **946** | headache | Episodic tension-type headache, not intractable | G44.219 | ICD10CM | neurological |
| **947** | headache | Menstrual migraine, not intractable, with status migrainosus | G43.821 | ICD10CM | neurological |
| **948** | headache | Ophthalmoplegic migraine, intractable | G43.B1 | ICD10CM | neurological |
| **949** | headache | Acute post-traumatic headache, intractable | G44.311 | ICD10CM | neurological |
| **950** | headache | Other migraine, intractable | G43.81 | ICD10CM | neurological |
| **951** | headache | Chronic tension-type headache | G44.22 | ICD10CM | neurological |
| **952** | headache | Primary cough headache | G44.83 | ICD10CM | neurological |
| **953** | headache | Persistent migraine aura without cerebral infarction, intractable, with status migrainosus | G43.511 | ICD10CM | neurological |
| **954** | headache | Cluster headache syndrome, unspecified, intractable | G44.001 | ICD10CM | neurological |
| **955** | headache | Ophthalmoplegic migraine, not intractable, without status migrainosus | G43.B09 | ICD10CM | neurological |
| **956** | headache | Vascular headache, not elsewhere classified | G44.1 | ICD10CM | neurological |
| **957** | headache | Persistent migraine aura with cerebral infarction, intractable | G43.61 | ICD10CM | neurological |
| **958** | headache | Other migraine, intractable | G43.891 | ICD10CM | neurological |
| **959** | headache | Hypnic headache | G44.81 | ICD10CM | neurological |
| **960** | headache | Migraine with aura, intractable, with status migrainosus | G43.111 | ICD10CM | neurological |
| **961** | headache | Primary thunderclap headache | G44.53 | ICD10CM | neurological |
| **962** | headache | Migraine, unspecified, not intractable, without status migrainosus | G43.909 | ICD10CM | neurological |
| **963** | headache | Vascular headache, not elsewhere classified, not intractable | G44.10 | ICD10CM | neurological |

| | | | | | |
|---|---|---|---|---|---|
| **964** | headache | Periodic headache syndromes in child or adult, not intractable, without status migrainosus | G43.C09 | ICD10CM | neurological |
| **965** | headache | Abdominal migraine, not intractable | G43.D0 | ICD10CM | neurological |
| **966** | headache | Migraine, unspecified, intractable, with status migrainosus | G43.911 | ICD10CM | neurological |
| **967** | headache | Hemiplegic migraine, not intractable, with status migrainosus | G43.401 | ICD10CM | neurological |
| **968** | headache | Chronic migraine without aura, intractable, with status migrainosus | G43.711 | ICD10CM | neurological |
| **969** | headache | Persistent migraine aura without cerebral infarction, intractable, without status migrainosus | G43.519 | ICD10CM | neurological |
| **970** | headache | Other migraine, intractable, without status migrainosus | G43.819 | ICD10CM | neurological |
| **971** | headache | Chronic migraine without aura, not intractable, with status migrainosus | G43.701 | ICD10CM | neurological |
| **972** | headache | Drug-induced headache, not elsewhere classified | G44.4 | ICD10CM | neurological |
| **973** | headache | Other migraine, not intractable, without status migrainosus | G43.809 | ICD10CM | neurological |
| **974** | headache | Migraine without aura, intractable | G43.01 | ICD10CM | neurological |
| **975** | headache | Periodic headache syndromes in child or adult, intractable, with status migrainosus | G43.C11 | ICD10CM | neurological |
| **976** | headache | Migraine without aura, not intractable, without status migrainosus | G43.009 | ICD10CM | neurological |
| **977** | headache | Cyclical vomiting, in migraine, intractable | G43.A1 | ICD10CM | neurological |
| **978** | headache | Migraine | G43 | ICD10CM | neurological |
| **979** | headache | Acute post-traumatic headache, not intractable | G44.319 | ICD10CM | neurological |
| **980** | headache | Chronic migraine without aura, not intractable, without status migrainosus | G43.709 | ICD10CM | neurological |
| **981** | headache | Other migraine, not intractable | G43.899 | ICD10CM | neurological |
| **982** | headache | Headache, unspecified | R51.9 | ICD10CM | neurological |
| **983** | headache | Tension-type headache, unspecified | G44.20 | ICD10CM | neurological |
| **984** | headache | Chronic cluster headache | G44.02 | ICD10CM | neurological |
| **985** | headache | Menstrual migraine, intractable, with status migrainosus | G43.D11 | ICD10CM | neurological |
| **986** | headache | Abdominal migraine | G43.D | ICD10CM | neurological |
| **987** | headache | Hemiplegic migraine, intractable, with status migrainosus | G43.411 | ICD10CM | neurological |
| **988** | headache | Primary stabbing headache | G44.85 | ICD10CM | neurological |
| **989** | headache | Menstrual migraine, not intractable, without status migrainosus | G43.D09 | ICD10CM | neurological |
| **990** | headache | Acute post-traumatic headache | G44.31 | ICD10CM | neurological |
| **991** | headache | Other trigeminal autonomic cephalgias (TAC) | G44.09 | ICD10CM | neurological |
| **992** | headache | Episodic cluster headache, intractable | G44.011 | ICD10CM | neurological |
| **993** | headache | Primary exertional headache | G44.84 | ICD10CM | neurological |
| **994** | headache | Headache with orthostatic component, not elsewhere classified | R51.0 | ICD10CM | neurological |

| | | | | | |
|---|---|---|---|---|---|
| **995** | headache | Tension-type headache, unspecified, not intractable | G44.209 | ICD10CM | neurological |
| **996** | headache | Chronic tension-type headache, not intractable | G44.229 | ICD10CM | neurological |
| **997** | headache | Episodic tension-type headache, intractable | G44.211 | ICD10CM | neurological |
| **998** | headache | Cluster headache syndrome, unspecified, not intractable | G44.009 | ICD10CM | neurological |
| **999** | headache | Chronic migraine without aura, not intractable | G43.70 | ICD10CM | neurological |
| **1000** | headache | Chronic post-traumatic headache, intractable | G44.321 | ICD10CM | neurological |
| **1001** | headache | Menstrual migraine, intractable, with status migrainosus | G43.831 | ICD10CM | neurological |
| **1002** | headache | Periodic headache syndromes in child or adult | G43.C | ICD10CM | neurological |
| **1003** | headache | Cyclical vomiting, in migraine, not intractable | G43.A0 | ICD10CM | neurological |
| **1004** | headache | Periodic headache syndromes in child or adult, not intractable | G43.C0 | ICD10CM | neurological |
| **1005** | nervous system signs and symptoms | Tetany | R29.0 | ICD10CM | neurological |
| **1006** | nervous system signs and symptoms | Transient paralysis | R29.5 | ICD10CM | neurological |
| **1007** | nervous system signs and symptoms | Unspecified symptoms and signs involving the nervous system | R29.90 | ICD10CM | neurological |
| **1008** | nervous system signs and symptoms | Tremor, unspecified | R25.1 | ICD10CM | neurological |
| **1009** | nervous system signs and symptoms | Meningismus | R29.1 | ICD10CM | neurological |
| **1010** | nervous system signs and symptoms | Abnormal reflex | R29.2 | ICD10CM | neurological |
| **1011** | nervous system signs and symptoms | Other abnormal involuntary movements | R25.8 | ICD10CM | neurological |
| **1012** | nervous system signs and symptoms | Unspecified abnormal involuntary movements | R25.9 | ICD10CM | neurological |
| **1013** | nervous system signs and symptoms | Fasciculation | R25.3 | ICD10CM | neurological |
| **1014** | nervous system signs and symptoms | Facial weakness | R29.810 | ICD10CM | neurological |
| **1015** | nervous system signs and symptoms | Abnormal head movements | R25.0 | ICD10CM | neurological |
| **1016** | nervous system signs and symptoms | Other symptoms and signs involving the nervous system | R29.818 | ICD10CM | neurological |
| **1017** | acute kidney injury | Other acute kidney failure | N17.8 | ICD10CM | renal |
| **1018** | acute kidney injury | Acute kidney failure with acute cortical necrosis | N17.1 | ICD10CM | renal |
| **1019** | acute kidney injury | Unspecified kidney failure | N19 | ICD10CM | renal |
| **1020** | acute kidney injury | Acute kidney failure with medullary necrosis | N17.2 | ICD10CM | renal |
| **1021** | acute kidney injury | Acute kidney failure with tubular necrosis | N17.0 | ICD10CM | renal |
| **1022** | acute kidney injury | Acute kidney failure, unspecified | N17.9 | ICD10CM | renal |
| **1023** | acute kidney injury | Acute kidney failure | N17 | ICD10CM | renal |
| **1024** | Creatinine | CREATININE (SERUM) \| CREATININE \| 2160-0 \| 15 | 2160-0 | LOINC | renal |

| | | | | | |
|---|---|---|---|---|---|
| **1025** | Creatinine | COMPREHENSIVE METABOLIC PANEL\|CREATININE\|80053 | 2160-0 | LOINC | renal |
| **1026** | Creatinine | CREATININE, EAST\|2160-0 | 2160-0 | LOINC | renal |
| **1027** | Creatinine | COMPREHENSIVE METABOLIC PANEL (CMP) -Q\|CREATININE\|80053 | 2160-0 | LOINC | renal |
| **1028** | Creatinine | Creatinine \| 2160-0 | 2160-0 | LOINC | renal |
| **1029** | Creatinine | CMP-NEWBORN <29 DAYS\|CREATININE\|2160-0 | 2160-0 | LOINC | renal |
| **1030** | Creatinine | CREATININE (WHOLE BLOOD) \| BASIC METABOLIC PANEL ( | 38483-4 | LOINC | renal |
| **1031** | Creatinine | TPN PANEL\|CREATININE\|84295 | 2160-0 | LOINC | renal |
| **1032** | Creatinine | CREATININE SERUM/PLASMA - UHC MN \| RENAL FUNCTION | 2160-0 | LOINC | renal |
| **1033** | Creatinine | CREATININE, POC\|2160-0 | 2160-0 | LOINC | renal |
| **1034** | Creatinine | CREATININE | 2160-0 | LOINC | renal |
| **1035** | Creatinine | CREATININE, MAU\|2160-0 | 2160-0 | LOINC | renal |
| **1036** | Creatinine | COMPREHENSIVE METABOLIC PANEL\|CREATININE, SERUM-LC\|2160-0 | 2160-0 | LOINC | renal |
| **1037** | Creatinine | CREATININE PROFILE\|CREATININE\|2160-0 | 2160-0 | LOINC | renal |
| **1038** | Creatinine | COMP MET PANEL,NO REFLEX\|CREATININE\|80053 | 2160-0 | LOINC | renal |
| **1039** | Creatinine | RENAL FUNCTION PANEL\|CREATININE\|2160-0 | 2160-0 | LOINC | renal |
| **1040** | Creatinine | CREATININE\|2160-0 | 2160-0 | LOINC | renal |
| **1041** | Creatinine | CREATININE LEVEL\|2160-0 | 2160-0 | LOINC | renal |
| **1042** | Creatinine | CREATININE, PLASMA OR SERUM\|2160-0 | 2160-0 | LOINC | renal |
| **1043** | Creatinine | Creatinine, Ser/Plas \| 2160-0 | 2160-0 | LOINC | renal |
| **1044** | Creatinine | Creatinine | 2160-0 | LOINC | renal |
| **1045** | Creatinine | CREATININE LEVEL EXT\|2160-0 | 2160-0 | LOINC | renal |
| **1046** | Creatinine | CREATININE SERUM/PLASMA - UHC MN \| BASIC METABOLIC | 2160-0 | LOINC | renal |
| **1047** | Creatinine | CREATININE SERUM/PLASMA - UHC MN \| NEONATAL COMPRE | 2160-0 | LOINC | renal |
| **1048** | Creatinine | CREATININE SERUM/PLASMA - UHC MN \| COMPREHENSIVE M | 2160-0 | LOINC | renal |
| **1049** | Creatinine | CREATININE (I-STAT)\|38483-4 | 38483-4 | LOINC | renal |
| **1050** | Creatinine | BASIC METABOLIC PANEL\|CREATININE\|2160-0 | 2160-0 | LOINC | renal |
| **1051** | Creatinine | CREATININE SERUM/PLASMA - UHC MN \| CREATININE BLOO | 2160-0 | LOINC | renal |
| **1052** | Creatinine | BASIC METABOLIC PANEL\|CREATININE, SERUM-LC\|2160-0 | 2160-0 | LOINC | renal |
| **1053** | Creatinine | COMPREHENSIVE METABOLIC PANEL (CMP) -Q\|CREATININE-Q\|80053 | 2160-0 | LOINC | renal |
| **1054** | Creatinine | TPN PANEL\|CREATININE\|2160-0 | 2160-0 | LOINC | renal |
| **1055** | Creatinine | CREATININE (POCT) \| POC BASIC METABOLIC PANEL - PE | 38483-4 | LOINC | renal |

| | | | | | |
|---|---|---|---|---|---|
| **1056** | Creatinine | CREATININE, SERUM\|2160-0 | 2160-0 | LOINC | renal |
| **1057** | Creatinine | COMPREHENSIVE METABOLIC PANEL (CMP) -Q\|CREATININE-Q\|2160-0 | 2160-0 | LOINC | renal |
| **1058** | Creatinine | TPN PANEL NEWBORN\|CREATININE\|2160-0 | 2160-0 | LOINC | renal |
| **1059** | Creatinine | CREATININE, PLASMA OR SERUM (EXT RSLT)\|2160-0 | 2160-0 | LOINC | renal |
| **1060** | Creatinine | CREATININE (SERUM) \| COMPREHENSIVE METABOLIC PANEL | 2160-0 | LOINC | renal |
| **1061** | Creatinine | COMP METABOLIC PANEL (12) - LC\|CREATININE, SERUM-LC\|2160-0 | 2160-0 | LOINC | renal |
| **1062** | Creatinine | CREATININE (SERUM) \| CREATININE \| 2160-0 \| 1230100 | 2160-0 | LOINC | renal |
| **1063** | Creatinine | CREATININE, CANAL WINCHESTER\|2160-0 | 2160-0 | LOINC | renal |
| **1064** | Creatinine | CREATININE (SERUM) \| RENAL FUNCTION PANEL \| 2160-0 | 2160-0 | LOINC | renal |
| **1065** | Creatinine | CREATININE, NORTHEAST\|2160-0 | 2160-0 | LOINC | renal |
| **1066** | Creatinine | BASIC METABOLIC PANEL\|CREATININE\|80048 | 2160-0 | LOINC | renal |
| **1067** | Creatinine | CREATININE (SERUM) \| BASIC METABOLIC PANEL \| 2160- | 2160-0 | LOINC | renal |
| **1068** | Creatinine | COMPREHENSIVE METABOLIC PANEL\|CREATININE-Q\|2160-0 | 2160-0 | LOINC | renal |
| **1069** | Creatinine | COMPREHENSIVE METABOLIC PANEL\|CREATININE\|2160-0 | 2160-0 | LOINC | renal |
| **1070** | dialysis | End-stage renal disease (ESRD) related services for dialysis less than a full month of service, per day; for patients 2-11 years of age | 90968 | CPT4 | renal |
| **1071** | dialysis | Hemodialysis | 39.95 | ICD9Proc | renal |
| **1072** | dialysis | Insertion of cannula for hemodialysis, other purpose (separate procedure); vein to vein | 36800 | CPT4 | renal |
| **1073** | dialysis | Dialysis training, patient, including helper where applicable, any mode, course not completed, per training session | 90993 | CPT4 | renal |
| **1074** | dialysis | Insertion of tunneled intraperitoneal catheter for dialysis, open | 49421 | CPT4 | renal |
| **1075** | dialysis | Dialysis training, patient, including helper where applicable, any mode, completed course | 90989 | CPT4 | renal |
| **1076** | dialysis | End-stage renal disease (ESRD) related services for home dialysis per full month, for patients 12-19 years of age to include monitoring for the adequacy of nutrition, assessment of growth and development, and counseling of parents | 90965 | CPT4 | renal |
| **1077** | dialysis | Hemodialysis procedure requiring repeated evaluation(s) with or without substantial revision of dialysis prescription | 90937 | CPT4 | renal |
| **1078** | dialysis | Unlisted dialysis procedure, inpatient or outpatient | 90999 | CPT4 | renal |
| **1079** | dialysis | Catheter, hemodialysis/peritoneal, short-term | C1752 | HCPCS | renal |
| **1080** | dialysis | Injection, darbepoetin alfa, 1 microgram (for esrd on dialysis) | J0882 | HCPCS | renal |
| **1081** | dialysis | Dialysis procedure other than hemodialysis (eg, peritoneal dialysis, hemofiltration, or other | 90945 | CPT4 | renal |

| | | | | | |
|---|---|---|---|---|---|
| | | continuous renal replacement therapies), with single evaluation by a physician or other qualified health care professional | | | |
| **1082** | dialysis | Injection, epoetin alfa, 100 units (for esrd on dialysis) | Q4081 | HCPCS | renal |
| **1083** | dialysis | End-stage renal disease (ESRD) related services for dialysis less than a full month of service, per day; for patients 12-19 years of age | 90969 | CPT4 | renal |
| **1084** | dialysis | Hemodialysis procedure with single evaluation by a physician or other qualified health care professional | 90935 | CPT4 | renal |
| **1085** | dialysis | Catheter, hemodialysis/peritoneal, long-term | C1750 | HCPCS | renal |
| **1086** | dialysis | End-stage renal disease (ESRD) related services for home dialysis per full month, for patients 20 years of age and older | 90966 | CPT4 | renal |
| **1087** | dialysis | Dialysis procedure other than hemodialysis (eg, peritoneal dialysis, hemofiltration, or other continuous renal replacement therapies) requiring repeated evaluations by a physician or other qualified health care professional, with or without substantial re | 90947 | CPT4 | renal |
| **1088** | dialysis | Peritoneal dialysis | 54.98 | ICD9Proc | renal |
| **1089** | dialysis | Insertion of tunneled intraperitoneal catheter (eg, dialysis, intraperitoneal chemotherapy instillation, management of ascites), complete procedure, including imaging guidance, catheter placement, contrast injection when performed, and radiological superv | 49418 | CPT4 | renal |
| **1090** | dialysis | End-stage renal disease (ESRD) related services for home dialysis per full month, for patients 2-11 years of age to include monitoring for the adequacy of nutrition, assessment of growth and development, and counseling of parents | 90964 | CPT4 | renal |
| **1091** | dialysis | End-stage renal disease (ESRD) related services for home dialysis per full month, for patients younger than 2 years of age to include monitoring for the adequacy of nutrition, assessment of growth and development, and counseling of parents | 90963 | CPT4 | renal |
| **1092** | fluid and electrolyte disturbance | Hypo-osmolality and hyponatremia | E87.1 | ICD10CM | renal |
| **1093** | fluid and electrolyte disturbance | Hyperkalemia | E87.5 | ICD10CM | renal |
| **1094** | fluid and electrolyte disturbance | Transfusion associated circulatory overload | E87.71 | ICD10CM | renal |
| **1095** | fluid and electrolyte disturbance | Hyperosmolality and hypernatremia | E87.0 | ICD10CM | renal |
| **1096** | fluid and electrolyte disturbance | Acidosis | E87.2 | ICD10CM | renal |
| **1097** | fluid and electrolyte disturbance | Fluid overload, unspecified | E87.70 | ICD10CM | renal |
| **1098** | fluid and electrolyte disturbance | Volume depletion, unspecified | E86.9 | ICD10CM | renal |
| **1099** | fluid and electrolyte disturbance | Alkalosis | E87.3 | ICD10CM | renal |
| **1100** | fluid and electrolyte disturbance | Other fluid overload | E87.79 | ICD10CM | renal |

| | | | | | |
|---|---|---|---|---|---|
| **1101** | fluid and electrolyte disturbance | Fluid overload | E87.7 | ICD10CM | renal |
| **1102** | fluid and electrolyte disturbance | Other disorders of fluid, electrolyte and acid-base balance | E87 | ICD10CM | renal |
| **1103** | fluid and electrolyte disturbance | Hypovolemia | E86.1 | ICD10CM | renal |
| **1104** | fluid and electrolyte disturbance | Dehydration | E86.0 | ICD10CM | renal |
| **1105** | fluid and electrolyte disturbance | Mixed disorder of acid-base balance | E87.4 | ICD10CM | renal |
| **1106** | fluid and electrolyte disturbance | Hypokalemia | E87.6 | ICD10CM | renal |
| **1107** | fluid and electrolyte disturbance | Other disorders of electrolyte and fluid balance, not elsewhere classified | E87.8 | ICD10CM | renal |
| **1108** | acute respiratory distress syndrome | Acute respiratory distress syndrome | J80 | ICD10CM | respiratory |
| **1109** | bronchiolitis | Acute bronchiolitis, unspecified | J21.9 | ICD10CM | respiratory |
| **1110** | bronchiolitis | Acute bronchiolitis due to other specified organisms | J21.8 | ICD10CM | respiratory |
| **1111** | bronchiolitis | Respiratory bronchiolitis interstitial lung disease | J84.115 | ICD10CM | respiratory |
| **1112** | bronchiolitis | Acute bronchiolitis due to human metapneumovirus | J21.1 | ICD10CM | respiratory |
| **1113** | bronchiolitis | Acute bronchiolitis due to respiratory syncytial virus | J21.0 | ICD10CM | respiratory |
| **1114** | bronchiolitis | Acute bronchiolitis | J21 | ICD10CM | respiratory |
| **1115** | bronchitis | Acute bronchitis, unspecified | J20.9 | ICD10CM | respiratory |
| **1116** | bronchitis | Acute bronchitis due to rhinovirus | J20.6 | ICD10CM | respiratory |
| **1117** | bronchitis | Acute bronchitis due to streptococcus | J20.2 | ICD10CM | respiratory |
| **1118** | bronchitis | Acute bronchitis due to Mycoplasma pneumoniae | J20.0 | ICD10CM | respiratory |
| **1119** | bronchitis | Acute bronchitis due to respiratory syncytial virus | J20.5 | ICD10CM | respiratory |
| **1120** | bronchitis | Acute bronchitis due to coxsackievirus | J20.3 | ICD10CM | respiratory |
| **1121** | bronchitis | Acute bronchitis | J20 | ICD10CM | respiratory |
| **1122** | bronchitis | Bronchitis, not specified as acute or chronic | J40 | ICD10CM | respiratory |
| **1123** | bronchitis | Acute bronchitis due to Hemophilus influenzae | J20.1 | ICD10CM | respiratory |
| **1124** | bronchitis | Acute bronchitis due to echovirus | J20.7 | ICD10CM | respiratory |
| **1125** | bronchitis | Acute bronchitis due to other specified organisms | J20.8 | ICD10CM | respiratory |
| **1126** | bronchitis | Acute bronchitis due to parainfluenza virus | J20.4 | ICD10CM | respiratory |
| **1127** | cough | Cough | R05 | ICD10CM | respiratory |
| **1128** | pleurisy, pleural effusion and pulmonary collapse | Pleural effusion, not elsewhere classified | J90 | ICD10CM | respiratory |
| **1129** | pleurisy, pleural effusion and pulmonary collapse | Pleural plaque with presence of asbestos | J92.0 | ICD10CM | respiratory |
| **1130** | pleurisy, pleural effusion and pulmonary collapse | Other specified pleural conditions | J94.8 | ICD10CM | respiratory |
| **1131** | pleurisy, pleural effusion and pulmonary collapse | Pleural effusion in other conditions classified elsewhere | J91.8 | ICD10CM | respiratory |

| | | | | | |
|---|---|---|---|---|---|
| **1132** | pleurisy, pleural effusion and pulmonary collapse | Compensatory emphysema | J98.3 | ICD10CM | respiratory |
| **1133** | pleurisy, pleural effusion and pulmonary collapse | Atelectasis | J98.11 | ICD10CM | respiratory |
| **1134** | pleurisy, pleural effusion and pulmonary collapse | Interstitial emphysema | J98.2 | ICD10CM | respiratory |
| **1135** | pleurisy, pleural effusion and pulmonary collapse | Fibrothorax | J94.1 | ICD10CM | respiratory |
| **1136** | pleurisy, pleural effusion and pulmonary collapse | Other pulmonary collapse | J98.19 | ICD10CM | respiratory |
| **1137** | pleurisy, pleural effusion and pulmonary collapse | Malignant pleural effusion | J91.0 | ICD10CM | respiratory |
| **1138** | pleurisy, pleural effusion and pulmonary collapse | Hemothorax | J94.2 | ICD10CM | respiratory |
| **1139** | pleurisy, pleural effusion and pulmonary collapse | Pyothorax with fistula | J86.0 | ICD10CM | respiratory |
| **1140** | pleurisy, pleural effusion and pulmonary collapse | Chylous effusion | J94.0 | ICD10CM | respiratory |
| **1141** | pleurisy, pleural effusion and pulmonary collapse | Pleural plaque without asbestos | J92.9 | ICD10CM | respiratory |
| **1142** | pleurisy, pleural effusion and pulmonary collapse | Tuberculous pleurisy | A15.6 | ICD10CM | respiratory |
| **1143** | pleurisy, pleural effusion and pulmonary collapse | Pyothorax without fistula | J86.9 | ICD10CM | respiratory |
| **1144** | pleurisy, pleural effusion and pulmonary collapse | Pleural condition, unspecified | J94.9 | ICD10CM | respiratory |
| **1145** | pneumonia | Congenital pneumonia due to streptococcus, group B | P23.3 | ICD10CM | respiratory |
| **1146** | pneumonia | Chlamydial pneumonia | J16.0 | ICD10CM | respiratory |
| **1147** | pneumonia | Lobar pneumonia, unspecified organism | J18.1 | ICD10CM | respiratory |
| **1148** | pneumonia | Pneumonia due to staphylococcus, unspecified | J15.20 | ICD10CM | respiratory |
| **1149** | pneumonia | Pneumonia due to Methicillin resistant Staphylococcus aureus | J15.212 | ICD10CM | respiratory |
| **1150** | pneumonia | Influenza due to unidentified influenza virus with unspecified type of pneumonia | J11.00 | ICD10CM | respiratory |
| **1151** | pneumonia | Pneumonia due to other streptococci | J15.4 | ICD10CM | respiratory |
| **1152** | pneumonia | Congenital pneumonia due to staphylococcus | P23.2 | ICD10CM | respiratory |
| **1153** | pneumonia | Rubella pneumonia | B06.81 | ICD10CM | respiratory |
| **1154** | pneumonia | Pulmonary toxoplasmosis | B58.3 | ICD10CM | respiratory |
| **1155** | pneumonia | Whooping cough, unspecified species with pneumonia | A37.91 | ICD10CM | respiratory |
| **1156** | pneumonia | Influenza due to other identified influenza virus with other specified pneumonia | J10.08 | ICD10CM | respiratory |
| **1157** | pneumonia | Congenital pneumonia due to Pseudomonas | P23.5 | ICD10CM | respiratory |
| **1158** | pneumonia | Cytomegaloviral pneumonitis | B25.0 | ICD10CM | respiratory |
| **1159** | pneumonia | Pneumonia due to Pseudomonas | J15.1 | ICD10CM | respiratory |
| **1160** | pneumonia | Congenital pneumonia due to other bacterial agents | P23.6 | ICD10CM | respiratory |
| **1161** | pneumonia | Bronchopneumonia, unspecified organism | J18.0 | ICD10CM | respiratory |

| | | | | | |
|---|---|---|---|---|---|
| **1162** | pneumonia | Pneumonia due to other infectious organisms, not elsewhere classified | J16 | ICD10CM | respiratory |
| **1163** | pneumonia | Influenza due to other identified influenza virus with the same other identified influenza virus pneumonia | J10.01 | ICD10CM | respiratory |
| **1164** | pneumonia | Respiratory syncytial virus pneumonia | J12.1 | ICD10CM | respiratory |
| **1165** | pneumonia | Pneumonia due to other specified infectious organisms | J16.8 | ICD10CM | respiratory |
| **1166** | pneumonia | Congenital pneumonia due to Escherichia coli | P23.4 | ICD10CM | respiratory |
| **1167** | pneumonia | Pneumonia due to Hemophilus influenzae | J14 | ICD10CM | respiratory |
| **1168** | pneumonia | Influenza due to identified novel influenza A virus with pneumonia | J09.X1 | ICD10CM | respiratory |
| **1169** | pneumonia | Other pneumonia, unspecified organism | J18.8 | ICD10CM | respiratory |
| **1170** | pneumonia | Whooping cough due to Bordetella pertussis with pneumonia | A37.01 | ICD10CM | respiratory |
| **1171** | pneumonia | Other viral pneumonia | J12.8 | ICD10CM | respiratory |
| **1172** | pneumonia | Ascariasis pneumonia | B77.81 | ICD10CM | respiratory |
| **1173** | pneumonia | Pneumonia due to Klebsiella pneumoniae | J15.0 | ICD10CM | respiratory |
| **1174** | pneumonia | Viral pneumonia, unspecified | J12.9 | ICD10CM | respiratory |
| **1175** | pneumonia | Pulmonary anthrax | A22.1 | ICD10CM | respiratory |
| **1176** | pneumonia | Congenital pneumonia due to other organisms | P23.8 | ICD10CM | respiratory |
| **1177** | pneumonia | Other viral pneumonia | J12.89 | ICD10CM | respiratory |
| **1178** | pneumonia | Chronic pulmonary coccidioidomycosis | B38.1 | ICD10CM | respiratory |
| **1179** | pneumonia | Ventilator associated pneumonia | J95.851 | ICD10CM | respiratory |
| **1180** | pneumonia | Pneumocystosis | B59 | ICD10CM | respiratory |
| **1181** | pneumonia | Pneumonia due to Streptococcus pneumoniae | J13 | ICD10CM | respiratory |
| **1182** | pneumonia | Pneumonia, unspecified organism | J18.9 | ICD10CM | respiratory |
| **1183** | pneumonia | Congenital pneumonia, unspecified | P23.9 | ICD10CM | respiratory |
| **1184** | pneumonia | Pneumonia due to Methicillin susceptible Staphylococcus aureus | J15.211 | ICD10CM | respiratory |
| **1185** | pneumonia | Gonococcal pneumonia | A54.84 | ICD10CM | respiratory |
| **1186** | pneumonia | Pulmonary nocardiosis | A43.0 | ICD10CM | respiratory |
| **1187** | pneumonia | Pneumonia due to Mycoplasma pneumoniae | J15.7 | ICD10CM | respiratory |
| **1188** | pneumonia | Pneumonia due to staphylococcus | J15.2 | ICD10CM | respiratory |
| **1189** | pneumonia | Whooping cough due to Bordetella parapertussis with pneumonia | A37.11 | ICD10CM | respiratory |
| **1190** | pneumonia | Whooping cough due to other Bordetella species with pneumonia | A37.81 | ICD10CM | respiratory |
| **1191** | pneumonia | Varicella pneumonia | B01.2 | ICD10CM | respiratory |
| **1192** | pneumonia | Unspecified bacterial pneumonia | J15.9 | ICD10CM | respiratory |
| **1193** | pneumonia | Pneumonic plague | A20.2 | ICD10CM | respiratory |
| **1194** | pneumonia | Hypostatic pneumonia, unspecified organism | J18.2 | ICD10CM | respiratory |
| **1195** | pneumonia | Pneumonia due to other Gram-negative bacteria | J15.6 | ICD10CM | respiratory |
| **1196** | pneumonia | Pneumonia due to other specified bacteria | J15.8 | ICD10CM | respiratory |

| | | | | | |
|---|---|---|---|---|---|
| **1197** | pneumonia | Influenza due to unidentified influenza virus with specified pneumonia | J11.08 | ICD10CM | respiratory |
| **1198** | pneumonia | Congenital pneumonia due to Chlamydia | P23.1 | ICD10CM | respiratory |
| **1199** | pneumonia | Adenoviral pneumonia | J12.0 | ICD10CM | respiratory |
| **1200** | pneumonia | Pneumonia due to streptococcus, group B | J15.3 | ICD10CM | respiratory |
| **1201** | pneumonia | Acute pulmonary coccidioidomycosis | B38.0 | ICD10CM | respiratory |
| **1202** | pneumonia | Salmonella pneumonia | A02.22 | ICD10CM | respiratory |
| **1203** | pneumonia | Human metapneumovirus pneumonia | J12.3 | ICD10CM | respiratory |
| **1204** | pneumonia | Pneumonia in diseases classified elsewhere | J17 | ICD10CM | respiratory |
| **1205** | pneumonia | Measles complicated by pneumonia | B05.2 | ICD10CM | respiratory |
| **1206** | pneumonia | Pulmonary tularemia | A21.2 | ICD10CM | respiratory |
| **1207** | pneumonia | Pulmonary candidiasis | B37.1 | ICD10CM | respiratory |
| **1208** | pneumonia | Influenza due to other identified influenza virus with unspecified type of pneumonia | J10.00 | ICD10CM | respiratory |
| **1209** | pneumonia | Acute pulmonary histoplasmosis capsulati | B39.0 | ICD10CM | respiratory |
| **1210** | pneumonia | Viral pneumonia, not elsewhere classified | J12 | ICD10CM | respiratory |
| **1211** | pneumonia | Congenital pneumonia due to viral agent | P23.0 | ICD10CM | respiratory |
| **1212** | pneumonia | Chronic pulmonary histoplasmosis capsulati | B39.1 | ICD10CM | respiratory |
| **1213** | pneumonia | Pneumonia due to other staphylococcus | J15.29 | ICD10CM | respiratory |
| **1214** | pneumonia | Syphilis of lung and bronchus | A52.72 | ICD10CM | respiratory |
| **1215** | pneumonia | Pulmonary mycobacterial infection | A31.0 | ICD10CM | respiratory |
| **1216** | pneumonia | Pulmonary histoplasmosis capsulati, unspecified | B39.2 | ICD10CM | respiratory |
| **1217** | pneumonia | Legionnaires' disease | A48.1 | ICD10CM | respiratory |
| **1218** | pneumonia | Typhoid pneumonia | A01.03 | ICD10CM | respiratory |
| **1219** | pneumonia | Bacterial pneumonia, not elsewhere classified | J15 | ICD10CM | respiratory |
| **1220** | pneumonia | Pneumonia due to staphylococcus aureus | J15.21 | ICD10CM | respiratory |
| **1221** | pneumonia | Abscess of lung with pneumonia | J85.1 | ICD10CM | respiratory |
| **1222** | pneumonia | Pneumonia, unspecified organism | J18 | ICD10CM | respiratory |
| **1223** | pneumonia | Parainfluenza virus pneumonia | J12.2 | ICD10CM | respiratory |
| **1224** | pneumonia | Pulmonary coccidioidomycosis, unspecified | B38.2 | ICD10CM | respiratory |
| **1225** | pneumonia | Pneumonia due to Escherichia coli | J15.5 | ICD10CM | respiratory |
| **1226** | respiratory | Acute idiopathic pulmonary hemorrhage in infants | R04.81 | ICD10CM | respiratory |
| **1227** | respiratory | Asphyxia and hypoxemia | R09.0 | ICD10CM | respiratory |
| **1228** | respiratory | Hypoxemia | R09.02 | ICD10CM | respiratory |
| **1229** | respiratory | Acute respiratory distress | R06.03 | ICD10CM | respiratory |
| **1230** | respiratory | Pain in throat | R07.0 | ICD10CM | respiratory |
| **1231** | respiratory | Sneezing | R06.7 | ICD10CM | respiratory |
| **1232** | respiratory | Hiccough | R06.6 | ICD10CM | respiratory |
| **1233** | respiratory | Hemorrhage from other sites in respiratory passages | R04.89 | ICD10CM | respiratory |
| **1234** | respiratory failure | Chronic respiratory failure with hypercapnia | J96.12 | ICD10CM | respiratory |
| **1235** | respiratory failure | Acute respiratory failure with hypoxia | J96.01 | ICD10CM | respiratory |

| | | | | | |
|---|---|---|---|---|---|
| **1236** | respiratory failure | Respiratory failure, not elsewhere classified | J96 | ICD10CM | respiratory |
| **1237** | respiratory failure | Acute and chronic respiratory failure with hypercapnia | J96.22 | ICD10CM | respiratory |
| **1238** | respiratory failure | Acute respiratory failure with hypercapnia | J96.02 | ICD10CM | respiratory |
| **1239** | respiratory failure | Acute and chronic respiratory failure, unspecified whether with hypoxia or hypercapnia | J96.20 | ICD10CM | respiratory |
| **1240** | respiratory failure | Respiratory arrest | R09.2 | ICD10CM | respiratory |
| **1241** | respiratory failure | Postprocedural respiratory failure | J95.82 | ICD10CM | respiratory |
| **1242** | respiratory failure | Respiratory failure, unspecified with hypercapnia | J96.92 | ICD10CM | respiratory |
| **1243** | respiratory failure | Respiratory failure, unspecified, unspecified whether with hypoxia or hypercapnia | J96.90 | ICD10CM | respiratory |
| **1244** | respiratory failure | Acute and chronic respiratory failure with hypoxia | J96.21 | ICD10CM | respiratory |
| **1245** | respiratory failure | Chronic respiratory failure, unspecified whether with hypoxia or hypercapnia | J96.10 | ICD10CM | respiratory |
| **1246** | respiratory failure | Acute and chronic postprocedural respiratory failure | J95.822 | ICD10CM | respiratory |
| **1247** | respiratory failure | Chronic respiratory failure with hypoxia | J96.11 | ICD10CM | respiratory |
| **1248** | respiratory failure | Respiratory failure, unspecified with hypoxia | J96.91 | ICD10CM | respiratory |
| **1249** | respiratory failure | Acute postprocedural respiratory failure | J95.821 | ICD10CM | respiratory |
| **1250** | respiratory failure | Acute respiratory failure, unspecified whether with hypoxia or hypercapnia | J96.00 | ICD10CM | respiratory |
| **1251** | ventilation | Ventilation assist and management, initiation of pressure or volume preset ventilators for assisted or controlled breathing; first day | 94656 | CPT4 | respiratory |
| **1252** | ventilation | Assistance with Respiratory Ventilation, Greater than 96 Consecutive Hours, Continuous Positive Airway Pressure | 5A09557 | ICD10PCS | respiratory |
| **1253** | ventilation | Tracheostomy, planned (separate procedure) | 31600 | CPT4 | respiratory |
| **1254** | ventilation | Intubation, endotracheal, emergency procedure | 31500 | CPT4 | respiratory |
| **1255** | ventilation | Ventilation assist and management, initiation of pressure or volume preset ventilators for assisted or controlled breathing; subsequent days | 94657 | CPT4 | respiratory |
| **1256** | ventilation | Assistance with Respiratory Ventilation, Less than 24 Consecutive Hours, Continuous Positive Airway Pressure | 5A09357 | ICD10PCS | respiratory |
| **1257** | ventilation | Continuous invasive mechanical ventilation for less than 96 consecutive hours | 96.71 | ICD9Proc | respiratory |
| **1258** | ventilation | Respiratory Ventilation, Less than 24 Consecutive Hours | 5A1935Z | ICD10PCS | respiratory |
| **1259** | ventilation | Nasal interface (mask or cannula type) used with positive airway pressure device, with or without head strap | A7034 | HCPCS | respiratory |
| **1260** | ventilation | Insertion of Endotracheal Airway into Trachea, Via Natural or Artificial Opening Endoscopic | 0BH18EZ | ICD10PCS | respiratory |
| **1261** | ventilation | Insertion of endotracheal tube | 96.04 | ICD9Proc | respiratory |
| **1262** | ventilation | Ventilation assist and management, initiation of pressure or volume preset ventilators for assisted or controlled breathing; hospital inpatient/observation, initial day | 94002 | CPT4 | respiratory |

| | | | | | |
|---|---|---|---|---|---|
| **1263** | ventilation | Respiratory Ventilation, Greater than 96 Consecutive Hours | 5A1955Z | ICD10PCS | respiratory |
| **1264** | ventilation | Tracheostomy, planned (separate procedure); younger than 2 years | 31601 | CPT4 | respiratory |
| **1265** | ventilation | Insertion of Endotracheal Airway into Trachea, Via Natural or Artificial Opening | 0BH17EZ | ICD10PCS | respiratory |
| **1266** | ventilation | Assistance with Respiratory Ventilation, Greater than 96 Consecutive Hours, Intermittent Positive Airway Pressure | 5A09558 | ICD10PCS | respiratory |
| **1267** | ventilation | Continuous invasive mechanical ventilation for 96 consecutive hours or more | 96.72 | ICD9Proc | respiratory |
| **1268** | ventilation | Assistance with Respiratory Ventilation, Less than 24 Consecutive Hours | 5A0935Z | ICD10PCS | respiratory |
| **1269** | ventilation | Respiratory Ventilation, 24-96 Consecutive Hours | 5A1945Z | ICD10PCS | respiratory |
| **1270** | ventilation | Non-invasive mechanical ventilation | 93.9 | ICD9Proc | respiratory |
| **1271** | ventilation | Ventilation assist and management, initiation of pressure or volume preset ventilators for assisted or controlled breathing; hospital inpatient/observation, each subsequent day | 94003 | CPT4 | respiratory |
| **1272** | ventilation | Continuous positive airway pressure ventilation (CPAP), initiation and management | 94660 | CPT4 | respiratory |
| **1273** | ventilation | Assistance with Respiratory Ventilation, 24-96 Consecutive Hours, Continuous Positive Airway Pressure | 5A09457 | ICD10PCS | respiratory |
| **1274** | ventilation | Tracheotomy tube change prior to establishment of fistula tract | 31502 | CPT4 | respiratory |
